# Dynamic fusion of genomics and functional network connectivity in UK biobank reveals schizophrenia-related SNP manifolds

**DOI:** 10.1101/2024.01.09.24301013

**Authors:** Jiayu Chen, Armin Iraji, Zening Fu, Marlena Duda, Pablo Andrés-Camazón, Bishal Thapaliya, Jingyu Liu, Vince D. Calhoun

## Abstract

**Background:** Many mental disorders show strong genetic influence. In parallel, dynamic functional network connectivity (dFNC) has shown high sensitivity to brain changes related to mental disorders. However, previous studies linking dFNC to genetics largely follow a paradigm to identify associations between one set of genetic factors and multiple sets of connectivity features from different dFNC states, ignoring the potential variability in genetic correlates across states.

**Methods:** We propose a novel joint ICA (jICA)-based “dynamic fusion” framework to identify dynamically-tuned genetic manifolds. A sliding window approach was utilized to estimate four dFNC states and compute subject-level state-average dFNC (sa-dFNC) features. The sa-dFNC features of each state were combined with schizophrenia risk SNPs within a jICA fusion framework, resulting in four parallel fusions in 32861 individuals of the UK Biobank cohort. The extracted four sets of joint SNP-dFNC components were further validated for clinical relevance in a combined schizophrenia cohort of 1237 individuals (528 patients). The similarity of SNP-dFNC components across four parallel fusions was evaluated as a measure of state variability.

**Results:** We observed a mixture of “state-invariant” and “state-variant” components for SNP and dFNC modalities. Particularly, the schizophrenia-related state-variant SNP components, or manifolds, complemented each other by capturing different genes involved in the same biological functions, revealing a partition of genomic risk particularly elicited by the dynamics of brain function.

**Conclusions:** By augmenting the SNP factors to state-variant manifolds, this dynamic fusion framework promises additional insights into underlying genetic risk of disease-related alterations in dynamic brain function.

## Introduction

It has been well established that many psychiatric and neurological disorders show significant heritability^1-4^. Family and twin studies have suggested heritability rates ranging from 0.80 for schizophrenia (SZ)^1^, to 0.90 for autism (ASD)^5^, and 0.60-0.80 for Alzheimer’s disease (AD)^6^. More recently, large-scale genome-wide association studies (GWASs) have confirmed the polygenic nature and significant estimates have been consistently reported across studies for common-variant-based heritability, varying from 0.10 for AD to 0.28 for obsessive-compulsive disorder (OCD), and consistently 0.23-0.24 for SZ^7,8^. Understanding how genetic factors influence predisposition, disease progression, and response to treatment is crucial in advancing personalized healthcare and precision medicine^9-11^.

A common notion is that in brain disorders, genetic factors exert influence on clinical manifestations by affecting the brain. The past decade has seen great efforts to unravel brain disorders by examining the relationship between genetic variants and changes in the brain, known as imaging genomics^12^, so that brain disorders may be described from a multi-scale multi-dimensional perspective for a comprehensive understanding of its underlying biology^13-15^. The hope is that such a dimensional framework would allow detecting and quantifying risk at an early stage and stratifying individuals based on biological underpinnings, ultimately leading to more efficient individualized treatment^16,17^.

Dynamic functional network connectivity (dFNC) measures time-resolved functional coupling between brain networks and has proven to identify disease-related changes in the brain^18-21^. Compared to statis functional connectivity^22-24^, dFNC provides rich information that describes how functional interactions vary across time. One well-established strategy to characterize dFNC is the sliding window approach, which captures the flow of functional connectivity through a sequence of windowed FNCs (wFNCs)^25^, and allows for estimating stable representative connectivity patterns, known as dFNC states^26,27^, for further investigation on clinical relevance^18,28^. The dFNC approach has proven to yield highly informative neurobiological markers for disease discrimination, e.g., classifying schizophrenia, bipolar disorder, autism, and mild traumatic brain injury^29-31^, and significantly outperform sFNC in classifying patients with SZ and bipolar disorder^29^.

While dFNC effectively captures disease-related alterations, its genetic correlates are understudied. The GWASs of brain imaging phenotypes conducted on the UKB data indicate that despite not showing as high heritability as structural phenotypes, 235 out of 1771 tested functional connectivity pairs showed significant heritability^32^. In addition, when ICA was applied to the 1771 connectivity pairs, the resulting connectivity components showed much stronger heritability^32^. These observations motivate further elucidations on genetic profiles underlying functional connectivity, particularly those disrupted in SZ. Prior work linking genetics to dFNC typically follow a paradigm to identify associations between one set of single nucleotide polymorphism (SNP) factors and multiple sets of connectivity features from different dynamic states. While this is a powerful approach, it ignores the possibility that there might be locally optimizable SNP-dFNC factors, such that unique SNP factors might be discovered by separately linking SNPs to each dFNC state.

The current work aims to address the gap by proposing a novel jICA-based “dynamic fusion” framework to identify manifolds of SNP-dFNC association through parallel data fusions between SZ-risk SNPs and individual dFNC states. We leveraged the most recent psychiatric genomic consortium GWAS of SZ to select out SNPs conferring risk for SZ^33^ and jointly analyzed them with dFNC features estimated using the NeuroMark pipeline^34^ in the UKB cohort. The identified joint SNP-dFNC components were further investigated for SZ relevance in a combined SZ cohort. We hypothesized that the joint SNP-dFNC components would show different levels of state variability. To the best of our knowledge, this is the first work exploring the dynamically-tuned SNP decompositions across multiple data fusions with dFNC. We hope that linking SNPs to dynamic functional brain information augments the SNP modality to across-state manifolds, and thus provides a unique lens to elicit unique SNP correlates missed in previous work.

## Methods and Materials

### Participants

#### UK Biobank

The current work leveraged the population-based UKB cohort which recruited more than 500K individuals across the United Kingdom ^35,36^. The UKB study was approved by the North West Haydock Research Ethics Committee, and the data used in our work were obtained under application 34175. Specifically, we used the imputed SNP data and resting-state functional magnetic resonance images (rsfMRI) of 32681 European ancestry individuals with both imaging and genetic modalities available after quality control (QC), including 15357 males and 17504 females, aged between 45-81 with a median of 64.

#### SZ cohorts

Additional SZ cohorts were utilized to validate the UKB findings for SZ relevance. For this purpose, four SZ cohorts were combined to boost power, including COBRE^37^, FBIRN^20^, B-SNIP1^38^, and MPRC^39^, resulting in 1237 individuals with rsfMRI data after QC, including 528 individuals with SZ and 709 controls (740 males and 497 females, aged between 16-79 with a median of 38). The institutional review board at each site approved the study, and all participants provided written informed consent. Details regarding recruitment and data collection can be found in previous publications^20,39-42^.

### SNP preprocessing and pre-screening

#### SNP data

The imputed SNP data released by UKB consisted of 487,320 individuals and ∼96 million variants (v3_s487320). Details of genotyping and imputation can be found in the paper that describes the UKB genomic data^35^.

#### Preprocessing of SNP

In this study, we first identified the participants that passed the UKB quality control (sex mismatch, missing rate, and heterozygosity) and also had rsfMRI data available. We excluded SNPs with minor allele frequencies < 0.01^43^. Individual relatedness (identify-by-descent) was estimated using PLINK^44^. For each group of individuals who were second-degree relatives or closer, only one individual was randomly selected and retained for subsequent analysis. Finally, we identified individuals of European ancestry to be those close (< 3SD) to the center of the ‘white’ cluster as defined by the top four principal components^43,45^.

#### Pre-screening

We selected SZ risk SNPs based on the 287 loci identified by PGC SCZ2022^33^, which were further pruned at *r^2^* < 0.2, a commonly used threshold for European populations^46^, resulting in a total of 12946 SNPs included in the dynamic fusion. Before fusion, univariate regression was conducted to remove sex and dummy-coded site effects, as well as the top four principal components of the genomic SNP data to control for population stratification^43,45^.

### Estimation of dynamic functional network connectivity

#### rsfMRI data

The data we obtained contain rsfMRI scans of ∼38K individuals. UKB used identical scanner models, coils, software, and protocols across centers to ensure data harmonization as much as possible^36^. The rsfMRI scan parameters are as follows: resolution = 2.4 mm × 2.4 mm × 2.4 mm, matrix = 88 × 88 × 64, TE = 39 ms, TR = 735 ms, alpha = 51°, multi-band factors = 8, duration = 6:10 mins^36^. For the SZ data, detailed scanning parameters of each cohort can be found in supplemental text 1 (**ST1)**.

#### Preprocessing of rsfMRI

We downloaded the preprocessed UKB data, normalized the images to standard MNI space (http://www.mni.mcgill.ca/) and resampled to 3 mm × 3 mm × 3 mm voxels using the SPM nonlinear registration, followed by smoothing using a 6mm FWHM Gaussian kernel. The rsfMRI data of the SZ cohorts were preprocessed using the fully automated NeuroMark pipeline (SPM12-based)^34,47^ (see **ST2** for details).

#### Estimation of windowed FNC

The Neuromark_fMRI_1.0 network template^34,47^ served as a reference in a spatial-constrained independent component analysis (scICA)^48^ to derive 53 subject-level intrinsic connectivity networks (ICNs) and their associated time courses (TCs). The subject-level TCs then underwent additional processing steps, including detrending, despiking, regressions of the six realignment parameters and their temporal derivatives, and band-pass filtering with 0.01–0.15 Hz. Windowed FNC was then estimated based on the processed TCs of 53 ICNs as described in our previous work^18,34^. In brief, a tapered sliding window (convolving a rectangular window of 20 TRs) was used to estimate covariance from the regularized precision matrix, resulting in a sequence of wFNCs of 1378 FNC pairs for each individual in the UKB and SZ cohorts.

#### Estimation of dFNC states

The wFNCs of all UKB individuals were concatenated, on which K-means clustering was applied to identify clusters of similar connectivity patterns across time windows and individuals, interpreted as dFNC states. In the current work, we identified four dynamic states, where the optimal number of states was determined as four based on the elbow method.

#### Subject-level dFNC features

For all the individuals in both UKB and SZ data, we assigned each wFNC to the nearest dFNC state based on its Euclidean distance to the state centroids. For each individual, wFNCs assigned to the same state were averaged to yield the most representative FNC patterns of this individual in a dynamic state, denoted as subject-specific state-average dFNC (sa-dFNC) in the following text. In total, four sets of sa-dFNCs were generated and used in the fusion analysis. As not all the individuals experienced all four states during the length of scan, sample sizes of dynamic fusion varied across states. Specifically, 31814/32775/17929/27497 individuals were utilized for SNP fusion with states 1/2/3/4, respectively in UKB, and 1018/1199/954/788 individuals were used in the SZ data to validate the dFNC findings for each state. For each connectivity feature, regression was conducted to remove age, sex, and dummy-coded site effects.

### Dynamic fusion

Figure 1 shows the diagram of dynamic fusion to link the same set of SZ risk SNPs separately with each sa-dFNC matrix via jICA^49^. JICA builds upon the Infomax algorithm, which achieves data decomposition by detecting features covarying with each other and clustering them into components following super-Gaussian distributions^50^. In this context, jICA captures joint SNP-dFNC components that are maximally independent, with each component exhibiting similar SNP and dFNC variations across subjects. By decomposition **X = AS**, jICA decomposes the data **X** (i.e., combined data matrix of X_SNP_ and X_dFNC_) into a linear combination of a set of source components (**S**) and their associated loadings (**A**). **S** largely reflects how individual SNP and dFNC features are linearly weighted to collectively define an independent source, where heavily weighted features contribute more to component characteristics that differentiate from others. **A** reflects how source components are loaded on each subject. To identify dynamically tuned SNP manifolds, we conducted four parallel SNP-dFNC fusions on the UKB data. For each fusion, the feature matrices (12946 SNPs and 1378 connectivity pairs) were concatenated horizontally for jICA decomposition^50^. The component number was estimated to be 35 based on the PCA scree plot. Finally we obtained four sets of joint SNP-dFNC components and included only super-Gaussian components (see **ST3** for details) to further investigate into state variability of SNP manifolds.

**Figure 1:**
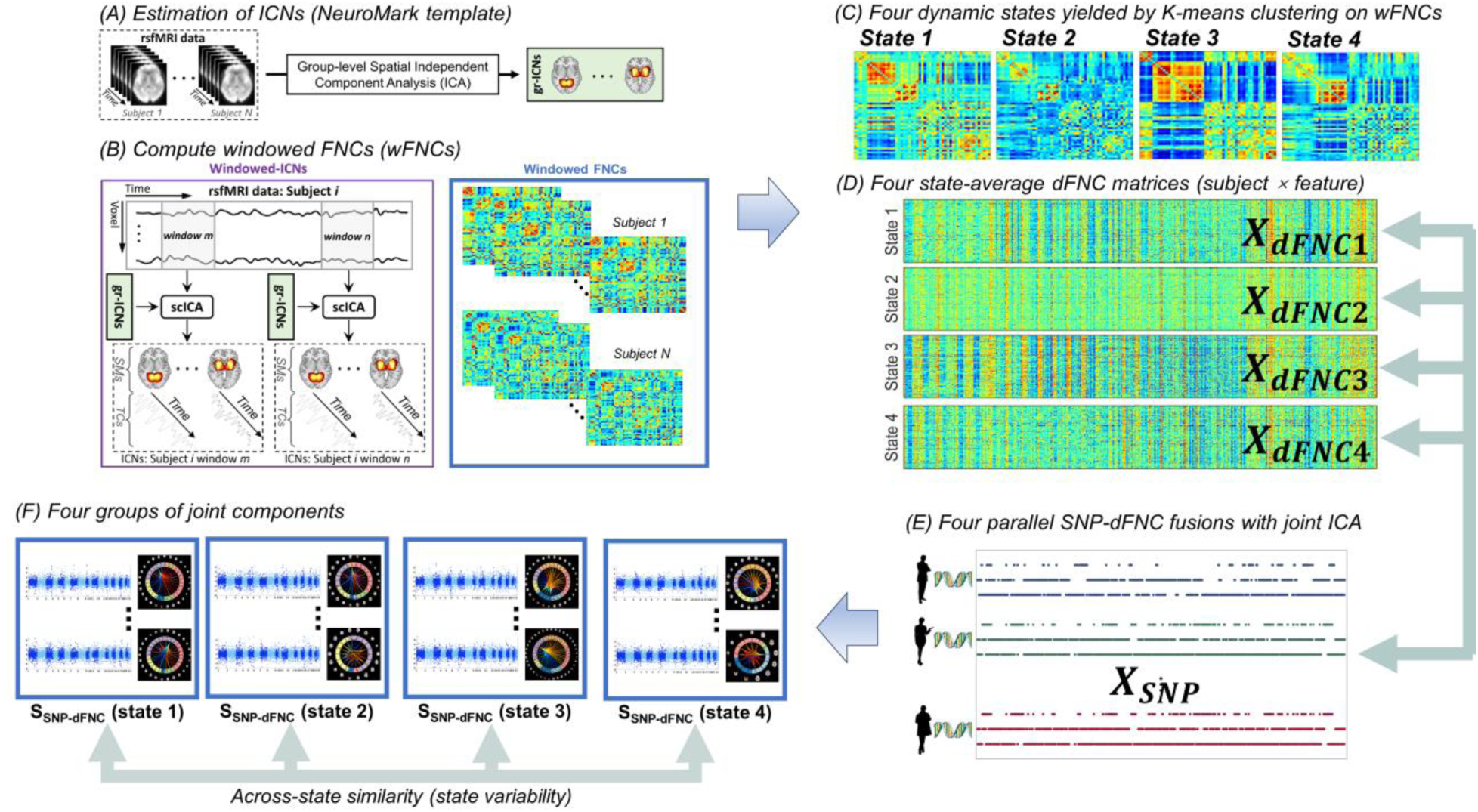
A diagram of dynamic fusion. The SNP data X_SNP_ was fused with the dFNC features of each state, e.g., X_dFNC_ (state1), in parallel. S_SNP-dFNC_ denotes the joint SNP-dFNC components yielded by the dynamic fusion. Across-state similarity was assessed between the four sets of components to indicate state variability, for SNP and dFNC modalities, respectively.

We conducted a three-fold stability test, where the full sample was randomly partitioned into three subsets which were expected to be still powered for SNP-dFNC fusion. The joint components extracted from the full sample were compared with those obtained from the subsets for stability (measured by Pearson correlation). We also conducted unimodal ICA on the SNP data, as well as jICA on combined SNP and sa-dFNC features of state 1 and state 2 with similar sample sizes. The results were compared with those of the dynamic fusion to examine the specificity of SNP manifolds.

#### SZ relevance

While the SZ-relevance of SNPs was grounded by the GWAS, the SZ-relevance of dFNC findings remained to be validated. For this purpose, we projected the dFNC elements onto the sa-dFNC features of the combined SZ data: 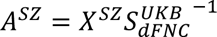. The resulting loadings (*A_SZ_*) were then investigated for differences between individuals with SZ and controls using a two-sample t-test after regressing out age, sex, and dummy-coded site effects. Significant SZ-discriminating dFNC elements were identified with FDR-corrected *p* < 0.05 across all the four sets of elements.

#### Interpretation of joint components

First, we evaluated the components’ across-state similarity (Figure 1f). For each SNP-dFNC component of a given state-specific fusion, its most similar SNP and dFNC elements were identified in each of the other three fusions based on the correlations of **S**. The correlation-based similarity measures were then averaged across the three most similar SNP and dFNC elements to reflect the overall across-state similarity. Lower similarity indicates a multivariate SNP/dFNC factor shows more state variability, suggesting high state-specificity. Second, for all the joint components whose dFNC elements showed significant SZ discrimination, the SNP/dFNC elements were thresholded at |z-score| > 3 to identify heavily weighted top SNPs and connectivity pairs that were more tightly coupled with each other. The top connectivity pairs were then interpreted based on the involved ICNs. The top SNPs were annotated to genes and investigated for enrichment in biological processes using Gene Ontology^51^.

## Results

We obtained four sets of joint SNP-dFNC components from the dynamic parallel fusions, each set consisting of 35 joint components extracted from 12946 SZ risk SNPs and 1378 sa-dFNC features of one dynamic state. Overall these joint components were highly stable in the three-fold test, presenting similar patterns as those obtained from the subset samples (mean correlation = 0.67, SD = 0.23).

By projecting dFNC elements to the SZ cohort, 53 joint components were validated as SZ-relevant, as summarized in Table 1, where the percentage of explained variance (*r^2^*) ranged between 0.46% and 8.18%. For each component, its most similar counterpart across the other three fusions was also listed, along with the SNP and dFNC similarities observed in that counterpart.

**Table 1:** Summary of SZ-relevant SNP-dFNC components.

| State | Comp | r<sup>2</sup> | p | FDR q | State_similar | Comp_similar | SNP similarity | dFNC similarity |
| --- | --- | --- | --- | --- | --- | --- | --- | --- |
| 1 | 2 | 0.86% | 3.02E-03 | 1.03E-02 | 3 | 22 | 0.06 | 0.57 |
| 1 | 3 | 1.41% | 1.48E-04 | 9.04E-04 | 4 | 23 | 0.26 | 0.47 |
| **1** | **4** | **1.32%** | **2.37E-04** | **1.27E-03** | **4** | **19** | **0.09** | **0.49** |
| **1** | **6** | **0.63%** | **1.15E-02** | **3.11E-02** | **4** | **14** | **0.07** | **0.39** |
| **1** | **9** | **5.34%** | **8.24E-14** | **3.84E-12** | **3** | **7** | **0.33** | **0.67** |
| 1 | 10 | 1.19% | 4.89E-04 | 2.21E-03 | 4 | 29 | 0.15 | 0.67 |
| 1 | 12 | 1.26% | 3.36E-04 | 1.57E-03 | 3 | 18 | 0.31 | 0.78 |
| **1** | **13** | **1.47%** | **1.06E-04** | **7.09E-04** | **3** | **5** | **0.35** | **0.78** |
| 1 | 16 | 1.43% | 1.33E-04 | 8.47E-04 | 4 | 17 | 0.08 | 0.41 |
| 1 | 19 | 1.61% | 4.98E-05 | 3.67E-04 | 2 | 2 | 0.42 | 0.62 |
| 1 | 29 | 0.90% | 2.48E-03 | 8.90E-03 | 2 | 29 | 0.98 | 0.19 |
| **1** | **30** | **1.35%** | **2.08E-04** | **1.16E-03** | **2** | **32** | **1.00** | **0.57** |
| 1 | 33 | 0.57% | 1.58E-02 | 3.97E-02 | 2 | 33 | 0.99 | 0.35 |
| 2 | 2 | 0.71% | 3.55E-03 | 1.16E-02 | 1 | 19 | 0.42 | 0.62 |
| 2 | 4 | 0.57% | 8.90E-03 | 2.49E-02 | 1 | 7 | 0.21 | 0.49 |
| 2 | 5 | 1.11% | 2.60E-04 | 1.35E-03 | 4 | 20 | 0.15 | 0.63 |
| 2 | 8 | 0.46% | 1.87E-02 | 4.43E-02 | 4 | 22 | 0.16 | 0.64 |
| 2 | 9 | 2.99% | 1.72E-09 | 6.01E-08 | 1 | 18 | 0.25 | 0.60 |
| 2 | 12 | 1.17% | 1.71E-04 | 9.98E-04 | 1 | 14 | 0.18 | 0.50 |
| 2 | 22 | 4.60% | 6.21E-14 | 3.84E-12 | 4 | 15 | 0.43 | 0.86 |
| 2 | 25 | 1.95% | 1.24E-06 | 1.45E-05 | 1 | 20 | 0.33 | 0.80 |
| 2 | 27 | 0.62% | 6.56E-03 | 2.00E-02 | 1 | 15 | 0.56 | 0.81 |
| 2 | 28 | 0.58% | 8.43E-03 | 2.46E-02 | 4 | 13 | 0.17 | 0.61 |
| 2 | 29 | 0.51% | 1.31E-02 | 3.47E-02 | 1 | 29 | 0.98 | 0.19 |
| 2 | 30 | 1.09% | 2.97E-04 | 1.44E-03 | 1 | 27 | 0.98 | 0.23 |
| 2 | 31 | 1.42% | 3.61E-05 | 2.81E-04 | 1 | 34 | 0.99 | 0.33 |
| 2 | 32 | 2.91% | 2.84E-09 | 7.95E-08 | 1 | 30 | 1.00 | 0.57 |
| 2 | 35 | 1.10% | 2.80E-04 | 1.40E-03 | 1 | 35 | 1.00 | 0.30 |
| 3 | 2 | 2.95% | 9.25E-08 | 1.62E-06 | 4 | 5 | 0.98 | 0.76 |
| 3 | 3 | 1.21% | 6.79E-04 | 2.88E-03 | 4 | 9 | 0.13 | 0.60 |
| 3 | 4 | 1.88% | 2.16E-05 | 1.78E-04 | 4 | 14 | 0.08 | 0.52 |
| 3 | 5 | 3.33% | 1.40E-08 | 3.26E-07 | 1 | 13 | 0.35 | 0.78 |
| 3 | 7 | 8.18% | 2.06E-19 | 2.88E-17 | 1 | 9 | 0.33 | 0.67 |
| 3 | 8 | 2.11% | 6.76E-06 | 6.31E-05 | 2 | 11 | 0.25 | 0.60 |
| 3 | 9 | 2.31% | 2.37E-06 | 2.56E-05 | 4 | 2 | 0.02 | 0.59 |
| 3 | 11 | 1.23% | 5.93E-04 | 2.60E-03 | 4 | 12 | 0.15 | 0.60 |
| 3 | 15 | 2.68% | 3.71E-07 | 5.19E-06 | 4 | 18 | 0.07 | 0.66 |
| 3 | 17 | 0.86% | 4.17E-03 | 1.33E-02 | 4 | 23 | 0.26 | 0.55 |
| 3 | 21 | 1.06% | 1.44E-03 | 5.61E-03 | 1 | 22 | 0.15 | 0.76 |
| 3 | 22 | 2.99% | 7.61E-08 | 1.52E-06 | 1 | 2 | 0.06 | 0.57 |
| 3 | 23 | 0.98% | 2.25E-03 | 8.28E-03 | 4 | 13 | 0.03 | 0.56 |
| 3 | 24 | 2.72% | 2.97E-07 | 4.62E-06 | 1 | 8 | 0.18 | 0.56 |
| 3 | 28 | 0.61% | 1.59E-02 | 3.97E-02 | 1 | 31 | 0.90 | 0.18 |
| 3 | 30 | 0.93% | 2.82E-03 | 9.86E-03 | 2 | 32 | 0.99 | 0.34 |
| 3 | 31 | 0.59% | 1.78E-02 | 4.29E-02 | 2 | 30 | 0.94 | 0.13 |
| 4 | 2 | 1.94% | 8.86E-05 | 6.20E-04 | 3 | 9 | 0.02 | 0.59 |
| 4 | 9 | 0.76% | 1.42E-02 | 3.69E-02 | 3 | 3 | 0.13 | 0.60 |
| 4 | 15 | 2.58% | 5.96E-06 | 5.96E-05 | 2 | 22 | 0.43 | 0.86 |
| 4 | 18 | 0.90% | 7.70E-03 | 2.29E-02 | 3 | 15 | 0.07 | 0.66 |
| 4 | 23 | 1.08% | 3.56E-03 | 1.16E-02 | 3 | 17 | 0.26 | 0.55 |
| 4 | 30 | 0.72% | 1.73E-02 | 4.25E-02 | 1 | 29 | 0.96 | 0.15 |
| 4 | 31 | 1.20% | 2.12E-03 | 8.01E-03 | 2 | 33 | 0.97 | 0.15 |
| 4 | 35 | 0.87% | 8.71E-03 | 2.49E-02 | 2 | 35 | 0.99 | 0.28 |

The top SNPs and connectivity pairs of SZ-relevant components were identified. Supplemental Table 1 provides the list of the genes annotated to the top SNPs. The top connectivity pairs were characterized by the anatomical labels of the two involved ICNs, as summarized in supplemental Table 2. In addition, Manhattan and connectogram plots were generated for the SNP and dFNC elements, respectively (see supplemental figures). Finally, for those SNP elements exhibiting a significant enrichment in pathway analysis, we summarized the involved major biological processes in Table 2.

**Table 2:** Summary of enriched biological processes identified from SZ-relevant SNP-dFNC components.

| State | Indcomp | GO pathway | Fold enrichment | FDR q |
| --- | --- | --- | --- | --- |
| 1 | 3 | regulation of signaling (GO:0023051) | 2.29 | 2.02E-02 |
|  |  | negative regulation of locomotion (GO:0040013) | 6.45 | 3.31E-02 |
|  |  | neurogenesis (GO:0022008) | 3.31 | 3.39E-02 |
|  |  | positive regulation of cell projection organization (GO:0031346) | 6.84 | 3.80E-02 |
|  |  | neuron development (GO:0048666) | 3.94 | 3.83E-02 |
| 1 | 4 | positive regulation of cell projection organization (GO:0031346) | 8.65 | 1.08E-03 |
| 1 | 6 | neurogenesis (GO:0022008) | 3.91 | 1.92E-03 |
|  |  | regulation of synaptic plasticity (GO:0048167) | 9.63 | 1.96E-02 |
| 1 | 9 | positive regulation of cell projection organization (GO:0031346) | 7.11 | 4.14E-03 |
|  |  | regulation of synaptic plasticity (GO:0048167) | 10.45 | 4.43E-03 |
|  |  | learning (GO:0007612) | 10.27 | 1.59E-02 |
|  |  | memory (GO:0007613) | 10.9 | 4.70E-02 |
| 1 | 12 | positive regulation of cell projection organization (GO:0031346) | 8.89 | 8.05E-04 |
|  |  | enzyme-linked receptor protein signaling pathway (GO:0007167) | 5.37 | 4.06E-03 |
|  |  | anatomical structure morphogenesis (GO:0009653) | 2.94 | 4.15E-03 |
|  |  | taxis (GO:0042330) | 6.02 | 4.37E-03 |
|  |  | locomotion (GO:0040011) | 5.8 | 4.45E-03 |
|  |  | multicellular organism development (GO:0007275) | 2.31 | 4.60E-03 |
|  |  | chemotaxis (GO:0006935) | 6.04 | 4.82E-03 |
|  |  | regulation of neuron projection development (GO:0010975) | 6.36 | 5.05E-03 |
|  |  | axon guidance (GO:0007411) | 8.56 | 1.61E-02 |
|  |  | ephrin receptor signaling pathway (GO:0048013) | 22.88 | 2.16E-02 |
|  |  | regulation of calcium ion transport (GO:0051924) | 7.61 | 2.30E-02 |
|  |  | neurogenesis (GO:0022008) | 3.25 | 2.35E-02 |
| 1 | 13 | positive regulation of molecular function (GO:0044093) | 3.19 | 1.60E-02 |
|  |  | synapse organization (GO:0050808) | 7.28 | 2.23E-02 |
|  |  | regulation of neuron projection development (GO:0010975) | 5.38 | 2.95E-02 |
|  |  | regulation of calcium ion transport (GO:0051924) | 7.37 | 3.07E-02 |
| 1 | 29 | general adaptation syndrome (GO:0051866) | > 100 | 2.15E-02 |
| 2 | 2 | regulation of intracellular signal transduction (GO:1902531) | 3.21 | 2.15E-03 |
|  |  | learning or memory (GO:0007611) | 7.94 | 3.65E-03 |
|  |  | positive regulation of cell projection organization (GO:0031346) | 5.63 | 3.24E-02 |
|  |  | cardiac muscle cell contraction (GO:0086003) | 20.6 | 4.16E-02 |
|  |  | regulation of long-term synaptic depression (GO:1900452) | 41.52 | 4.49E-02 |
| 2 | 5 | organic substance transport (GO:0071702) | 2.78 | 7.51E-03 |
|  |  | establishment of localization (GO:0051234) | 2.17 | 8.82E-03 |
|  |  | regulation of monoatomic ion transport (GO:0043269) | 4.82 | 2.46E-02 |
| 2 | 9 | calcium ion import (GO:0070509) | 60.21 | 1.82E-02 |
| 2 | 22 | positive regulation of cell projection organization (GO:0031346) | 6.92 | 3.41E-02 |
| 2 | 25 | regulation of trans-synaptic signaling (GO:0099177) | 6.44 | 3.08E-03 |
|  |  | regulation of calcium ion transmembrane transport (GO:1903169) | 11.33 | 3.68E-03 |
|  |  | learning (GO:0007612) | 10.03 | 3.34E-02 |
|  |  | cardiac muscle cell action potential (GO:0086001) | 21.72 | 3.61E-02 |
| 2 | 27 | regulation of lyase activity (GO:0051339) | 17.65 | 3.34E-02 |
|  |  | regulation of cell projection organization (GO:0031344) | 4.64 | 3.41E-02 |
|  |  | positive regulation of collateral sprouting (GO:0048672) | 69.33 | 3.97E-02 |
|  |  | positive regulation of axonogenesis (GO:0050772) | 16.3 | 4.13E-02 |
|  |  | synapse organization (GO:0050808) | 6.87 | 5.94E-02 |
| 3 | 1 | regulation of voltage-gated calcium channel activity (GO:1901385) | > 100 | 3.99E-02 |
| 3 | 2 | regulation of cell projection organization (GO:0031344) | 5.15 | 2.81E-02 |
| 3 | 4 | regulation of trans-synaptic signaling (GO:0099177) | 6.32 | 3.28E-02 |
|  |  | modulation of chemical synaptic transmission (GO:0050804) | 6.34 | 6.45E-02 |
| 3 | 5 | negative regulation of lyase activity (GO:0051350) | 41.18 | 2.37E-02 |
| 3 | 7 | cell-cell signaling (GO:0007267) | 3.97 | 1.66E-02 |
|  |  | system development (GO:0048731) | 2.37 | 2.68E-02 |
|  |  | positive regulation of cell projection organization (GO:0031346) | 6.89 | 2.83E-02 |
|  |  | multicellular organism development (GO:0007275) | 2.19 | 2.95E-02 |
| 3 | 8 | regulation of cell projection organization (GO:0031344) | 5.17 | 1.75E-03 |
|  |  | calcium ion transmembrane transport (GO:0070588) | 8.42 | 2.46E-02 |
| 3 | 9 | regulation of synaptic plasticity (GO:0048167) | 10.6 | 2.06E-02 |
|  |  | regulation of neuron projection development (GO:0010975) | 5.97 | 4.70E-02 |
|  |  | regulation of heart rate by cardiac conduction (GO:0086091) | 29.12 | 4.75E-02 |
| 3 | 15 | action potential (GO:0001508) | 18.16 | 2.83E-03 |
|  |  | cardiac muscle cell contraction (GO:0086003) | 29.56 | 6.33E-03 |
|  |  | regulation of calcium ion transport (GO:0051924) | 7.73 | 1.91E-02 |
| 3 | 17 | cell projection organization (GO:0030030) | 3.83 | 1.76E-03 |
|  |  | locomotion (GO:0040011) | 5.63 | 2.07E-03 |
|  |  | neuron development (GO:0048666) | 4.43 | 2.33E-03 |
|  |  | positive regulation of neurogenesis (GO:0050769) | 8.47 | 5.29E-03 |
|  |  | regulation of long-term synaptic depression (GO:1900452) | 47.67 | 1.89E-02 |
| 3 | 21 | regulation of trans-synaptic signaling (GO:0099177) | 5.83 | 2.53E-02 |
|  |  | regulation of adenylate cyclase activity (GO:0045761) | 21.45 | 2.73E-02 |
|  |  | regulation of synaptic plasticity (GO:0048167) | 9.15 | 3.15E-02 |
|  |  | glutamate receptor signaling pathway (GO:0007215) | 22.88 | 3.75E-02 |
|  |  | calcium ion transmembrane transport (GO:0070588) | 8.38 | 3.80E-02 |
| 3 | 22 | cell-cell signaling (GO:0007267) | 3.96 | 1.69E-02 |
|  |  | calcium ion transmembrane transport (GO:0070588) | 9.46 | 2.05E-02 |
|  |  | atrial cardiac muscle cell action potential (GO:0086014) | 54.48 | 4.20E-02 |
| 3 | 24 | regulation of cell projection organization (GO:0031344) | 4.43 | 1.92E-02 |
|  |  | anatomical structure morphogenesis (GO:0009653) | 2.32 | 4.94E-02 |
|  |  | regulation of calcium-mediated signaling (GO:0050848) | 12.68 | 5.01E-02 |
|  |  | learning or memory (GO:0007611) | 5.96 | 5.04E-02 |
|  |  | regulation of neuron projection development (GO:0010975) | 4.62 | 5.27E-02 |
| 4 | 18 | regulation of cell communication (GO:0010646) | 2.25 | 9.98E-03 |
|  |  | localization (GO:0051179) | 1.95 | 1.95E-02 |
|  |  | actin filament-based movement (GO:0030048) | 13.84 | 2.08E-02 |
|  |  | regulation of neuron projection development (GO:0010975) | 4.87 | 4.18E-02 |

Figure 2a shows the state variability of SNP and dFNC elements respectively, as measured by the average across-state similarity and plotted in a sorted manner for each state. Note that for a joint component, its SNP and dFNC elements might present different levels of state variability. Overall we observed a mixture of state-invariant and state-variant components for both SNP and dFNC modalities. The SZ-relevant components were highlighted by asterisks, which did not appear to be enriched in the high or low end of state variability.

**Figure 2:**
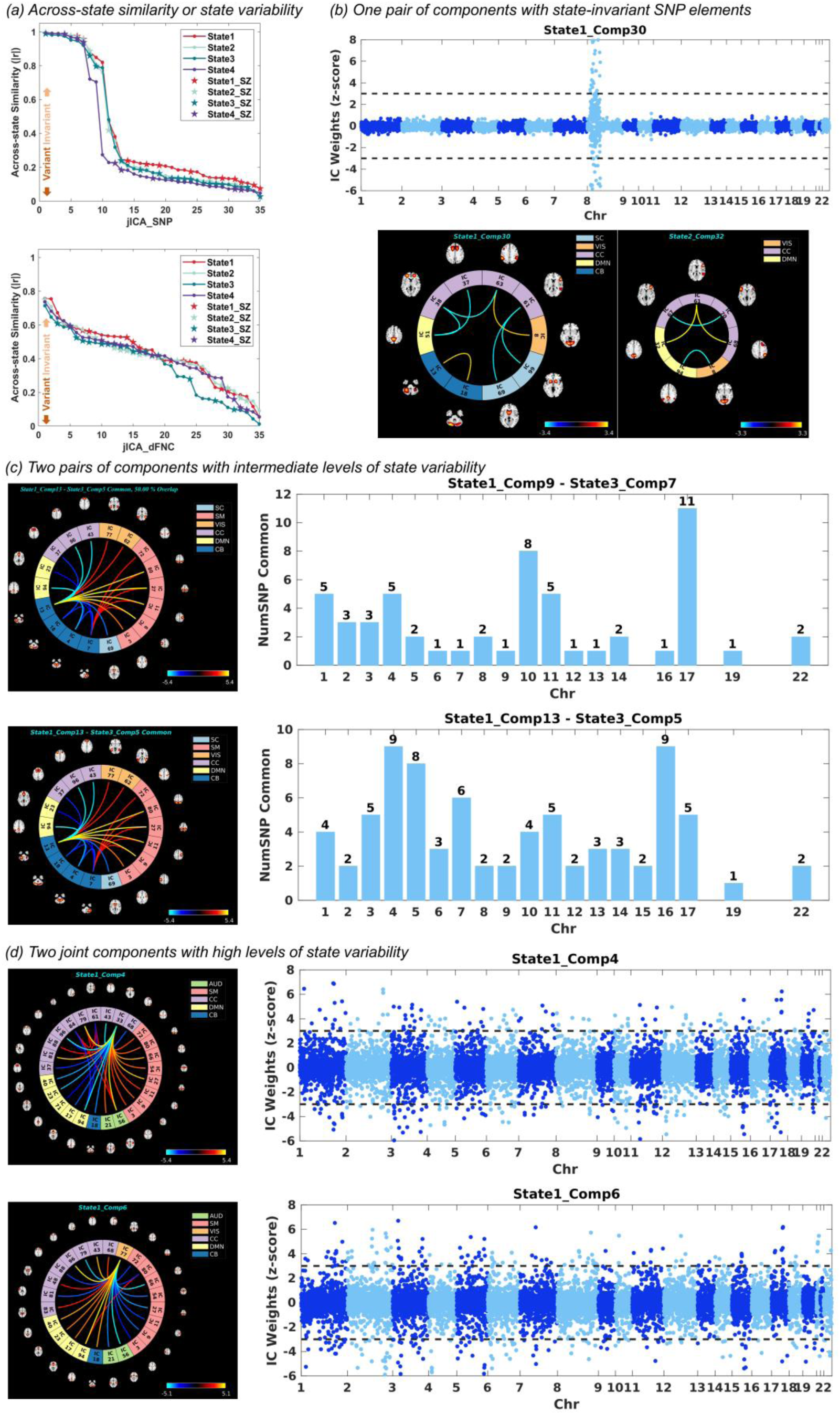
**(a)** Across-state similarity of SNP and dFNC elements respectively, where asterisks highlight SZ relevance. **(b)** Example of one pair of joint components with state-invariant SNP elements, including the Manhattan plot of State1_Comp30 where dashed lines indicate |z-score| > 3, and the connectogram plots of State1_Comp30 and State2_Comp32. **(c)** Example of two pairs of joint components with intermediate levels of state variability, State1_Comp9 - State3_Comp7, and State1_Comp13 - State3_Comp5. The bar plots and connectogram plots show the top SNPs and connectivity features shared between the paired components. **(d)** Example of two joint components with high levels of state variability. The Manhattan and connectogram plots show the top SNPs and top connectivity features identified from State1_Comp4 and State1_Comp6 respectively.

For illustrative purposes, Figure 2b presents the Manhattan plot of component 30 of SNP-dFNC_state1_ fusion (denoted as State1_Comp30), as well as the connectograms of State1_Comp30 (lower left) and State2_Comp32 (lower right), serving as an example of two fusions yielding joint components with highly similar SNP correlates (correlation = 1, as highlighted in Table 1). Only the SNP component of State1_Comp30 was plotted in Figure 2b to avoid redundancy, which was sparse and reflected the effects of a small set of top SNPs in chromosome 8. The annotated three genes were 100% overlapping between State1_Comp30 and State2_Comp32, with no significant pathway enrichment. Meanwhile, the dFNC elements showed an intermediate level of similarity. Despite not sharing top connectivity pairs, similar connections were noted between cognitive control (CC) and default mode network (DMN) ICNs.

Figure 2c shows two examples of parallel fusions yielding pairs of SZ-relevant components of an intermediate levels of similarity. The upper panel of Figure 2c presents the paired State1_Comp9 and State3_Comp7. An overlapping ratio of 37% was noted between their annotated genes, with a bar plot showing how the overlapping top SNPs were distributed across the genome. As shown in Table 2, both State1_Comp9 and State3_Comp7 were significantly enriched for the regulation of cell projection organization. Other enriched pathways included synaptic plasticity, learning, and memory for State1_Comp9, and cell signaling and system development for State3_Comp7. Figure 2c also shows the connectogram of the overlapping top connectivity features, which shared similar thalamus- and subthalamus/hypothalamus-hub connections to other ICNs (see Table S2 for labels of the ICNs).

State1_Comp13 and State3_Comp5 make another pair of components showing an intermediate level of state variability. The lower panel of Figure 2c shows the bar plot of the overlapping top SNPs and the connectogram plot of the overlapping connectivity features of these two components. An overlapping ratio of 31% was observed between the top genes. State1_Comp13 was significantly enriched for neuron projection, synapse organization, and calcium ion transport, while State3_Comp5 was enriched for the regulation of lyase activity. Both dFNC elements highlighted cerebellum connections to other ICNs.

Finally, Figure 2d shows State1_Comp4 and State1_Comp6 as two examples showing a high level of state variability, and therefore considered as state-variant manifolds. The SNP element of State1_Comp4 was significantly enriched for regulation of cell projection, and the dFNC element reflected insula connections to other ICNs. For State1_Comp6, its SNP element was significantly enriched for neurogenesis and synaptic plasticity with the dFNC element capturing connections of the middle temporal gyrus (MTG) with other ICNs.

Notably, the parallel dynamic fusions yielded different SNP components compared to applying unimodal ICA on the SNP data. Specifically, 9 out of 35 unimodal SNP components were able to find counterparts in dynamic fusion with correlation > 0.85, echoing the aforementioned state-invariant components. For the remaining unimodal SNP components, the similarity with dynamic fusion ranged between 0.05-0.72 with a mean of 0.19. Similarly, when the SNPs were fused with sa-dFNC features of both state 1 and state 2, 5 out of 35 components found counterparts, with the remaining similarity ranging between 0.08 and 0.79 with a mean of 0.45. These observations indicated that locally optimizable SNP factors might be discovered by separately linking SNPs to each dFNC state.

## Discussion

As expected, both SNP and dFNC elements yielded by the dynamic fusion showed a wide range of state variability, while differences were also noted between the two modalities. As shown in Figure 2a, the SNP modality reached higher across-state similarity while more dFNC elements presented an intermediate level of state variability. The highly similar, or state-invariant SNP elements were in general accompanied by dFNC elements with intermediate to low similarity (Table 1), and typically highlighted a small set of top genes and connectivity pairs, as shown in Figure 2b for an example. We interpret this type of SNP-dFNC associations as localized or regional genetic variants affecting different small sets of connectivity pairs in different states.

Across four parallel fusions, a total of 53 out of 131 dFNC elements of the joint components showed significant differences between SZ cases and controls in the combined independent cohorts. These 53 joint components were considered SZ-relevant, overall evenly distributed across the range of state variability, as shown in Table 1 and Figure 2a, indicating no bias in high or low state variability for SZ relevance.

State3_Comp 7 was paired with State1_Comp 9 as an example of component showing an intermediate level of state variability, which happened to the top two SZ-discriminating components, explaining 8.18% and 5.43% of the variance in the case-control difference respectively. Both of their dFNC elements, as shown in Figure 2c and supplemental figures, highlighted a subset of the connectome which could be described as thalamus- and subthalamus/hypothalamus-hub connections to other ICNs. Thalamus plays a key role in information processing, and the abnormalities in the frontal-thalamic-cerebellar circuitry are involved in a wide range of symptoms, including SZ^52,53^. Thalamic functional dysconnectivity has also been documented in SZ^54-56^. Our findings echo previous work to confirm disrupted thalamic connections in SZ, and further reveal a thalamus-hub connectivity pattern that existed in multiple dynamic states, likely reflecting a primary composition of the connectome.

The linked genomic factors of State1_Comp9 and State3_Comp7 shared 37% of the annotated genes, both enriched for regulation of cell projection organization, a biological process that results in the assembly, arrangement of constituent parts, or disassembly of a prolongation or process extending from a cell, e.g., a flagellum or axon, as defined by Gene Ontology. This raises the question of whether the disrupted cell projection organization may contribute to functional dysconnectivity, which requires the integration of multi-scale data resources to verify and understand how it happens. State1_Comp9 was also enriched for the regulation of synaptic plasticity, as well as learning and memory. There has been the hypothesis of synaptic dysfunction in SZ^57,58^. Regional decrease in the density of postsynaptic elements has been reported for SZ in a meta-analysis of postmortem brain studies^59^. The current finding also finds support from previous work where reduced thalamic-prefrontal connectivity was associated with impaired working memory in SZ^55^. While in-depth functional annotation is beyond the scope of the current study, our observation encourages further investigations at the cellular level on how the identified genomic risk translates into functional dysconnectivity between the thalamus and other brain networks that are characteristic of SZ.

Another pair of SZ-relevant components with an intermediate level of variability included State1_Comp13 and State3_Comp5, both of which highlighted cerebellum-hub connections to other ICNs (Figure 2c), explaining 1.47% and 3.33% variances in case-control differences, respectively. Also as an important region of the frontal-thalamo-cerebellar circuitry, the role of the cerebellum in cognitive symptoms of SZ has been more and more emphasized ^60-62^. More recently, gray matter reduction in the cerebellum has been suggested to modulate static and dynamic cerebellum dysconnections with the thalamus and frontal regions in SZ^63^. Our findings of the cerebellum-hub connectivity pattern, along with the thalamus-hub components discussed above, are in high concert with the literature to emphasize the key hub roles of these two regions in SZ-related dysconnectivity. The linked genomic factors of these two pairs shared 31% of the annotated genes, although enriched in different biological processes. It is noted that in addition to synapse organization, whose relevance to SZ has been discussed above, State1_Comp13 was enriched for regulation of neuron projection development, which also plays an important role in the neuropathology of SZ. Neuron projection may refer to any process extending from a neural cell, such as axons or dendrites, that are involved in communication between neurons and hence functional connectivity. Again, despite that these enriched biological processes may be reasonably linked to the dFNC features, further verification is warranted and investigations are needed to characterize whether genomic risk affects the brain through regulating gene expression or other mechanisms.

State1_Comp30 and State2_Comp32 are examples of state-invariant localized SNP elements, which showed a high correlation of 1 (Table 1). Their SNP elements highlighted the same annotated genes: *MFHAS1, MSRA, and SGK223* (Table S1). The linked dFNC elements reflected connections between visual, cognitive control, and default mode networks (Figure 2b). It is noted that *SGK223* (also known as *PRAG1* or pragmin) induces Rho-dependent cell contraction in neuronal cells and may regulate neurite outgrowth^64^. It remains to be elucidated whether any additional pathways may bridge between these genes and connectivity features.

Last but not least, we have identified joint components located at the high end of state variability, such as State1_Comp4 and State1_Comp6 (Figure 2d), which could likely be elicited only upon dynamic fusion. And we interpret them as state-variant manifolds. The connectogram of State1_Comp4 presents an insula- hub connectivity pattern while its SNP element was enriched for regulation of cell projection organization. Note that State1_Comp4 and State1_Comp13 were enriched for the same biological process even though they were not correlated with one another. This observation indicated that the State1_Comp4 SNPs might complement State1_Comp13 to provide a more complete picture of the genomic factor that confers risk to SZ by affecting cell projection organization. This also presents the benefit of dynamic fusion to augment the SNP factors to across-state manifolds that may provide additional insights into the underlying biology. There has been plenty of evidence for the insula pathology in SZ^65^, including functional dysconnectivity that showed correlation with cognitive function and negative symptoms^66^, as well as implicated in the SZ-related aberrant salience network connectivity during information processing^67^. State1_Comp6 describes an association between MTG-hub connections and a genomic factor enriched for the regulation of synaptic plasticity. Both structural abnormalities in MTG and MTG dysconnectivity have been linked to auditory verbal hallucinations in SZ^68,69^. The SNPs and genes identified in State1_Comp6 served as a complement to those of State1_Comp9 and State1_Comp13 to provide a more complete view of the synapse-related genomic risk for SZ. Given that synaptic plasticity has been suggested to underlie brain connectivity^70^, it is reasonable that several of our identified SNP elements were enriched for this biological process. While these SNP elements did not significantly overlap, it is intriguing to investigate whether the element-specific genes show tissue or regional specificity to impact different connectivity pairs.

This work presents a novel framework and some initial evidence to showcase how dynamic SNP-dFNC fusion expands the SNP modality for biologically meaningful state-variant manifolds that offer additional insights into SZ. The results should be interpreted in light of the following limitations. First, a more sophisticated adaptive normalization may be needed to better compensate for the difference in SNP and dFNC data structure so that more balanced data decomposition can be achieved to allow both modalities contributing equally to the components^71^. Second, fuzzy clustering or meta-state approaches may be employed to allow each dFNC time window to have a probabilistic rather than categorical state membership, or represent a linear combination of dynamic states rather than being assigned to one state^28,72^. Third, the current SNP-dFNC dynamic fusion utilized the large population cohort of UKB for discovery. Although we validated the findings in the combined SZ cohort, it remains a question of whether a dynamic fusion directly applied to SZ cohorts may yield additional insights. Fourth, we did not strictly calibrate the window lengths of UKB and SZ data in the dFNC analysis. With that said, the dynamic fusion framework appears to robustly identify meaningful components that could be validated for disease relevance.

In conclusion, this work aims to present a novel dynamic fusion framework to identify state-variant manifolds of SNP-dFNC associations for additional biological insights. Overall, results support the hypothesis that dynamic fusion yields meaningful state-variant manifolds. Particularly, the SNP manifolds uniquely elicited by the dynamic fusion complement each other in terms of biological interpretation. This dynamic fusion framework thus provides a unique lens to elicit unique SNP correlates otherwise unseen, promising additional insights into how genetic risk impacts disease-related dysconnectivity.

## Authors’ Contributions

J.C. and V.D.C. designed the study. J.C. conducted the analyses and drafted the paper with feedback from all authors. Z.F. contributed to the data processing. All authors contributed to the writing process and have approved the final manuscript.

## Data Availability

All data produced in the present study are available upon reasonable request to the authors.

## Acknowledgments

This project was funded by the National Institutes of Health (P20GM103472, R01EB005846, 1R01EB006841, R01MH106655, 5R01MH094524) and the National Science Foundation (1539067).

