## Supplemental Figure 1 for "Dynamic fusion of genomics and functional network connectivity in UK biobank reveals schizophrenia-related SNP manifolds"

Supplemental Figure 1: Manhattan plots of the SNP elements of the SZ-related components.

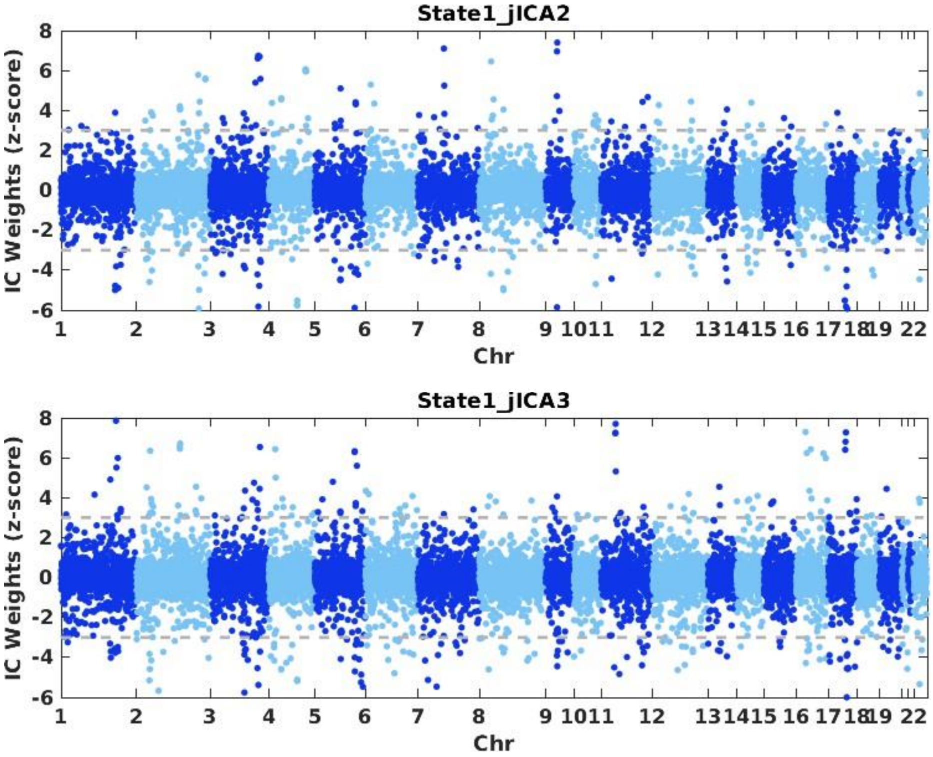

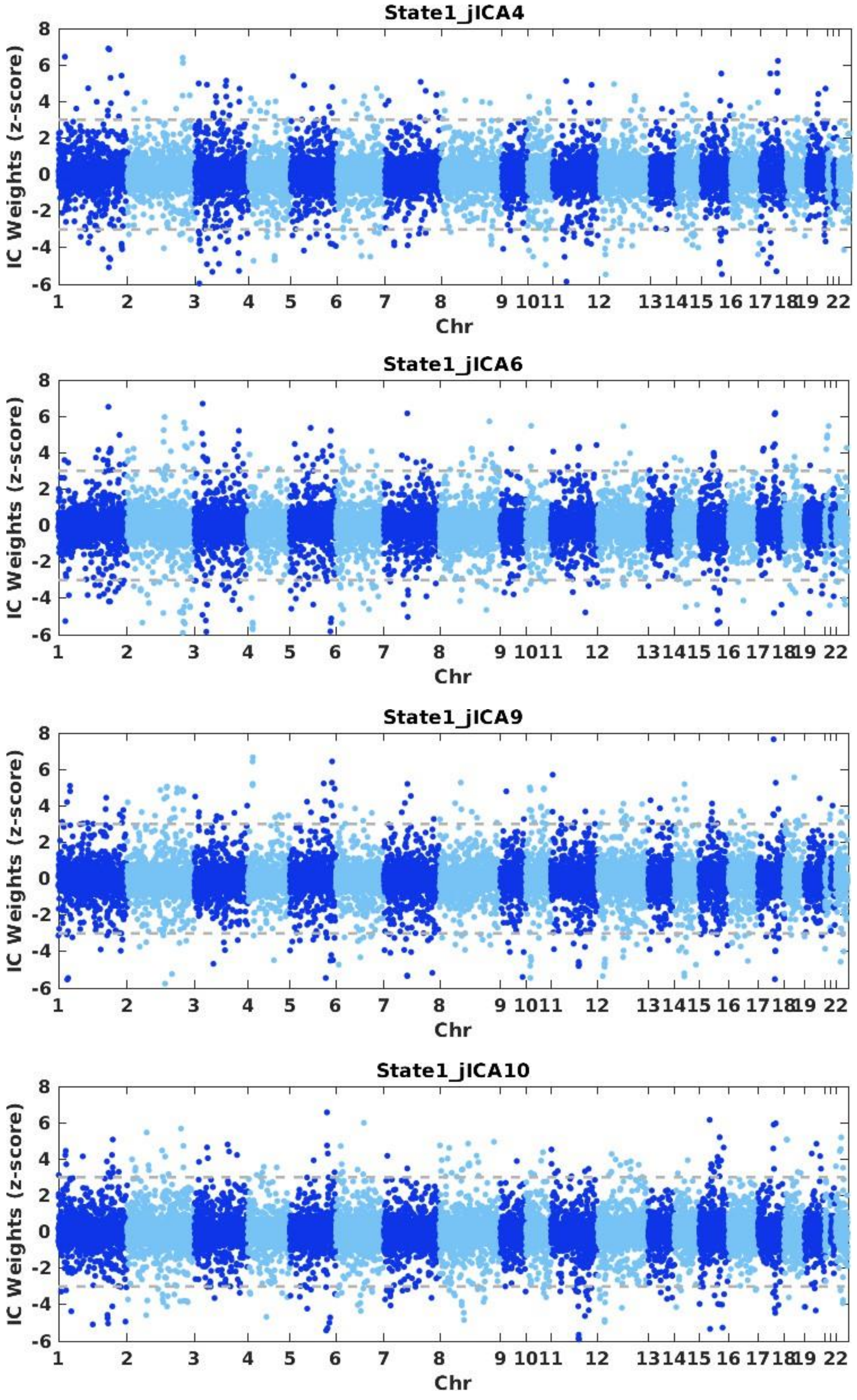
State1_jICA4
IC Weights (z-score)
Chr
State1_jICA6
IC Weights (z-score)
Chr
State1_jICA9
IC Weights (z-score)
Chr
State1_jICA10
IC Weights (z-score)
Chr

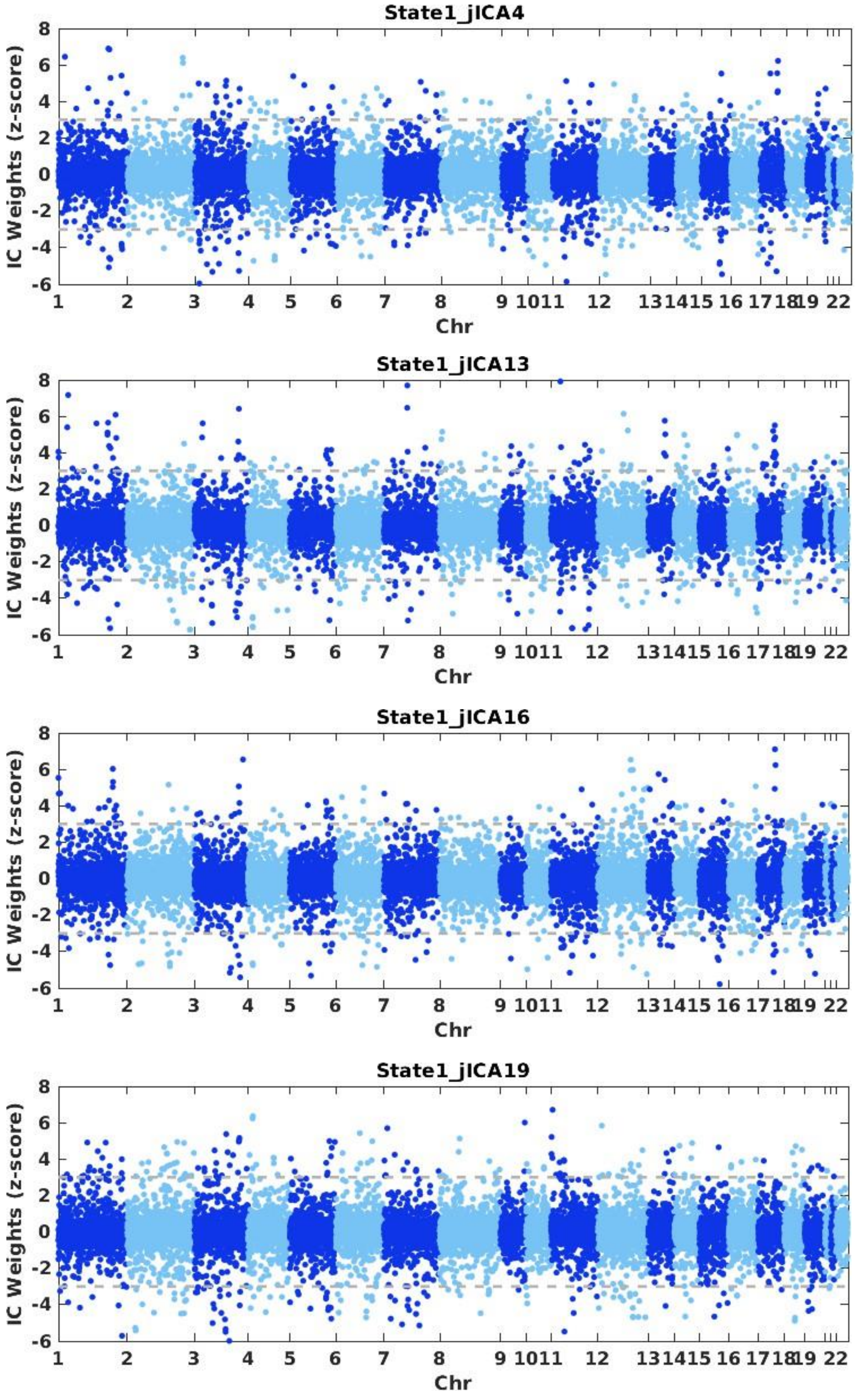
State1_jICA4
State1_jICA13
State1_jICA16
State1_jICA19
IC Weights (z-score)
Chr
8
6
4
2
0
-2
-4
-6
1
2
3
4
5
6
7
8
9 1011
12
131415 16 171819 22

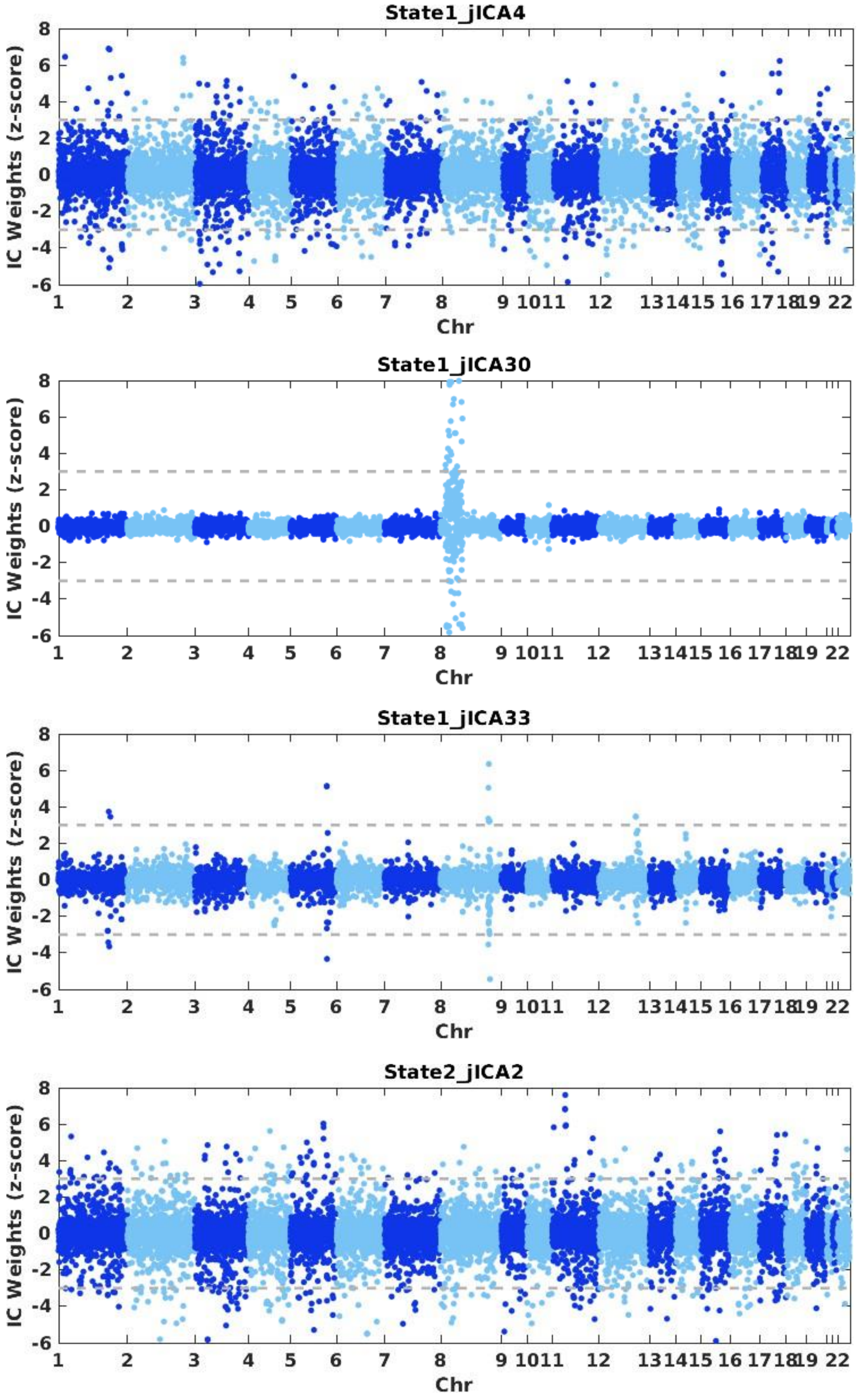
State1_jICA4
State1_jICA30
State1_jICA33
State2_jICA2
IC Weights (z-score)
8
6
4
2
0
-2
-4
-6
1 2 3 4 5 6 7 8 9 10 11 12 13 14 15 16 17 18 19 22
Chr

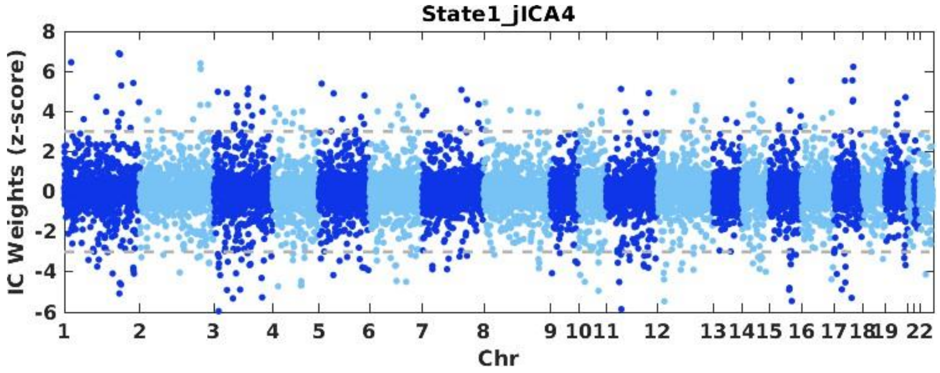
State1_jICA4
IC Weights (z-score)
8 6 4 2 0 -2 -4 -6
1 2 3 4 5 6 7 8 9 1011 12 131415 16 1718 19 22
Chr

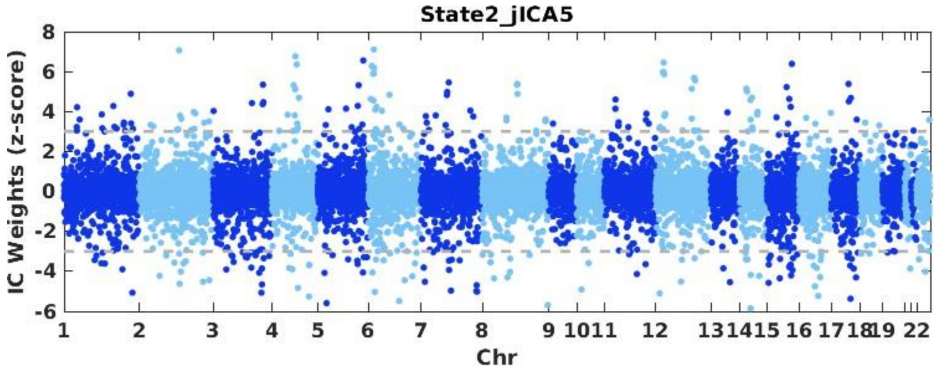
State2_jICA5
IC Weights (z-score)
8 6 4 2 0 -2 -4 -6
1 2 3 4 5 6 7 8 9 1011 12 131415 16 1718 19 22
Chr

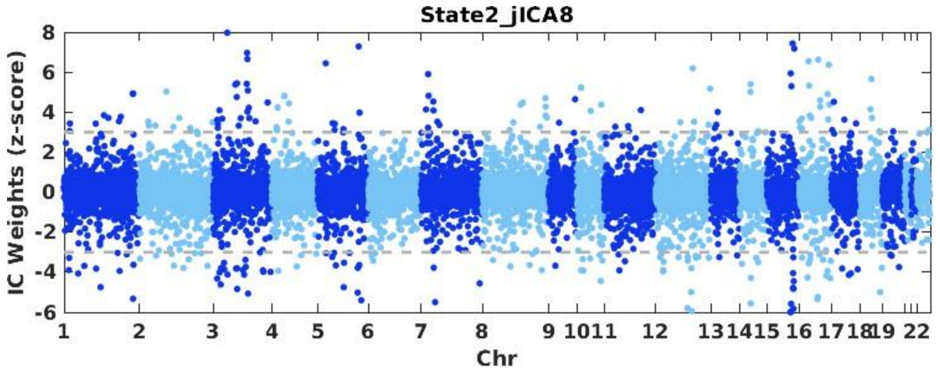
State2_jICA8
IC Weights (z-score)
8 6 4 2 0 -2 -4 -6
1 2 3 4 5 6 7 8 9 1011 12 131415 16 1718 19 22
Chr

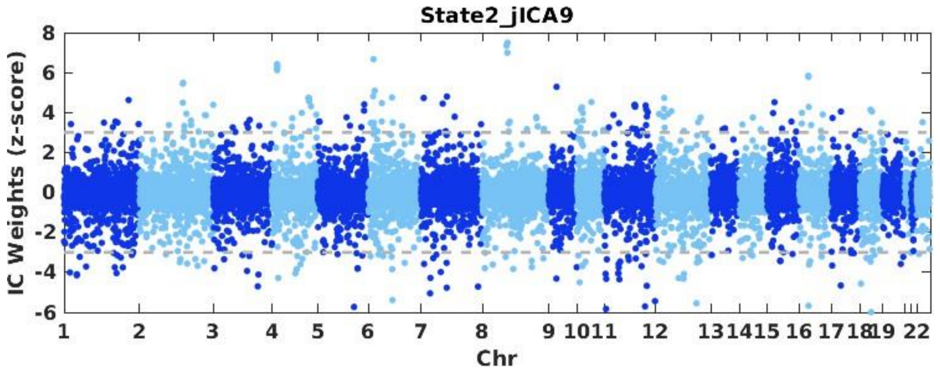
State2_jICA9
IC Weights (z-score)
8 6 4 2 0 -2 -4 -6
1 2 3 4 5 6 7 8 9 1011 12 131415 16 1718 19 22
Chr

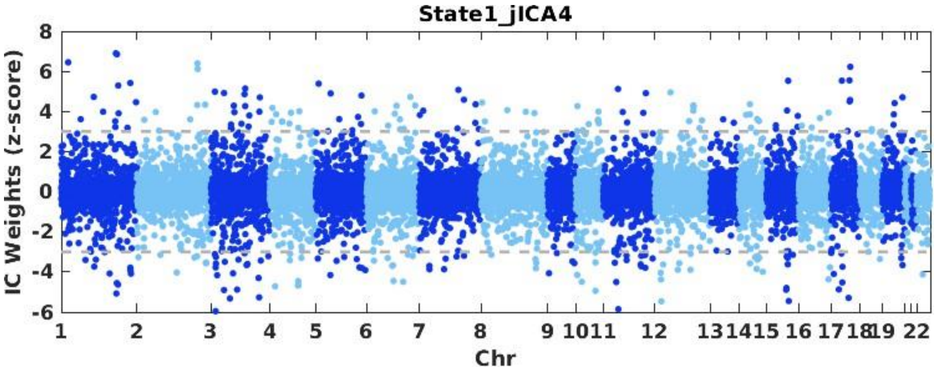
State1_jICA4
IC Weights (z-score)
8
6
4
2
0
-2
-4
-6
1 2 3 4 5 6 7 8 9 1011 12 131415 16 171819 22
Chr

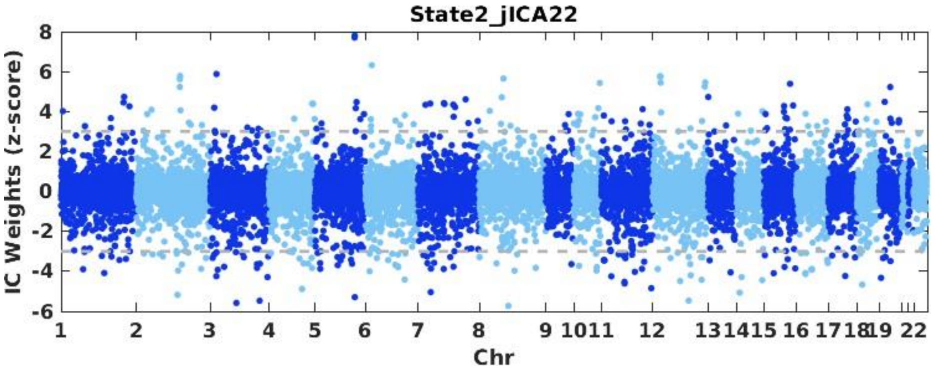
State2_jICA22
IC Weights (z-score)
8
6
4
2
0
-2
-4
-6
1 2 3 4 5 6 7 8 9 1011 12 131415 16 171819 22
Chr

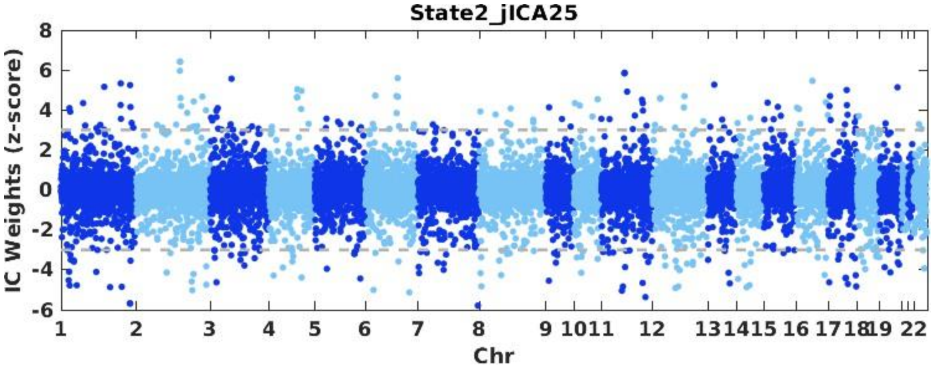
State2_jICA25
IC Weights (z-score)
8
6
4
2
0
-2
-4
-6
1 2 3 4 5 6 7 8 9 1011 12 131415 16 171819 22
Chr

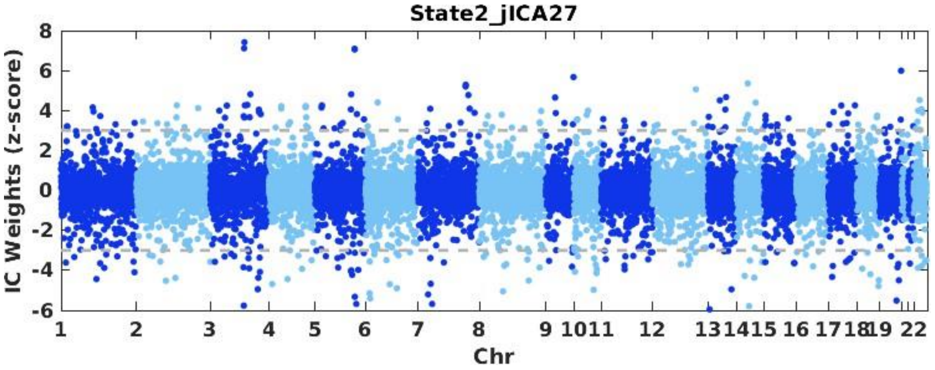
State2_jICA27
IC Weights (z-score)
8
6
4
2
0
-2
-4
-6
1 2 3 4 5 6 7 8 9 1011 12 131415 16 171819 22
Chr

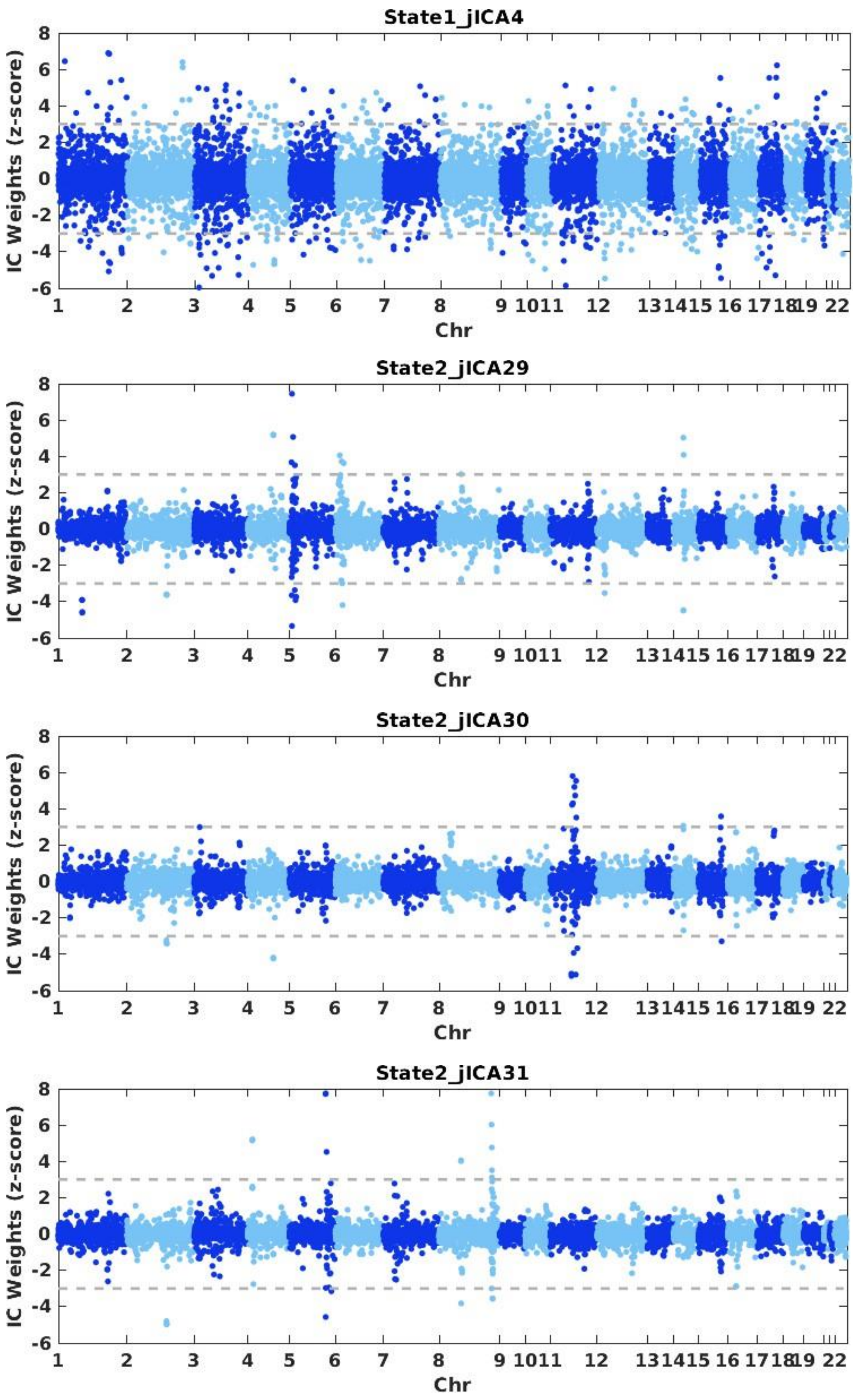
State1_jICA4
State2_jICA29
State2_jICA30
State2_jICA31
IC Weights (z-score)
Chr

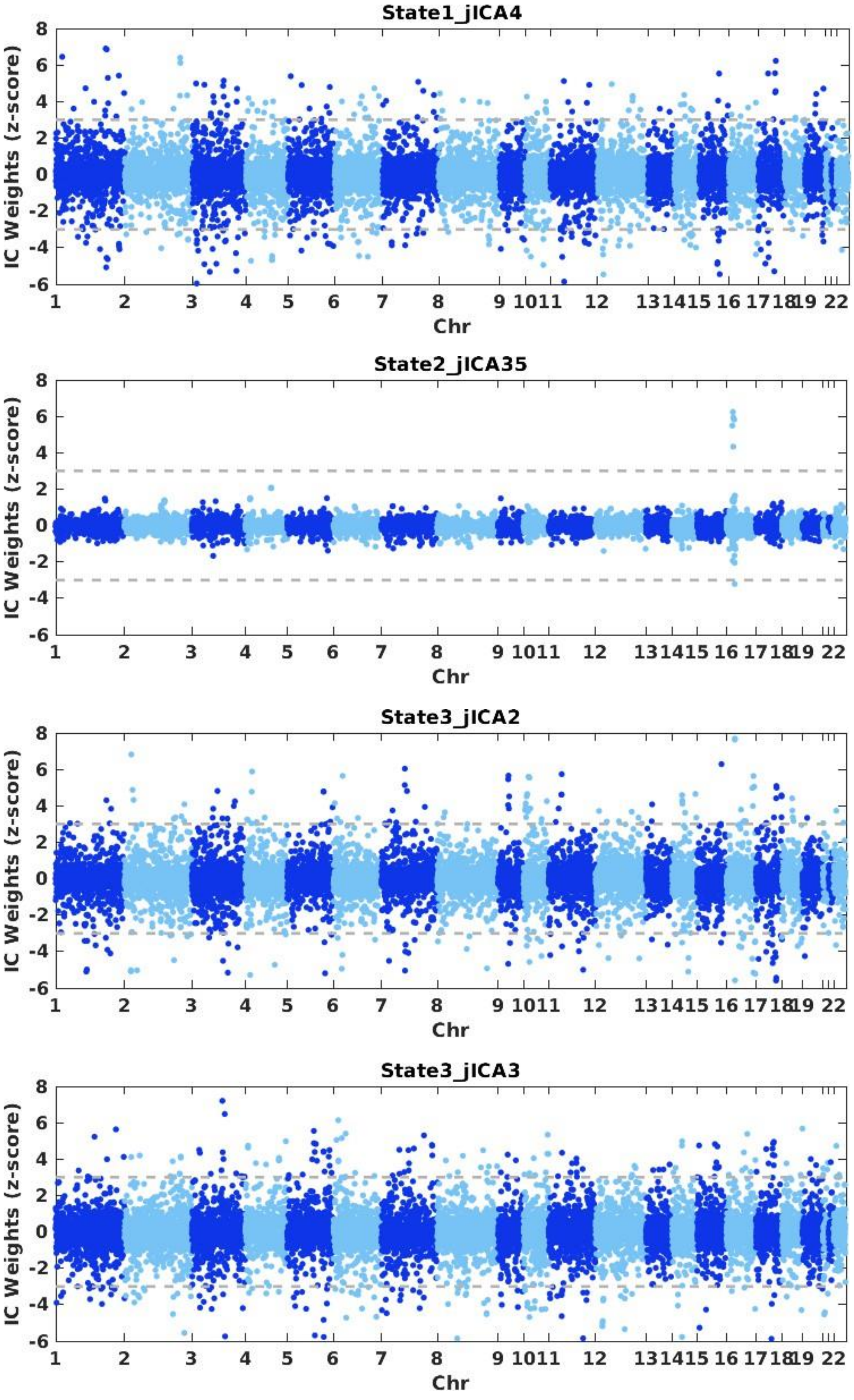
State1_jICA4
State2_jICA35
State3_jICA2
State3_jICA3
IC Weights (z-score)
Chr
1 2 3 4 5 6 7 8 9 1011 12 131415 16 1718 19 22

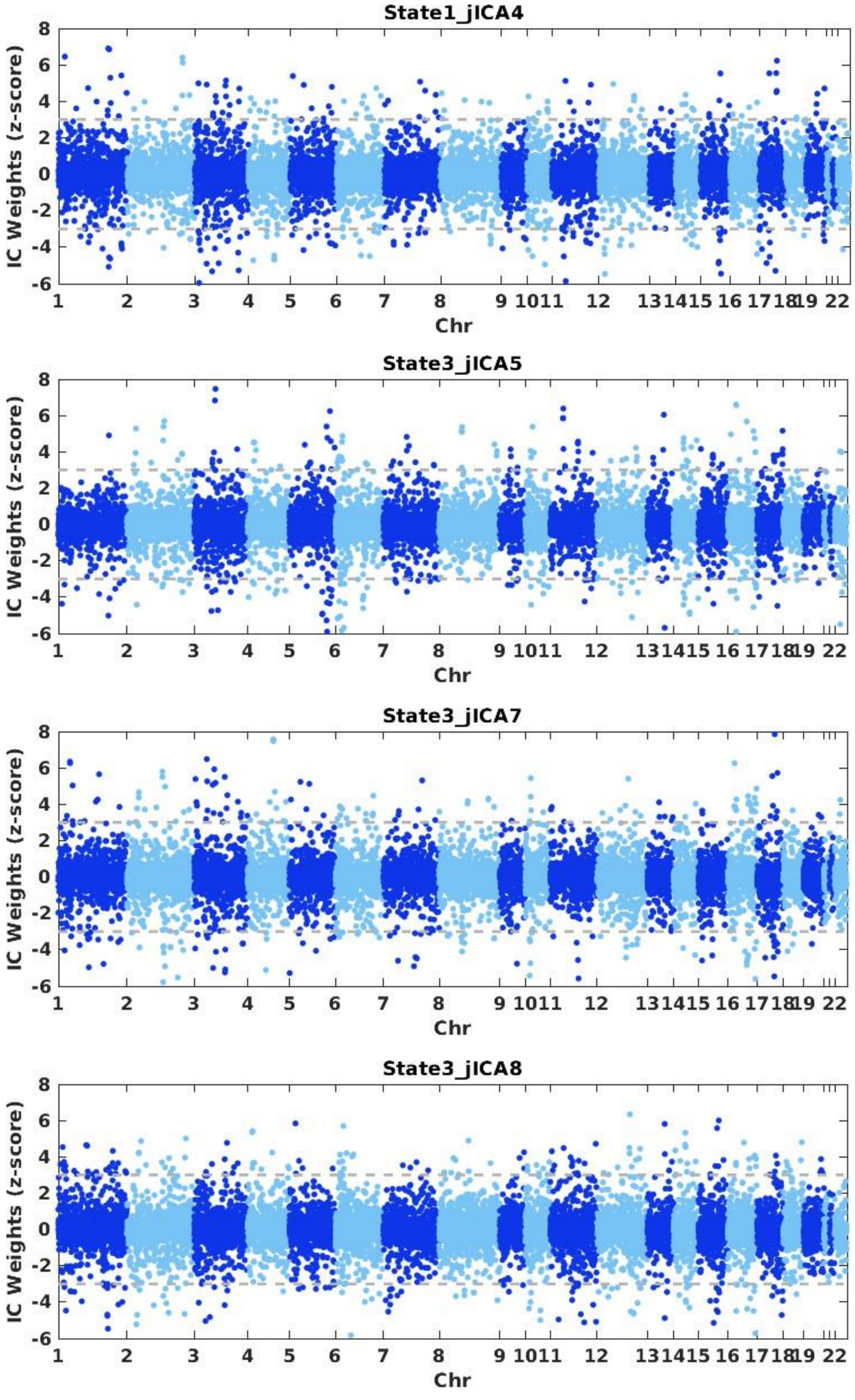
State1_jICA4
IC Weights (z-score)
Chr
State3_jICA5
IC Weights (z-score)
Chr
State3_jICA7
IC Weights (z-score)
Chr
State3_jICA8
IC Weights (z-score)
Chr

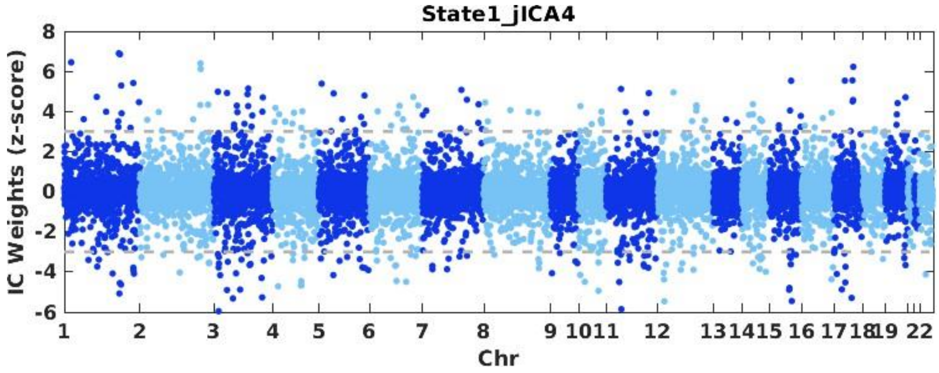
State1_jICA4
IC Weights (z-score)
8 6 4 2 0 -2 -4 -6
1 2 3 4 5 6 7 8 9 1011 12 131415 16 171819 22
Chr

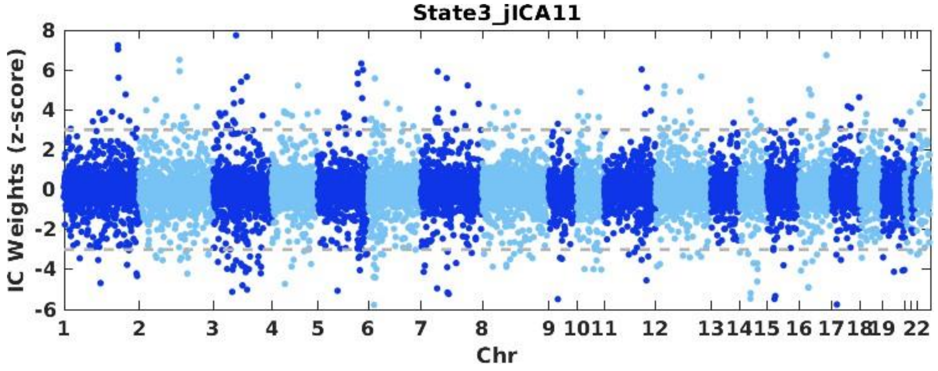
State3_jICA11
IC Weights (z-score)
8 6 4 2 0 -2 -4 -6
1 2 3 4 5 6 7 8 9 1011 12 131415 16 171819 22
Chr

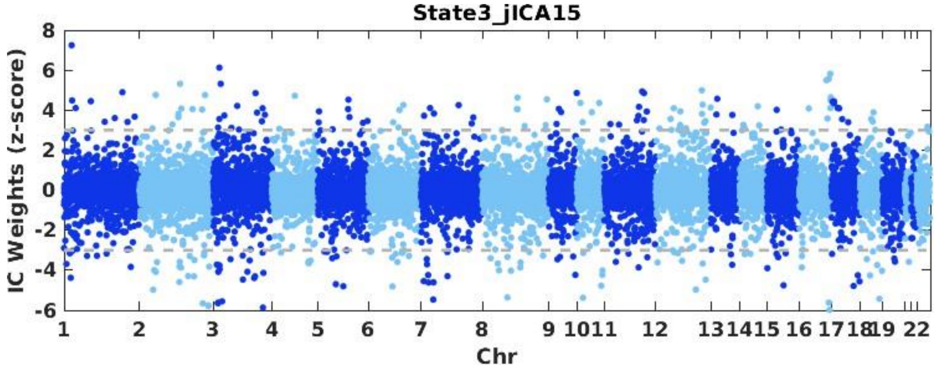
State3_jICA15
IC Weights (z-score)
8 6 4 2 0 -2 -4 -6
1 2 3 4 5 6 7 8 9 1011 12 131415 16 171819 22
Chr

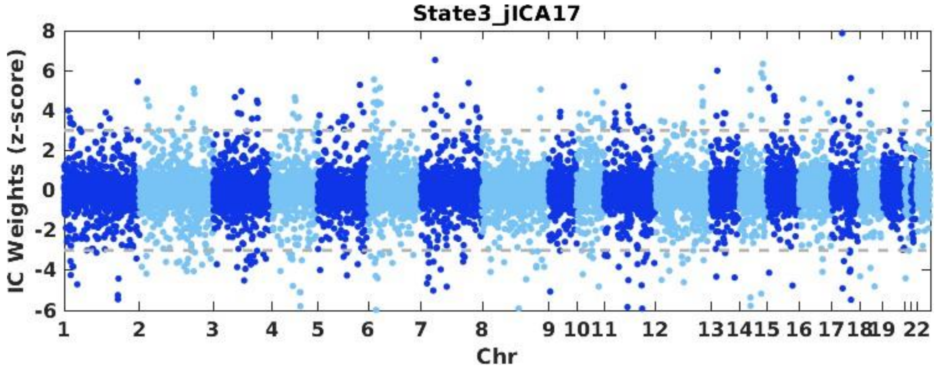
State3_jICA17
IC Weights (z-score)
8 6 4 2 0 -2 -4 -6
1 2 3 4 5 6 7 8 9 1011 12 131415 16 171819 22
Chr

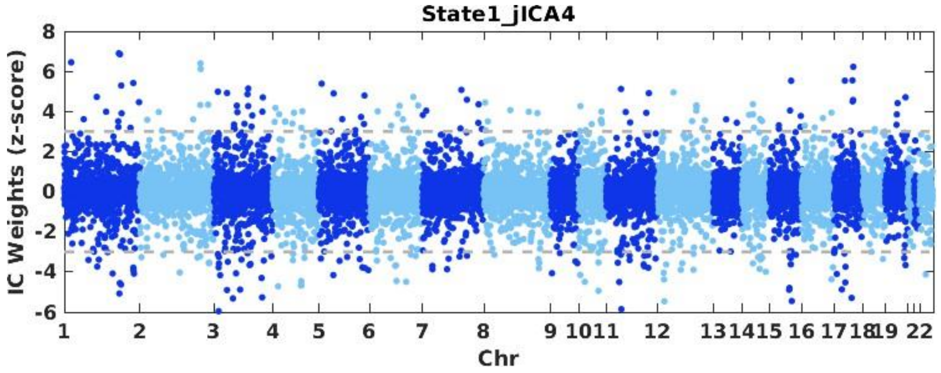
State1_jICA4
IC Weights (z-score)
Chr

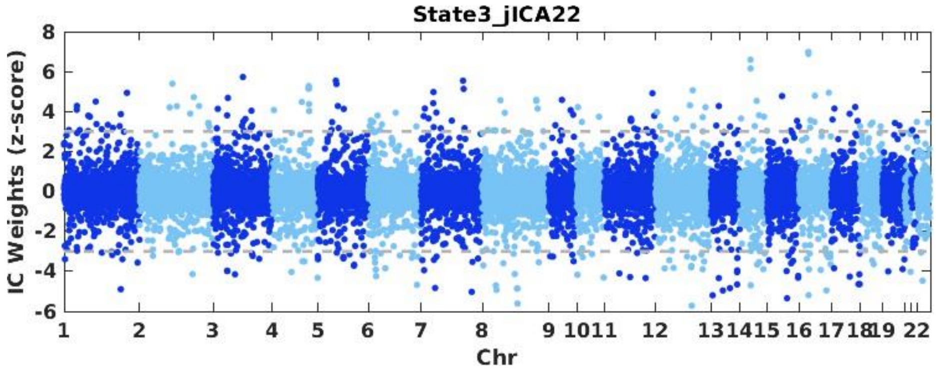
State3_jICA22
IC Weights (z-score)
Chr

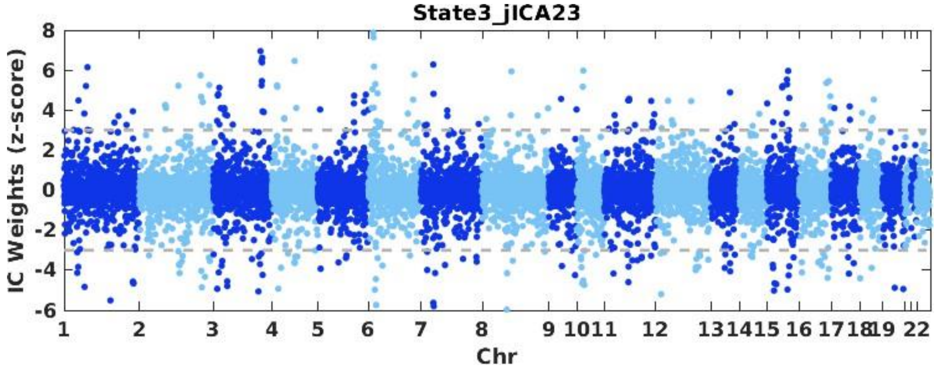
State3_jICA23
IC Weights (z-score)
Chr

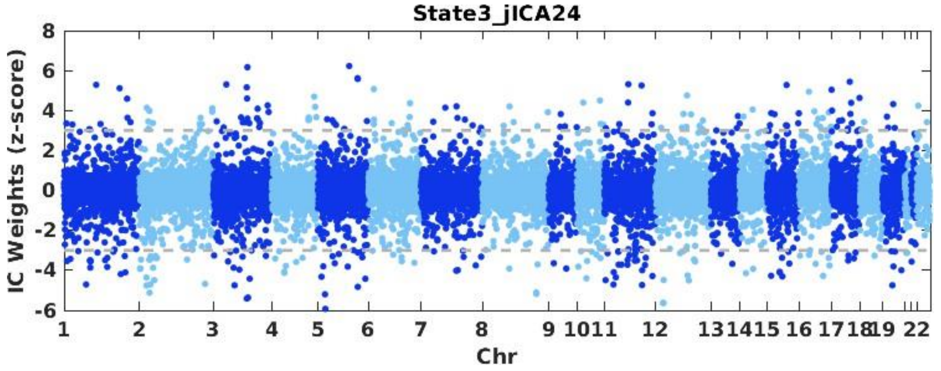
State3_jICA24
IC Weights (z-score)
Chr

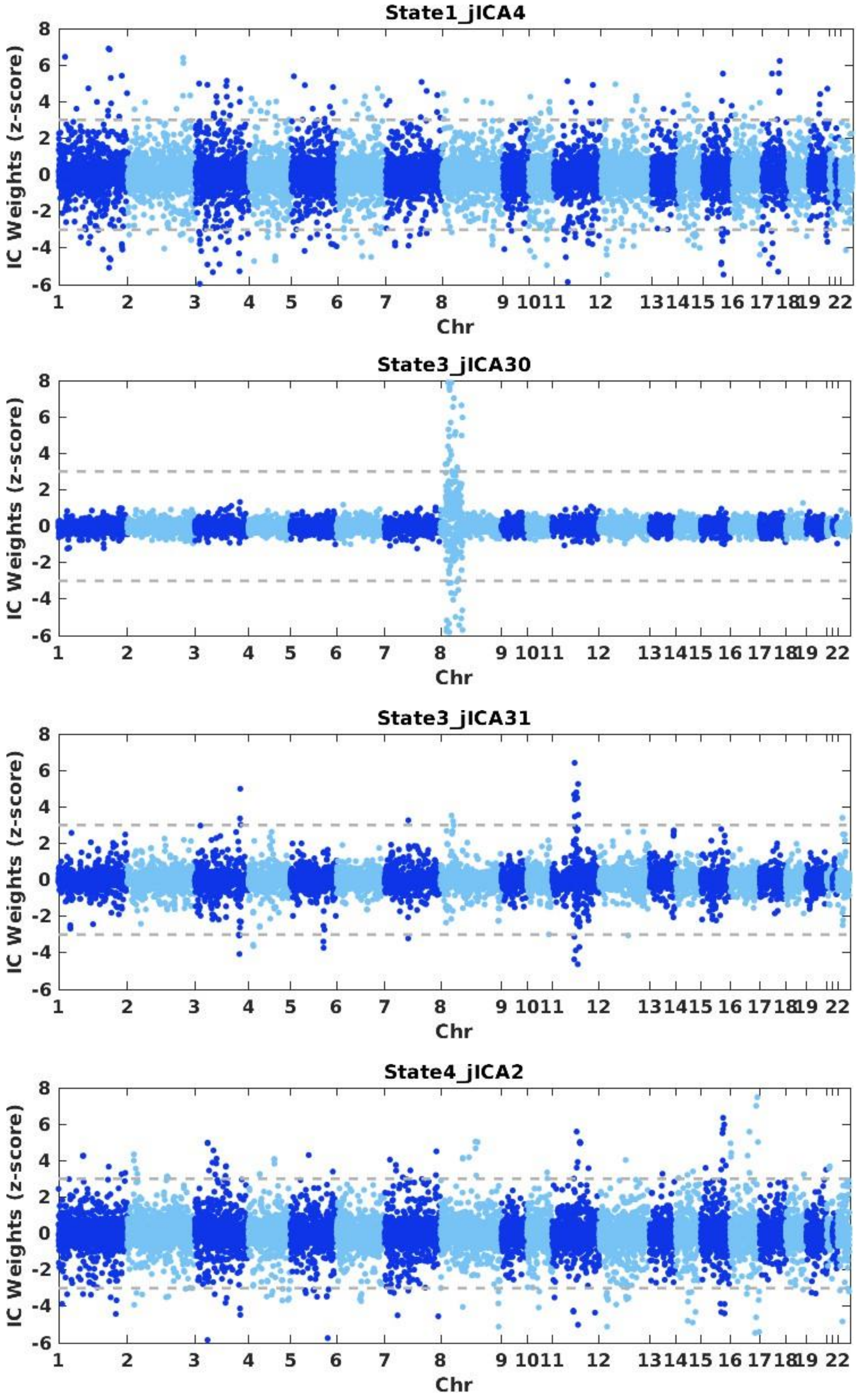
State1_jICA4
State3_jICA30
State3_jICA31
State4_jICA2
IC Weights (z-score)
Chr
8
6
4
2
0
-2
-4
-6
1 2 3 4 5 6 7 8 9 10 11 12 13 14 15 16 17 18 19 22

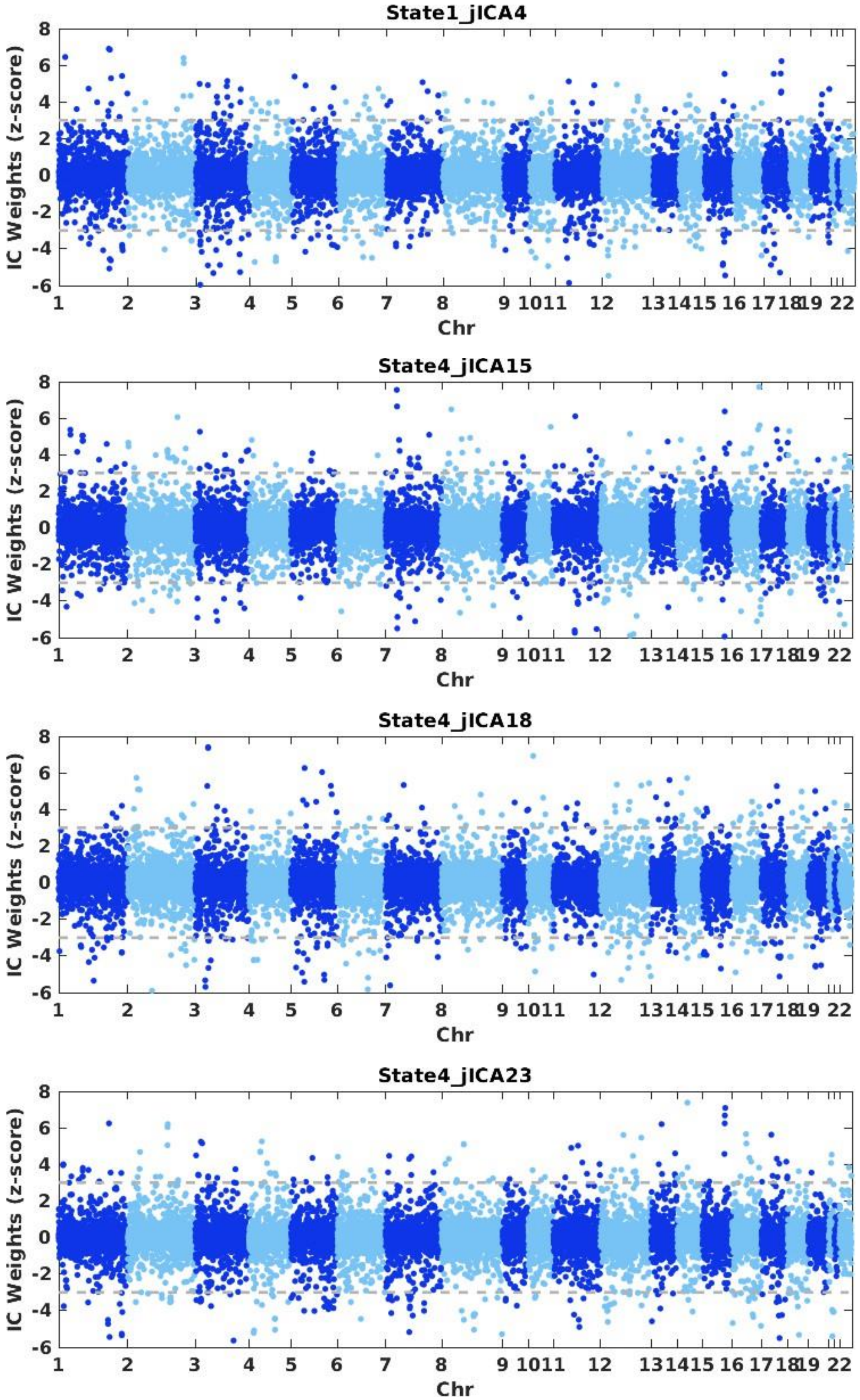
State1_jICA4
State4_jICA15
State4_jICA18
State4_jICA23
IC Weights (z-score)
Chr
1 2 3 4 5 6 7 8 9 10 11 12 13 14 15 16 17 18 19 22
8 6 4 2 0 -2 -4 -6

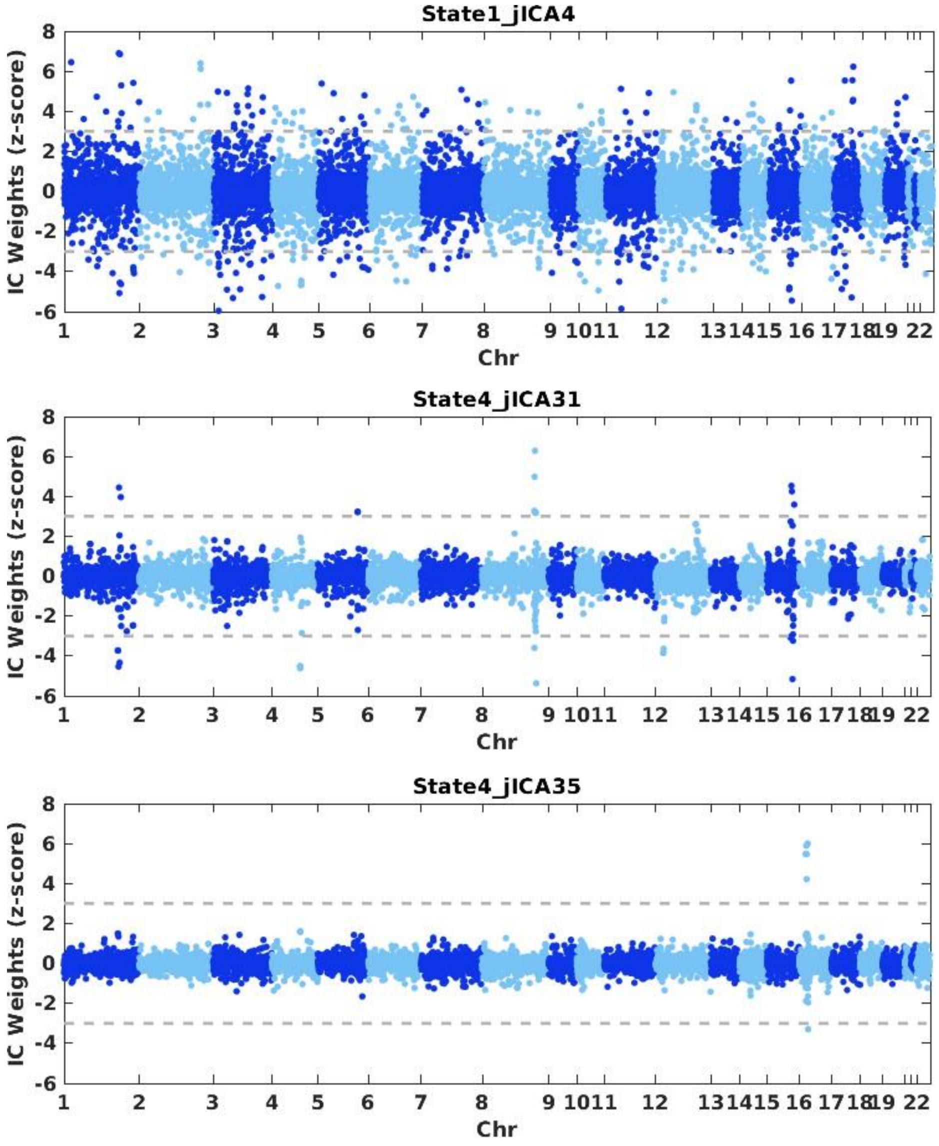


*Published with MATLAB® R2020b*
