## Supplemental Figure 2 for "Dynamic fusion of genomics and functional network connectivity in UK biobank reveals schizophrenia-related SNP manifolds"

Supplemental Figure 2: Connectogram plots of the dFNC elements of the SZ-related components.

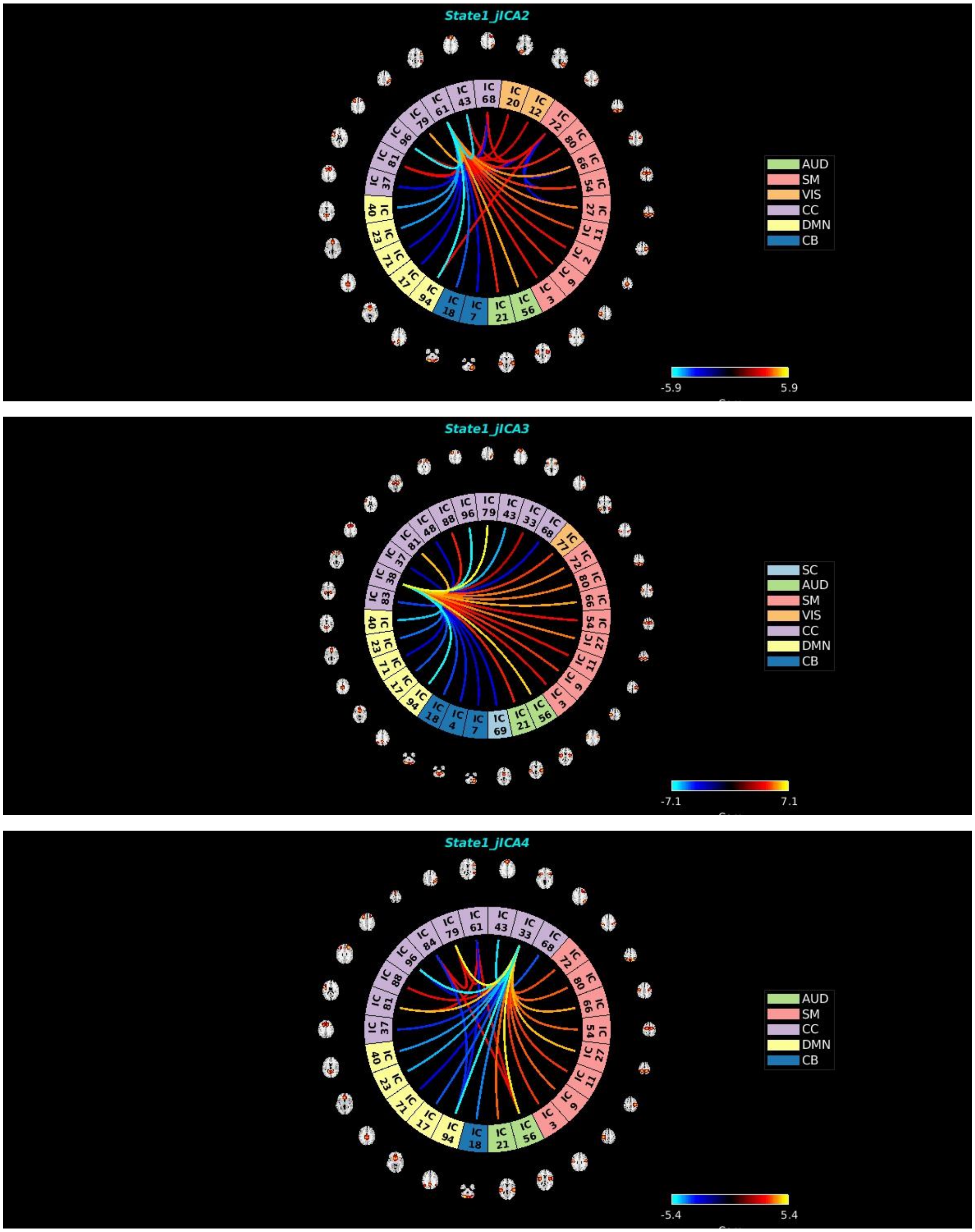

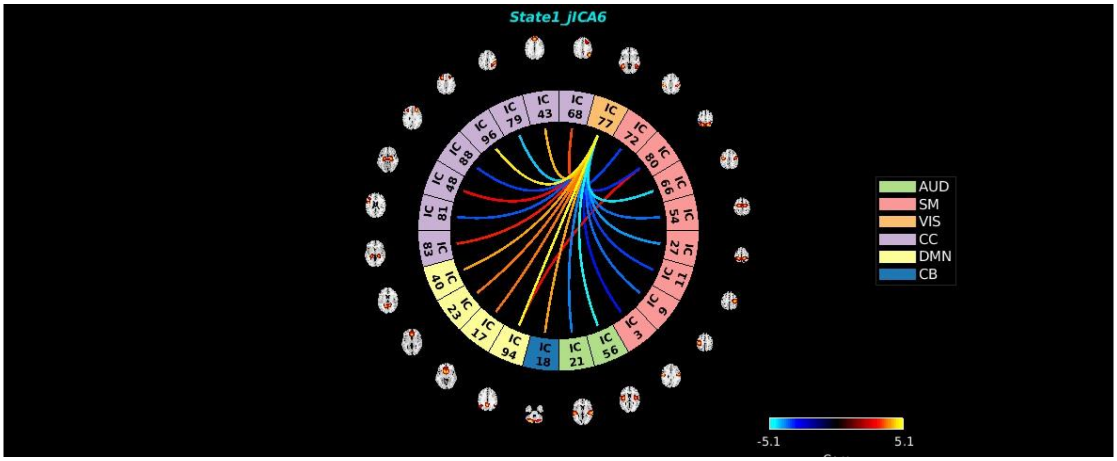
State1_jICA6
AUD
SM
VIS
CC
DMN
CB
-5.1
5.1

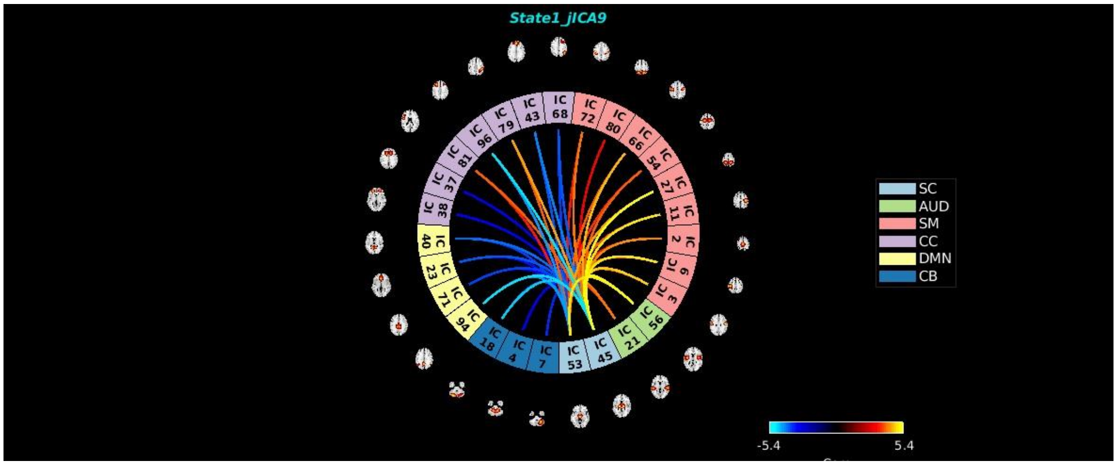
State1_jICA9
SC
AUD
SM
CC
DMN
CB
-5.4
5.4

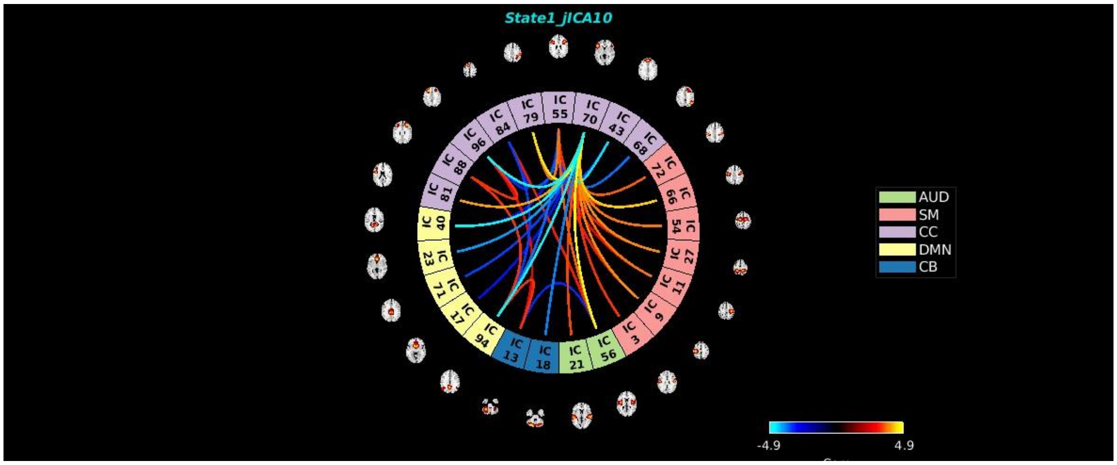
State1_jICA10
AUD
SM
CC
DMN
CB
-4.9
4.9

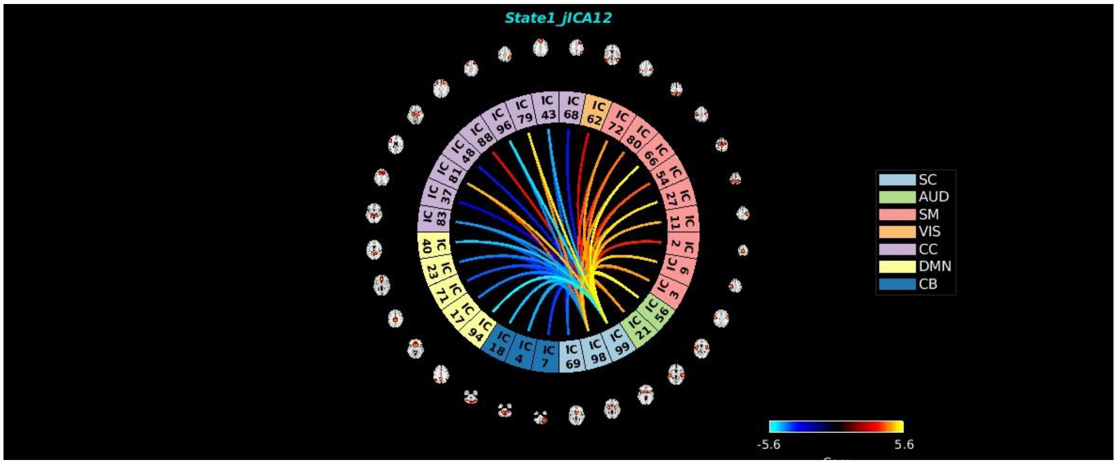
State1_jICA12
SC
AUD
SM
VIS
CC
DMN
CB
-5.6
5.6

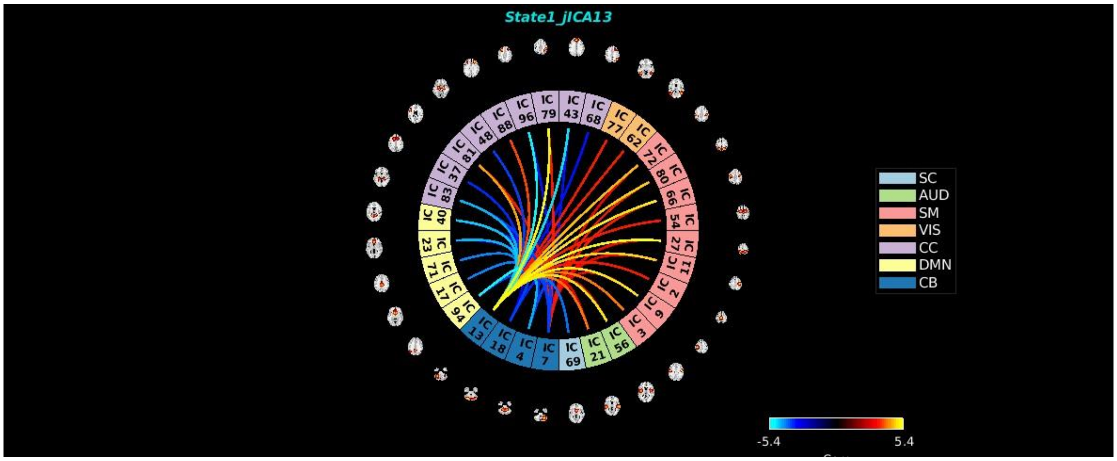
State1_jICA13
SC
AUD
SM
VIS
CC
DMN
CB
-5.4
5.4

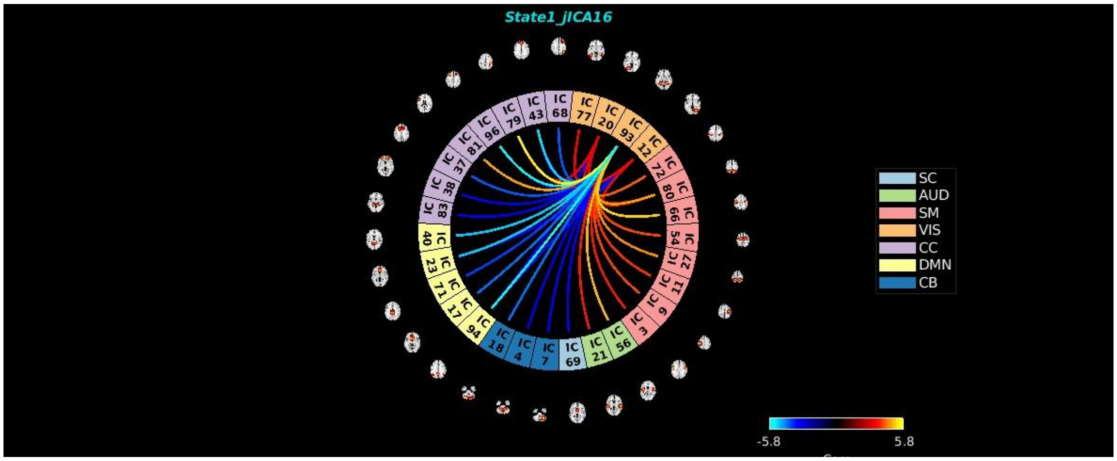
State1_jICA16
SC
AUD
SM
VIS
CC
DMN
CB
-5.8
5.8

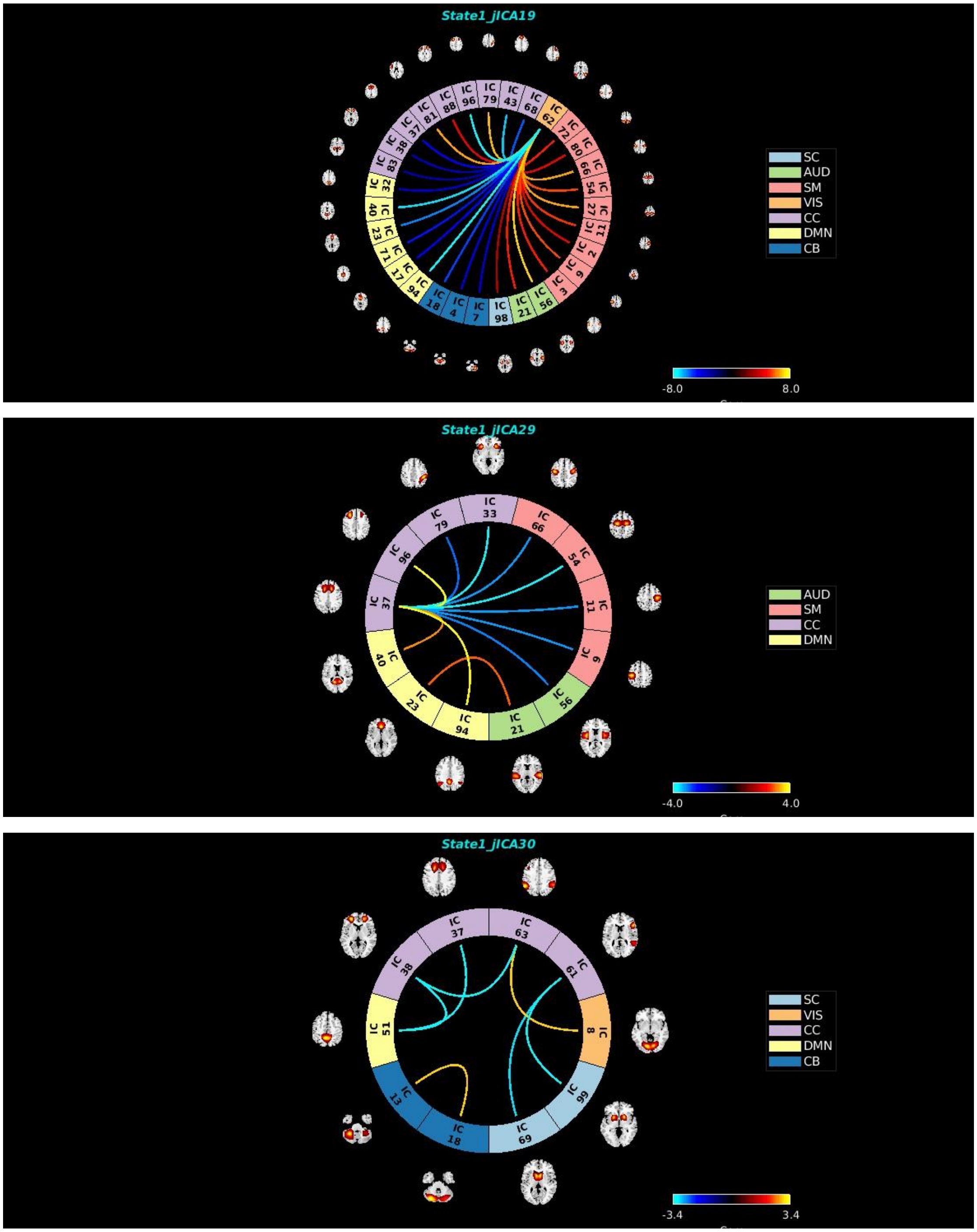
State1_jICA19
SC
AUD
SM
VIS
CC
DMN
CB
-8.0
8.0
State1_jICA29
AUD
SM
CC
DMN
-4.0
4.0
State1_jICA30
SC
VIS
CC
DMN
CB
-3.4
3.4

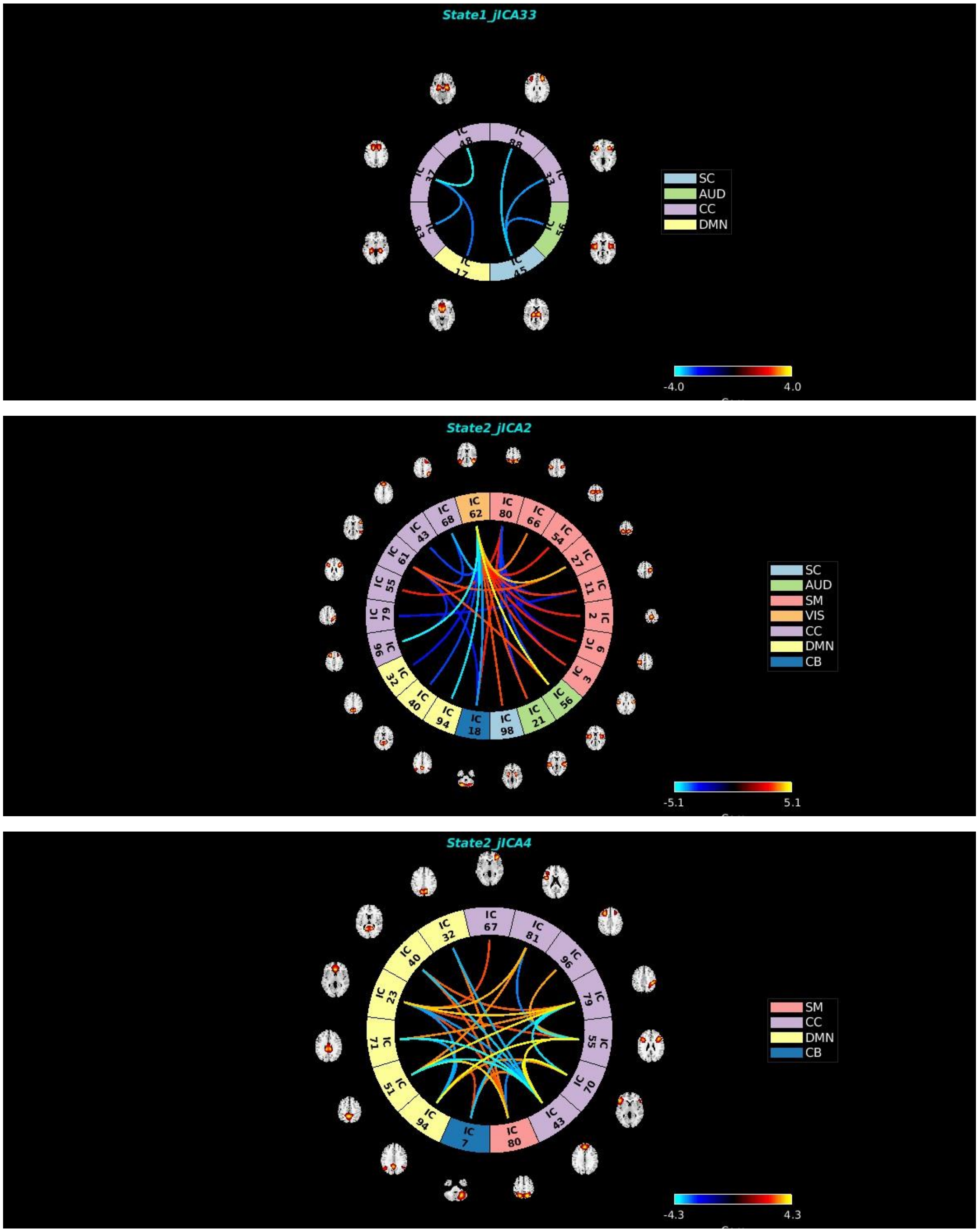
State1_jICA33
SC
AUD
CC
DMN
-4.0
4.0
State2_jICA2
SC
AUD
SM
VIS
CC
DMN
CB
-5.1
5.1
State2_jICA4
SM
CC
DMN
CB
-4.3
4.3

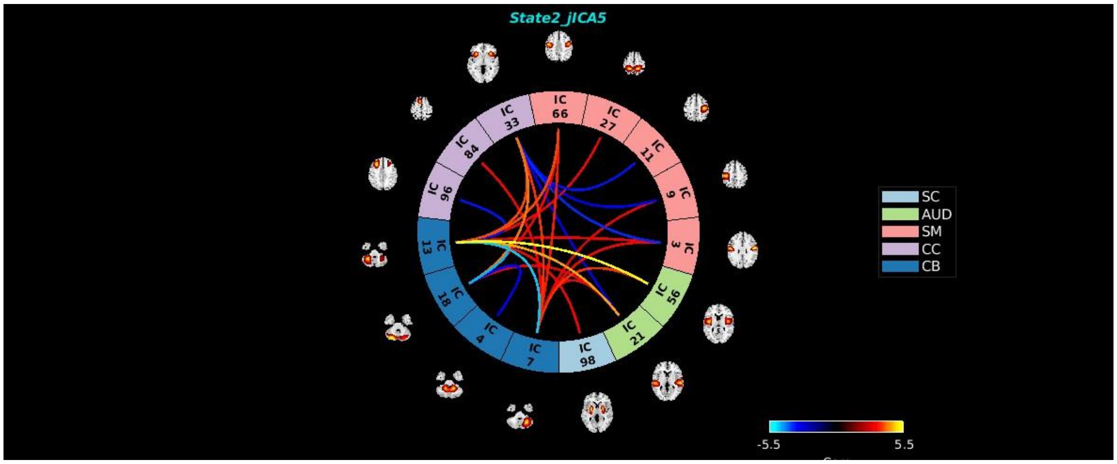
State2_jICA5
IC 66
IC 27
IC 11
IC 9
IC 3
IC 56
IC 21
IC 98
IC 7
IC 4
IC 18
IC 13
IC 96
IC 84
IC 33
SC
AUD
SM
CC
CB
-5.5
5.5

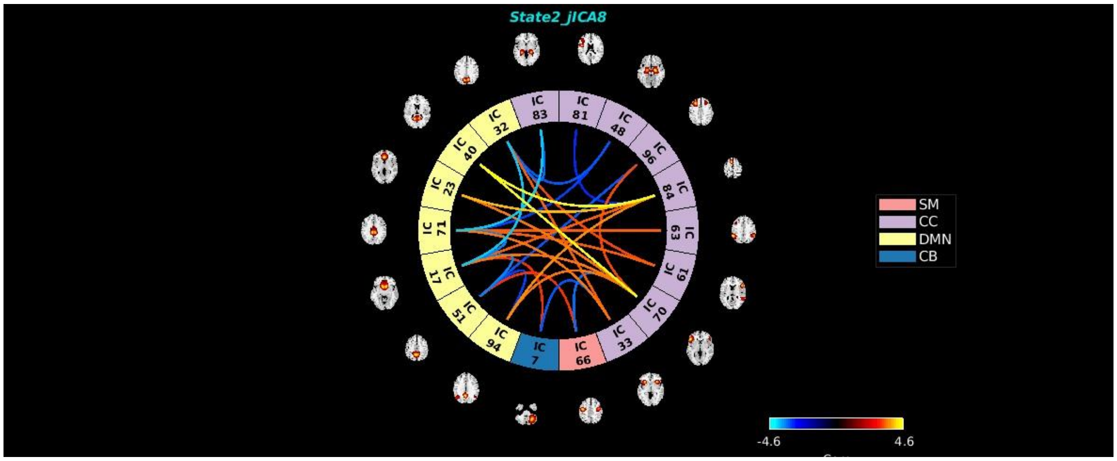
State2_jICA8
IC 83
IC 81
IC 48
IC 96
IC 84
IC 63
IC 19
IC 70
IC 33
IC 66
IC 7
IC 94
IC 51
IC 17
IC 71
IC 23
IC 40
IC 32
SM
CC
DMN
CB
-4.6
4.6

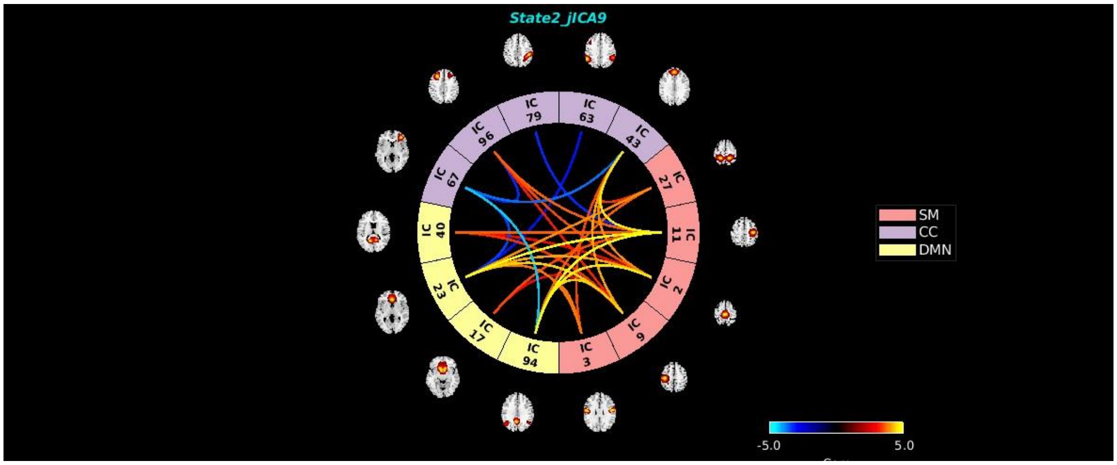
State2_jICA9
IC 79
IC 63
IC 43
IC 27
IC 11
IC 2
IC 9
IC 3
IC 94
IC 17
IC 23
IC 40
IC 67
IC 96
SM
CC
DMN
-5.0
5.0

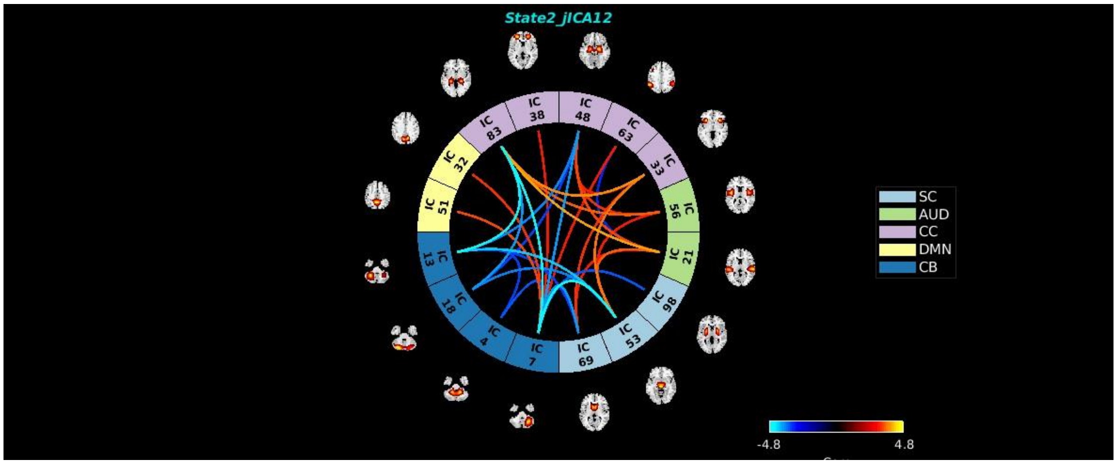
State2_jICA12
IC 38
IC 48
IC 63
IC 33
IC 56
IC 21
IC 98
IC 53
IC 69
IC 7
IC 4
IC 18
IC 13
IC 51
IC 32
IC 83
SC
AUD
CC
DMN
CB
-4.8
4.8

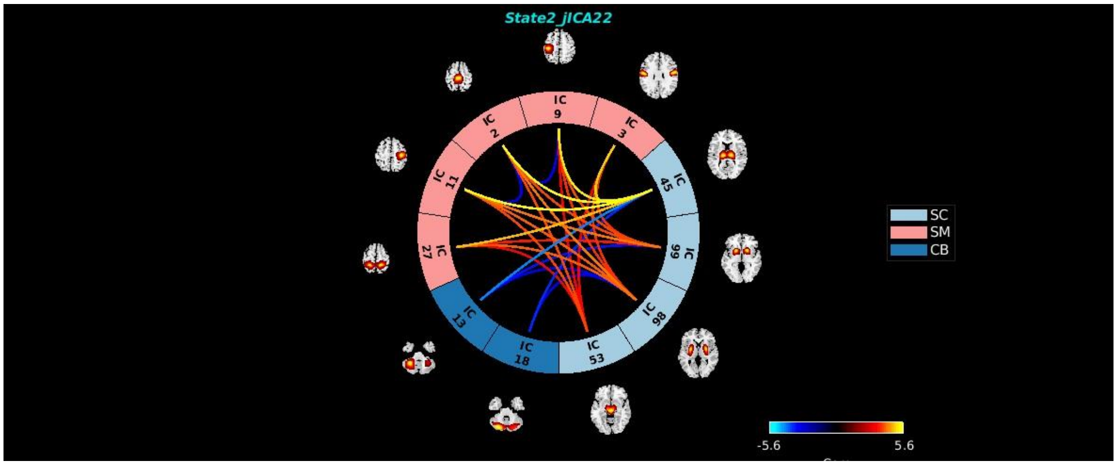
State2_jICA22
IC 9
IC 3
IC 45
IC 66
IC 98
IC 53
IC 18
IC 13
IC 27
IC 11
IC 2
SC
SM
CB
-5.6
5.6

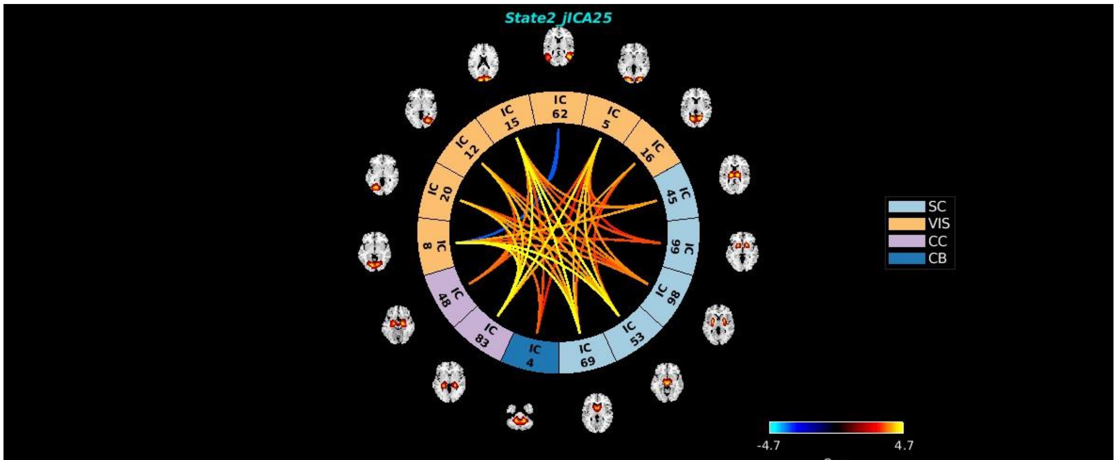
State2_jICA25
IC 62
IC 5
IC 16
IC 45
IC 66
IC 98
IC 53
IC 69
IC 4
IC 83
IC 48
IC 8
IC 20
IC 12
IC 15
SC
VIS
CC
CB
-4.7
4.7

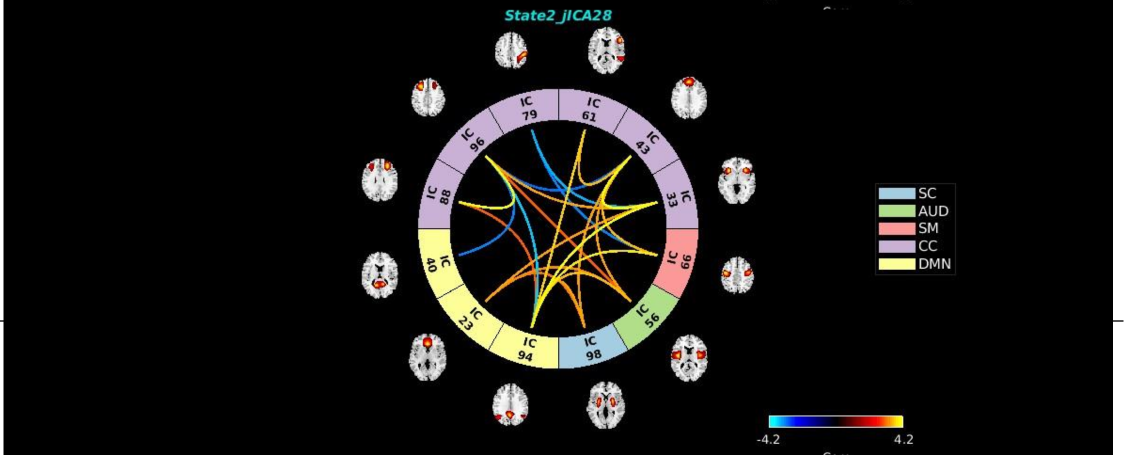
State2_jICA28
IC 79
IC 61
IC 43
IC 33
IC 69
IC 56
IC 98
IC 94
IC 23
IC 40
IC 88
IC 96
SC
AUD
SM
CC
DMN
-4.2
4.2

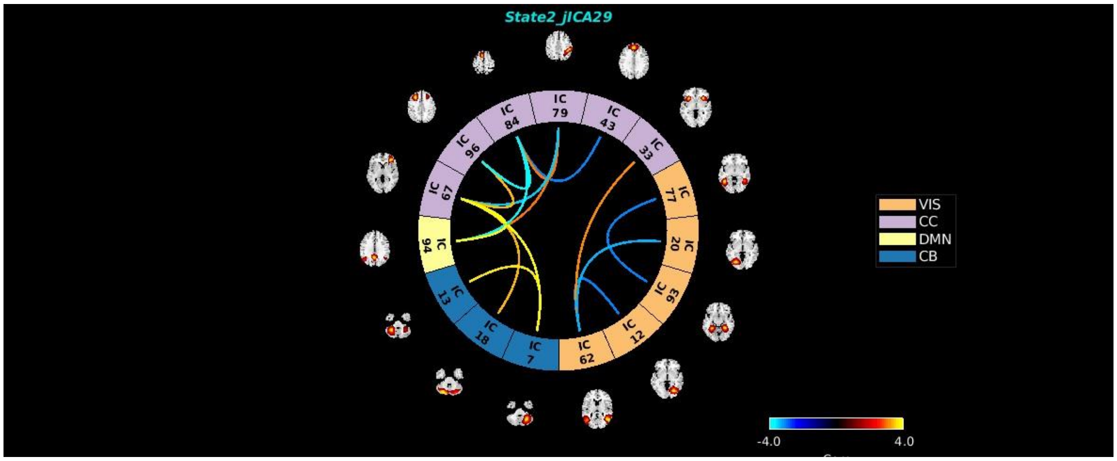
State2_jICA29
IC 79
IC 43
IC 33
IC 77
IC 20
IC 93
IC 12
IC 62
IC 7
IC 18
IC 13
IC 94
IC 67
IC 96
IC 84
VIS
CC
DMN
CB
-4.0
4.0

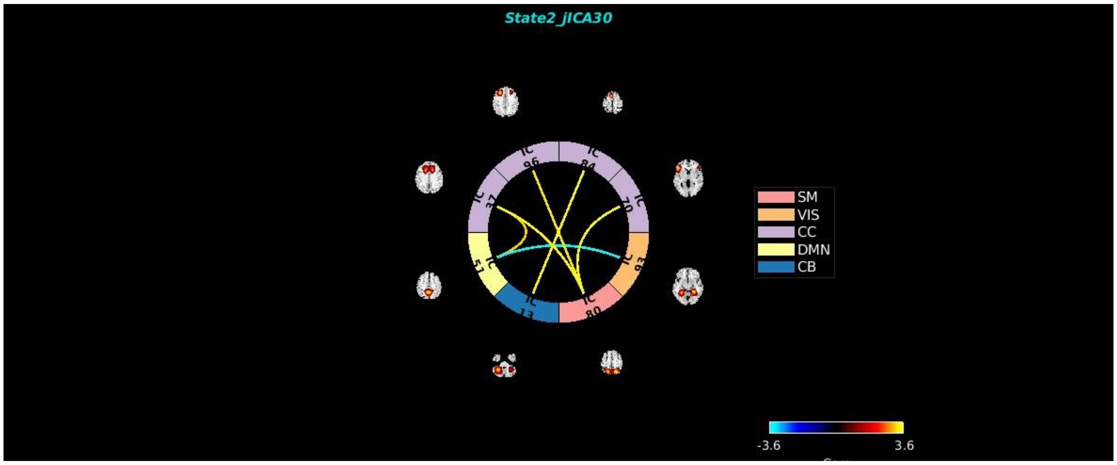
State2_jICA30
IC 96
IC 84
IC 70
IC 93
IC 80
IC 13
IC 51
IC 37
SM
VIS
CC
DMN
CB
-3.6
3.6

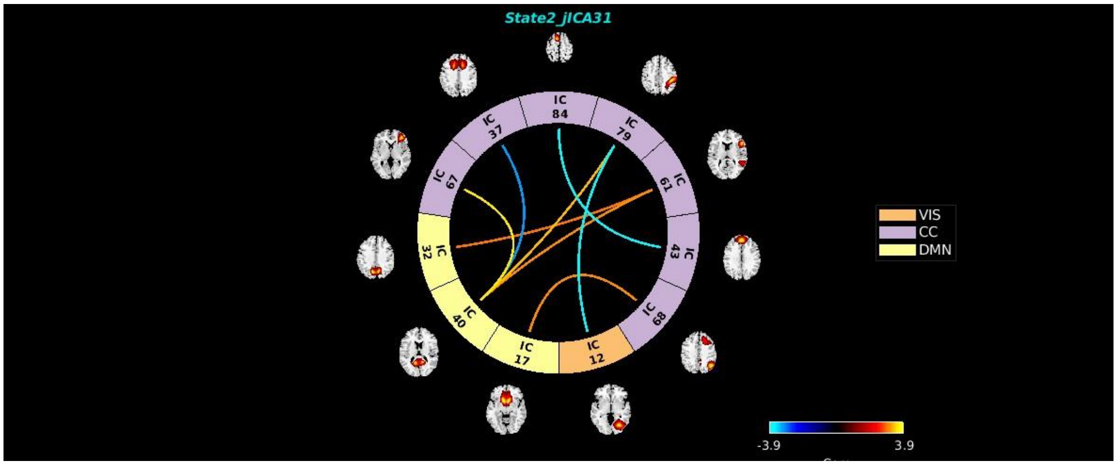
State2_jICA31
IC 84
IC 79
IC 61
IC 43
IC 68
IC 12
IC 17
IC 40
IC 32
IC 67
IC 37
VIS
CC
DMN
-3.9
3.9

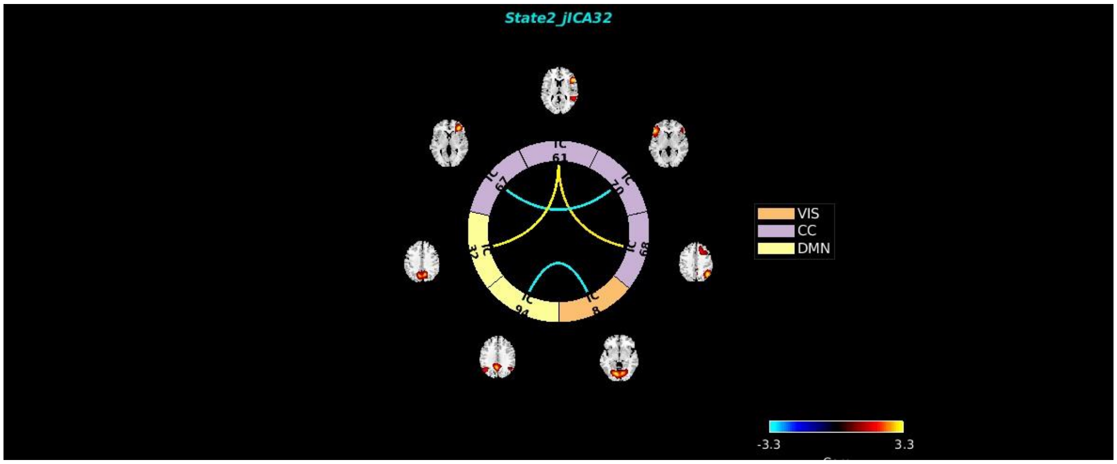
State2_jICA32
IC 61
IC 70
IC 69
IC 8
IC 94
IC 32
IC 67
VIS
CC
DMN
-3.3
3.3

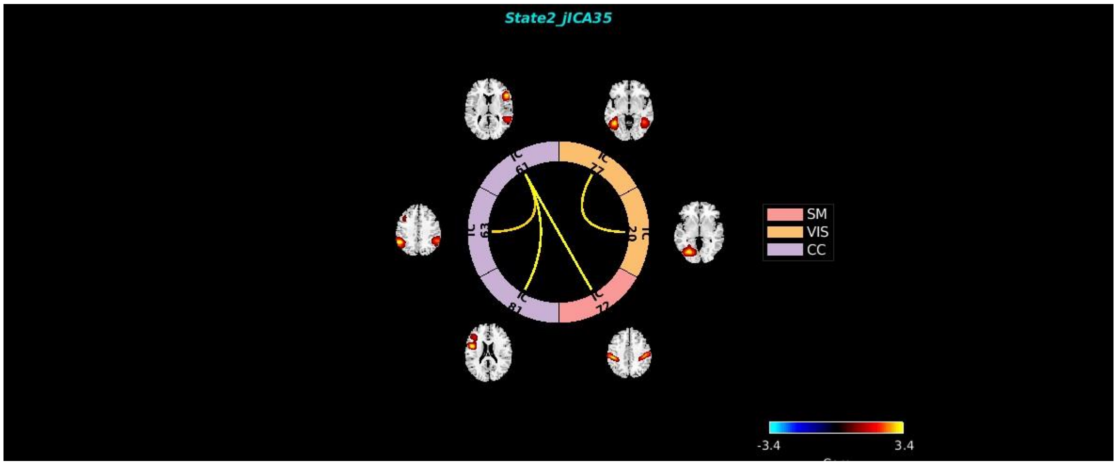
State2_jICA35
IC 61
IC 77
IC 20
IC 72
IC 81
IC 63
SM
VIS
CC
-3.4
3.4

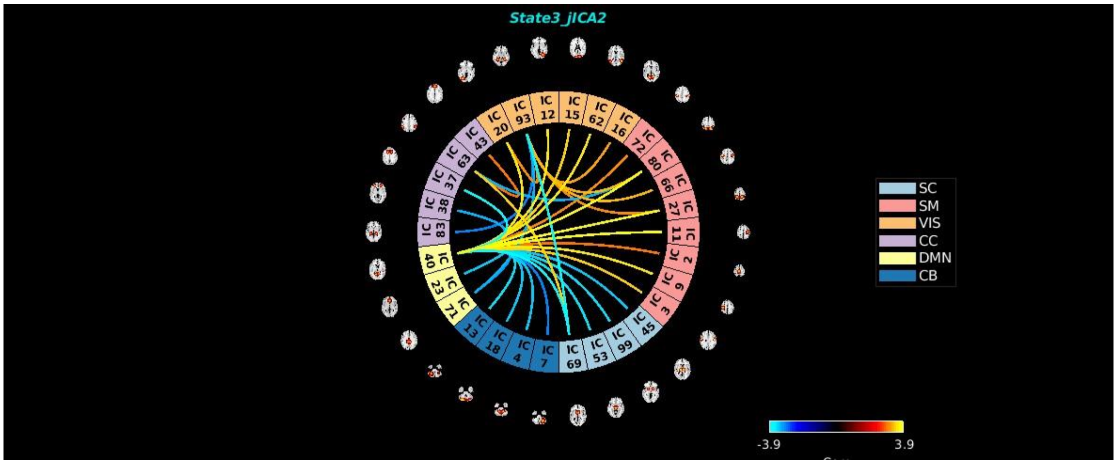
State3_jICA2
IC 20
IC 93
IC 12
IC 15
IC 62
IC 16
IC 72
IC 80
IC 66
IC 27
IC 11
IC 2
IC 9
IC 3
IC 45
IC 99
IC 53
IC 69
IC 7
IC 4
IC 18
IC 13
IC 71
IC 23
IC 40
IC 83
IC 38
IC 37
IC 63
IC 43
SC
SM
VIS
CC
DMN
CB
-3.9
3.9

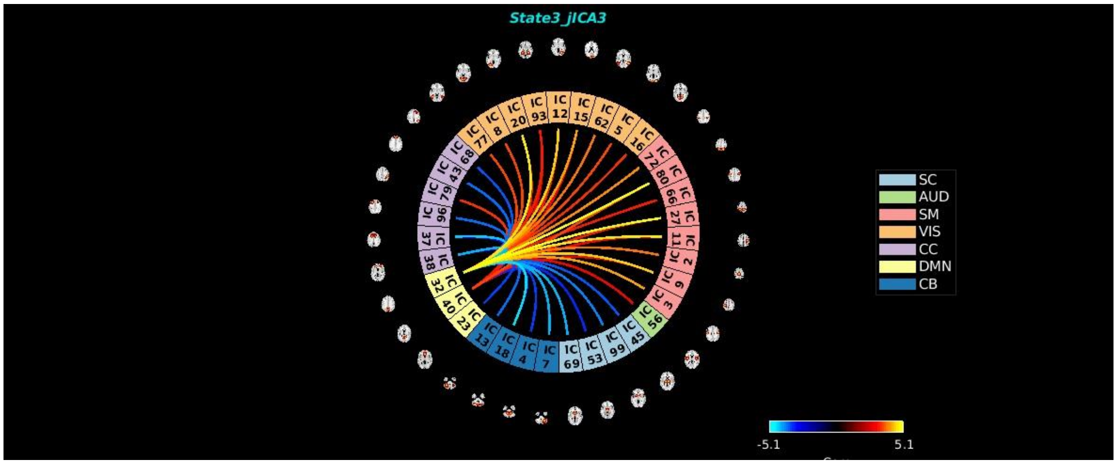
State3_jICA3
SC
AUD
SM
VIS
CC
DMN
CB
-5.1
5.1

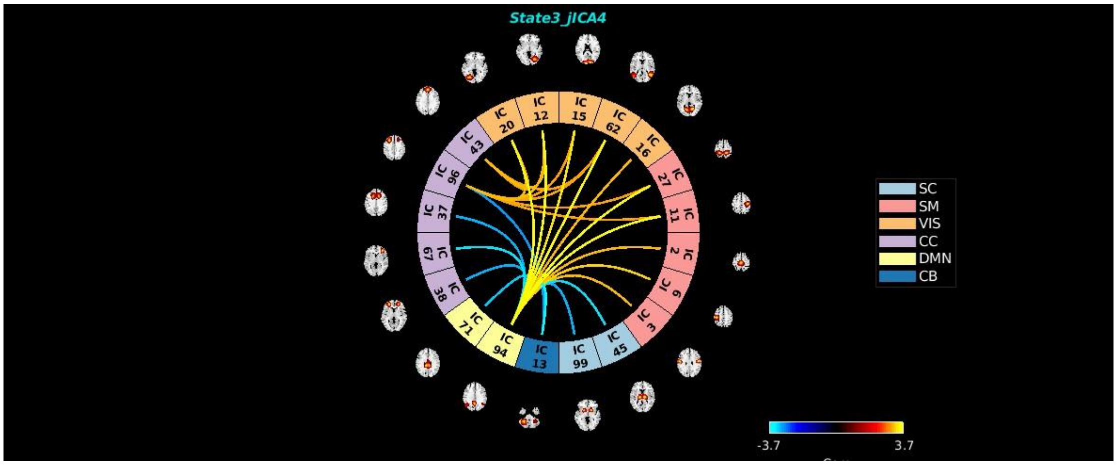
State3_jICA4
SC
SM
VIS
CC
DMN
CB
-3.7
3.7

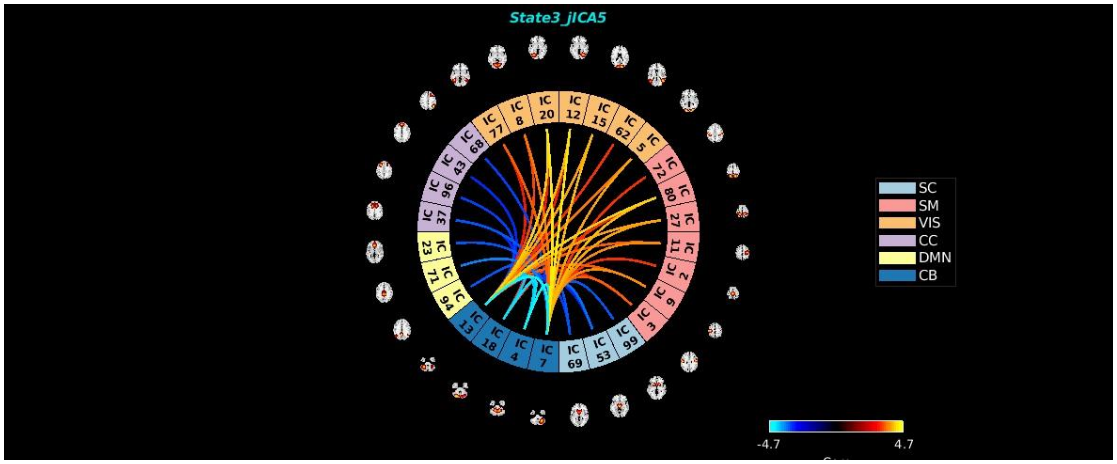
State3_jICA5
SC
SM
VIS
CC
DMN
CB
-4.7
4.7

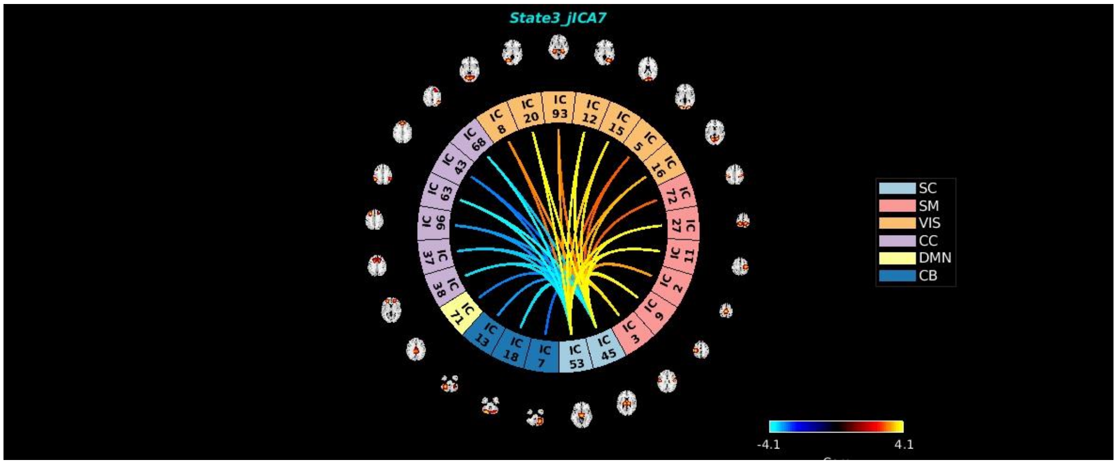
State3_jICA7
SC
SM
VIS
CC
DMN
CB
-4.1
4.1

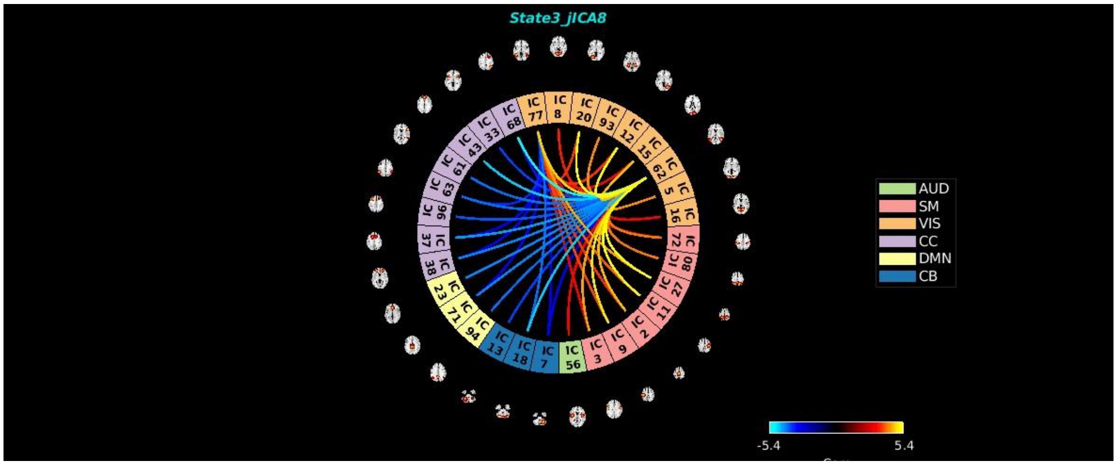
State3_jICA8
AUD
SM
VIS
CC
DMN
CB
-5.4
5.4

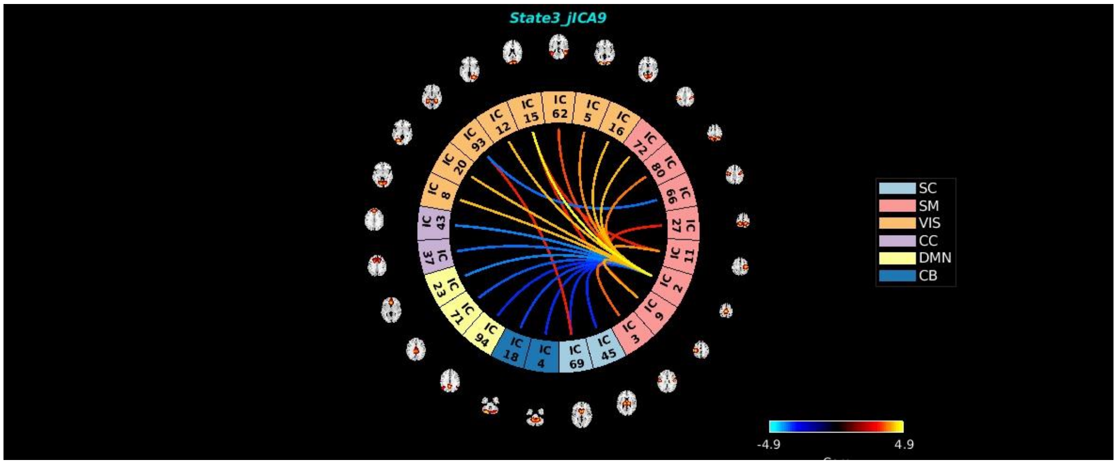
State3_jICA9
SC
SM
VIS
CC
DMN
CB
-4.9
4.9

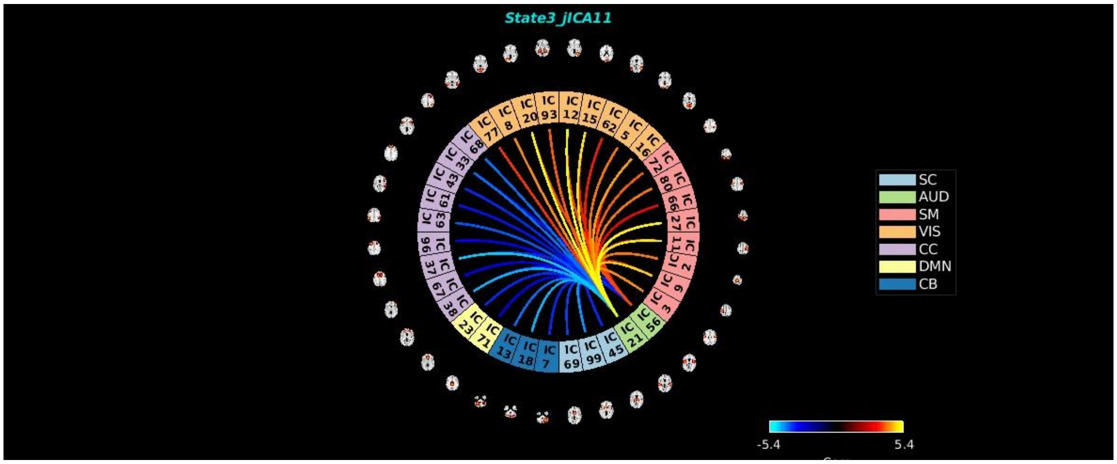
State3_jICA11
SC
AUD
SM
VIS
CC
DMN
CB
-5.4
5.4

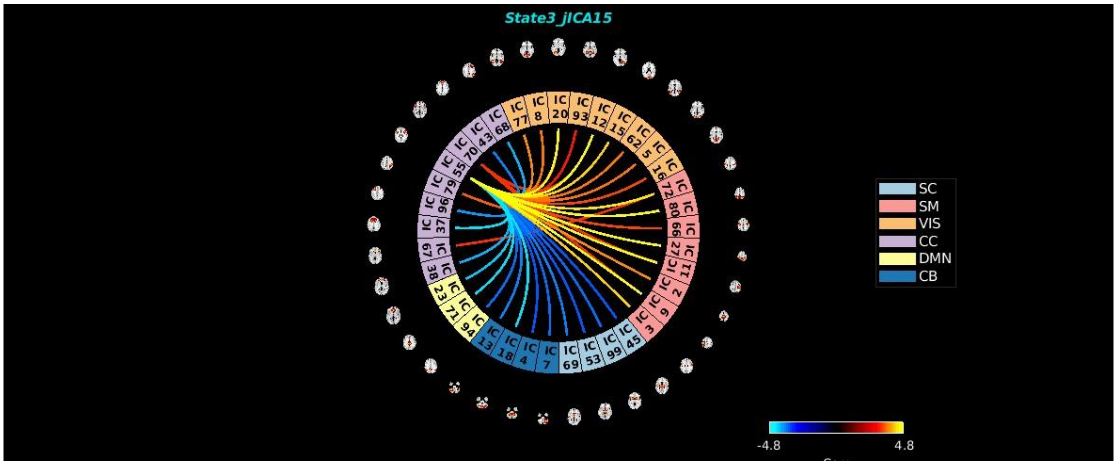
State3_jICA15
SC
SM
VIS
CC
DMN
CB
-4.8
4.8

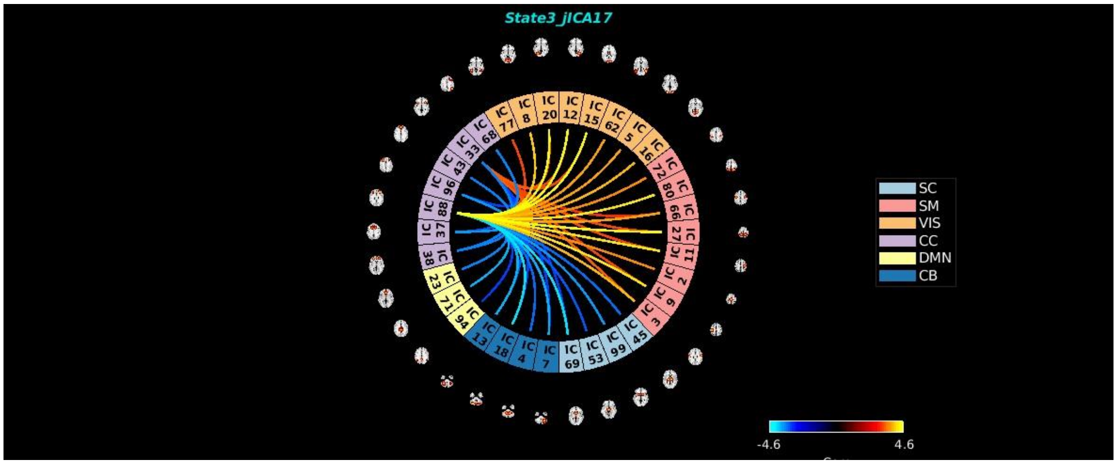
State3_jICA17
SC
SM
VIS
CC
DMN
CB
-4.6
4.6

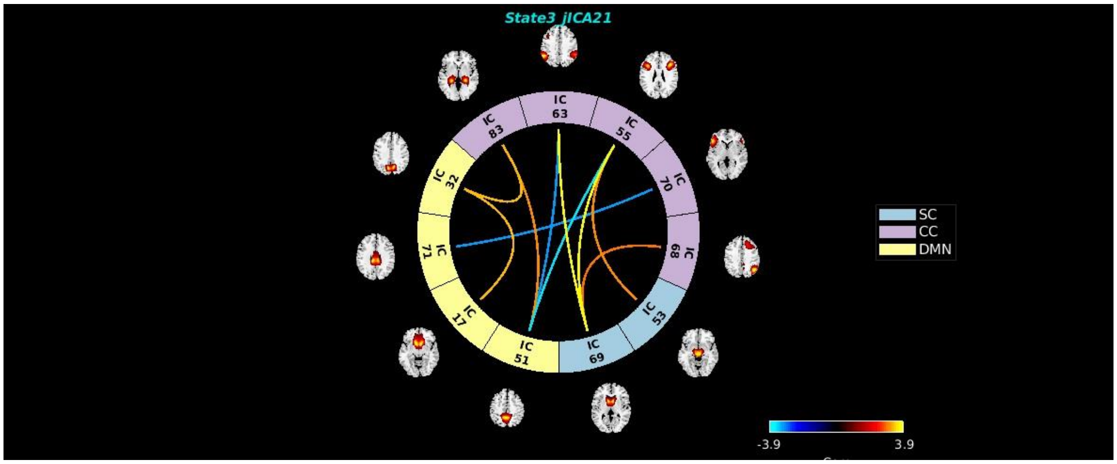
State3_jICA21
SC
CC
DMN
-3.9
3.9

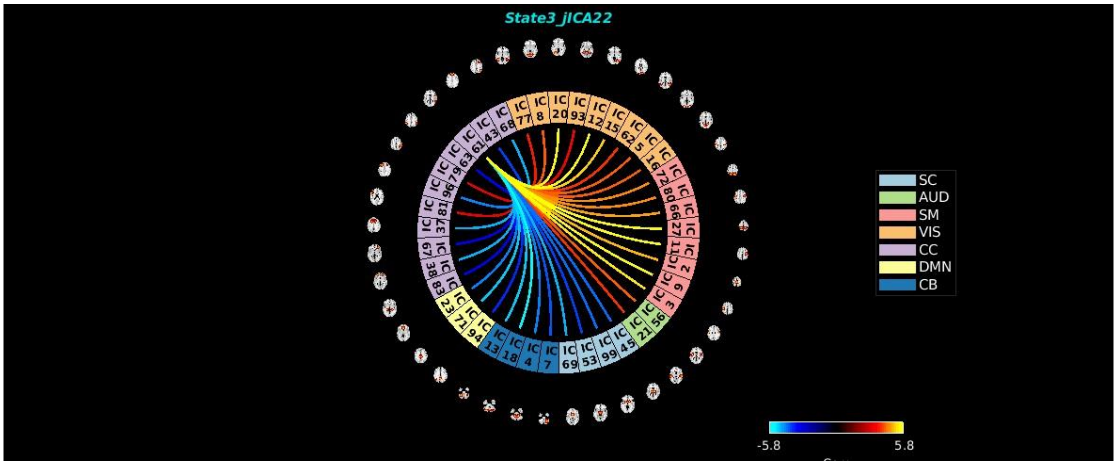
State3_jICA22
SC
AUD
SM
VIS
CC
DMN
CB
-5.8
5.8

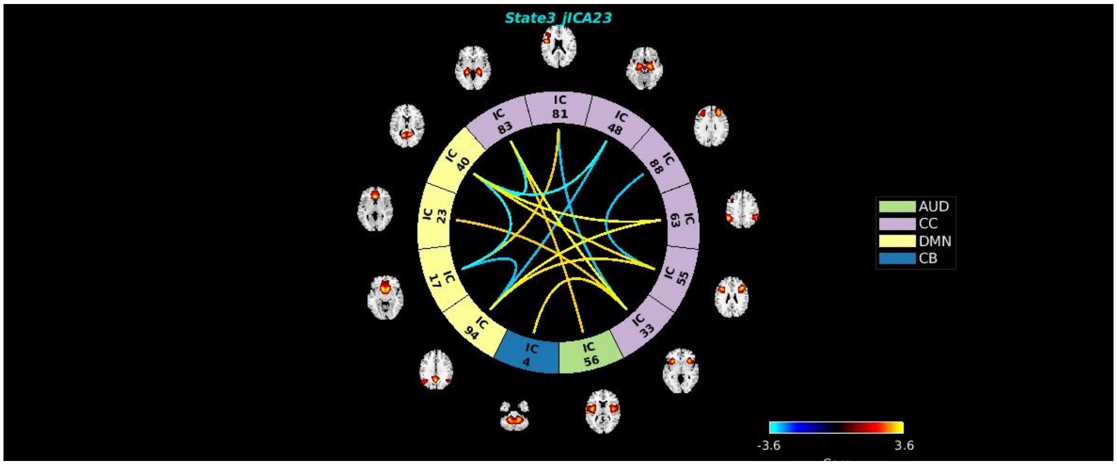
State3_jICA23
AUD
CC
DMN
CB
-3.6
3.6

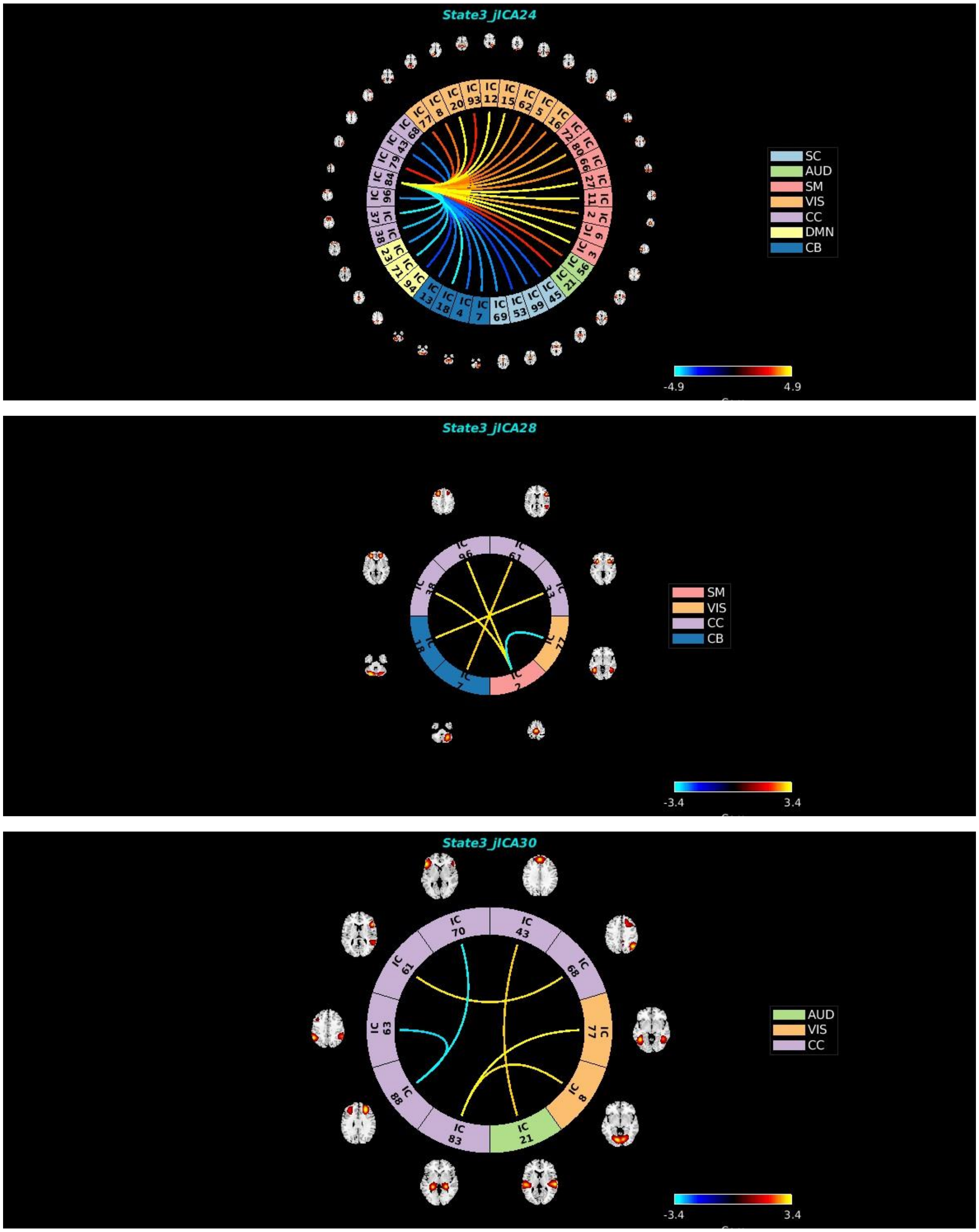
State3_jICA24
SC
AUD
SM
VIS
CC
DMN
CB
-4.9
4.9
State3_jICA28
IC 96
IC 61
IC 33
IC 77
IC 2
IC 7
IC 18
IC 38
SM
VIS
CC
CB
-3.4
3.4
State3_jICA30
IC 70
IC 43
IC 68
IC 77
IC 8
IC 21
IC 83
IC 88
IC 63
IC 61
AUD
VIS
CC
-3.4
3.4

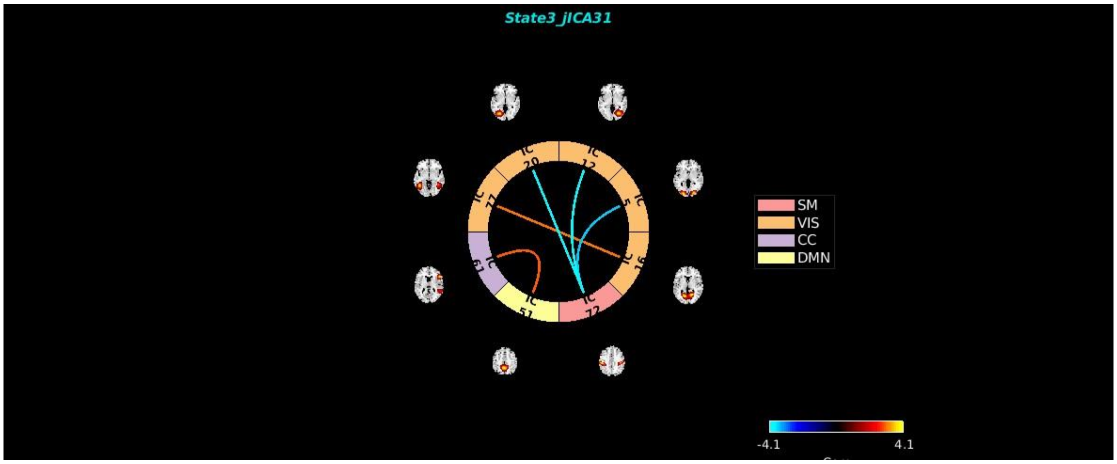
State3_jICA31
IC 20
IC 12
IC 5
IC 16
IC 72
IC 51
IC 69
IC 77
SM
VIS
CC
DMN
-4.1
4.1

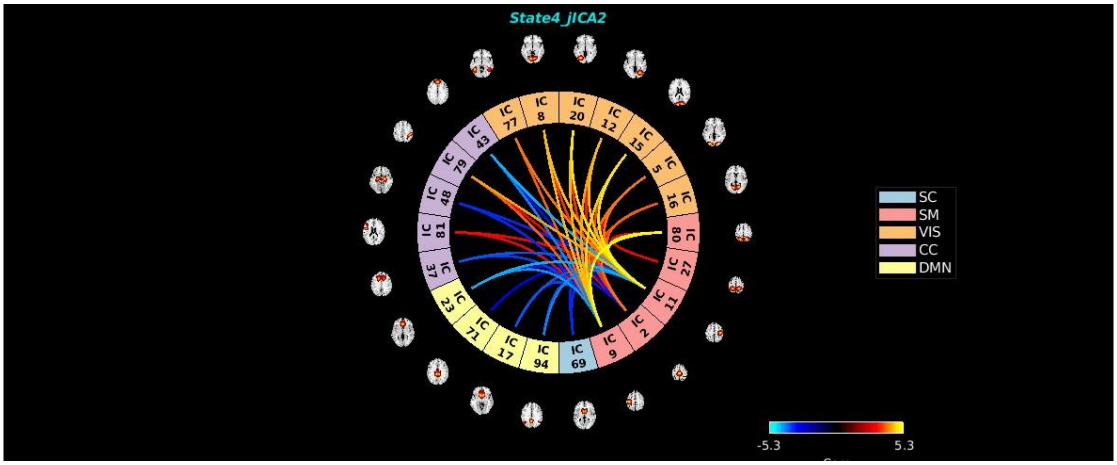
State4_jICA2
IC 77
IC 8
IC 20
IC 12
IC 15
IC 5
IC 16
IC 80
IC 27
IC 11
IC 2
IC 9
IC 69
IC 94
IC 17
IC 71
IC 23
IC 37
IC 81
IC 48
IC 79
IC 43
SC
SM
VIS
CC
DMN
-5.3
5.3

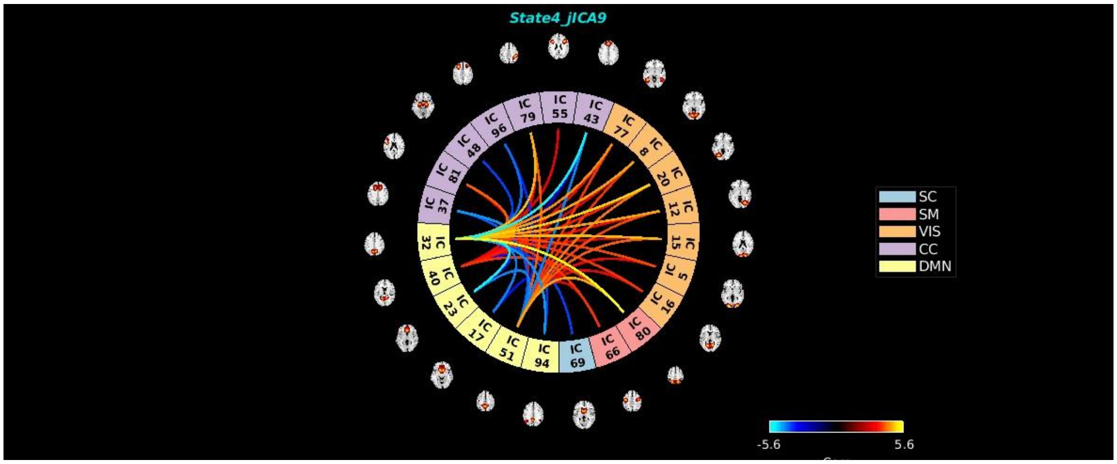
State4_jICA9
IC 79
IC 55
IC 43
IC 77
IC 8
IC 20
IC 12
IC 15
IC 5
IC 16
IC 80
IC 66
IC 69
IC 94
IC 51
IC 17
IC 23
IC 40
IC 32
IC 37
IC 81
IC 48
IC 96
SC
SM
VIS
CC
DMN
-5.6
5.6

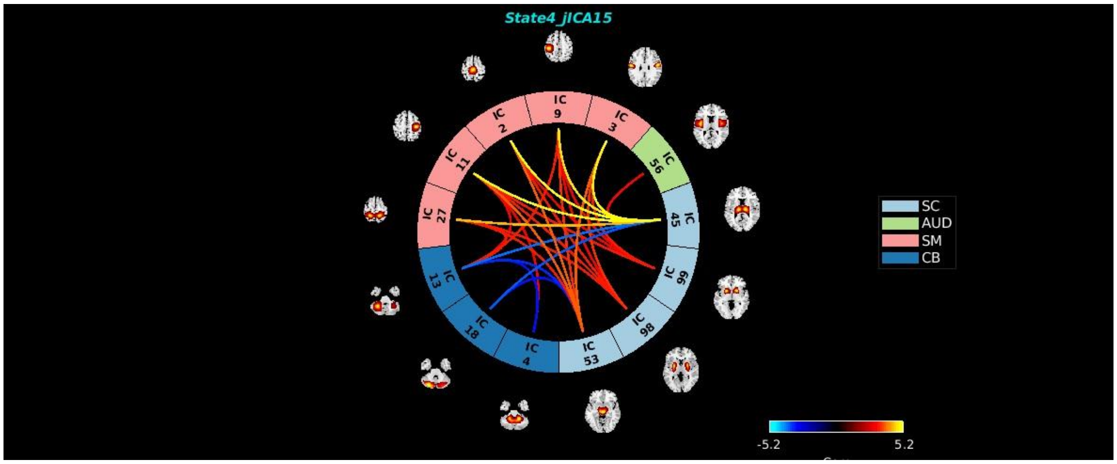
State4_jICA15
SC
AUD
SM
CB
-5.2
5.2

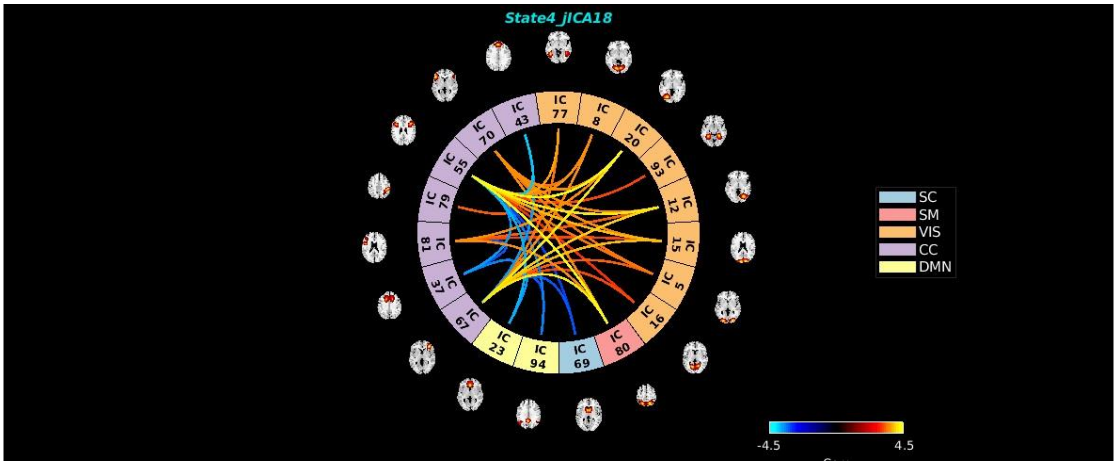
State4_jICA18
SC
SM
VIS
CC
DMN
-4.5
4.5

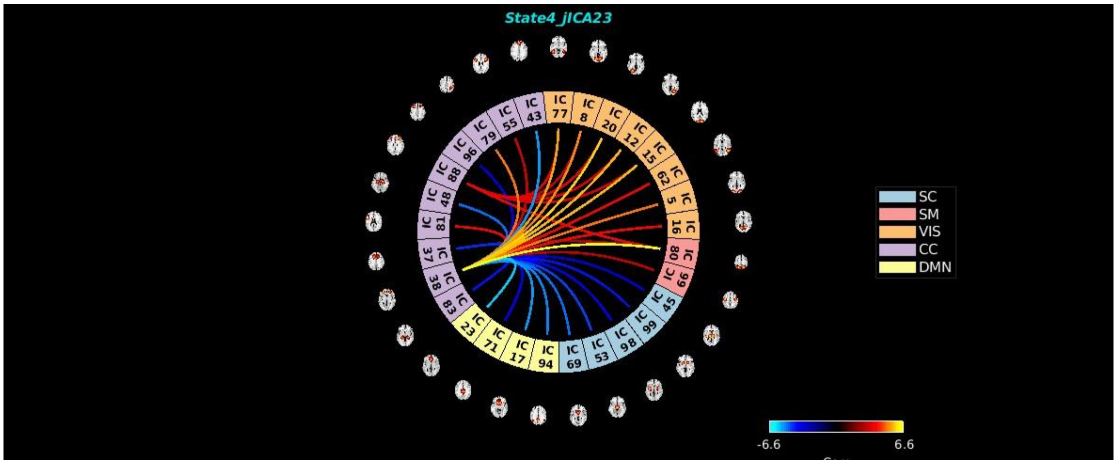
State4_jICA23
SC
SM
VIS
CC
DMN
-6.6
6.6

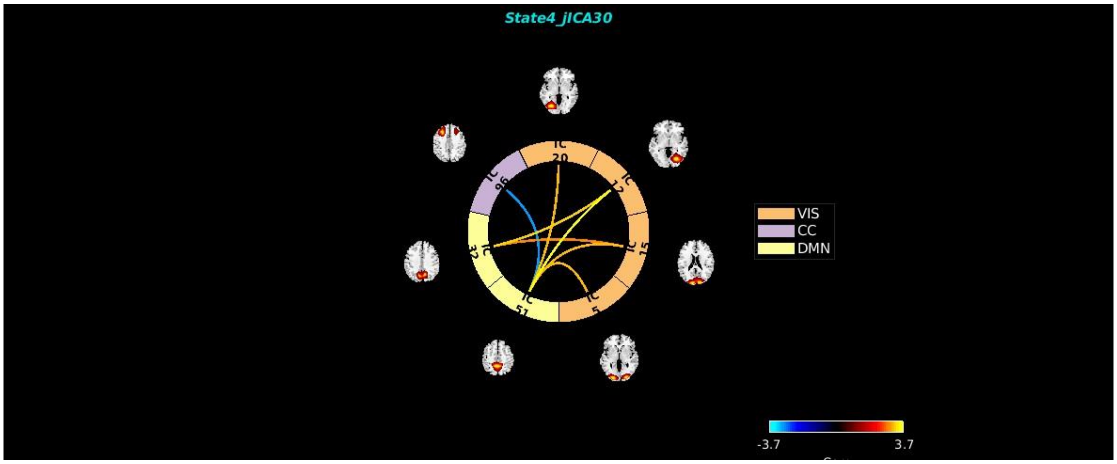


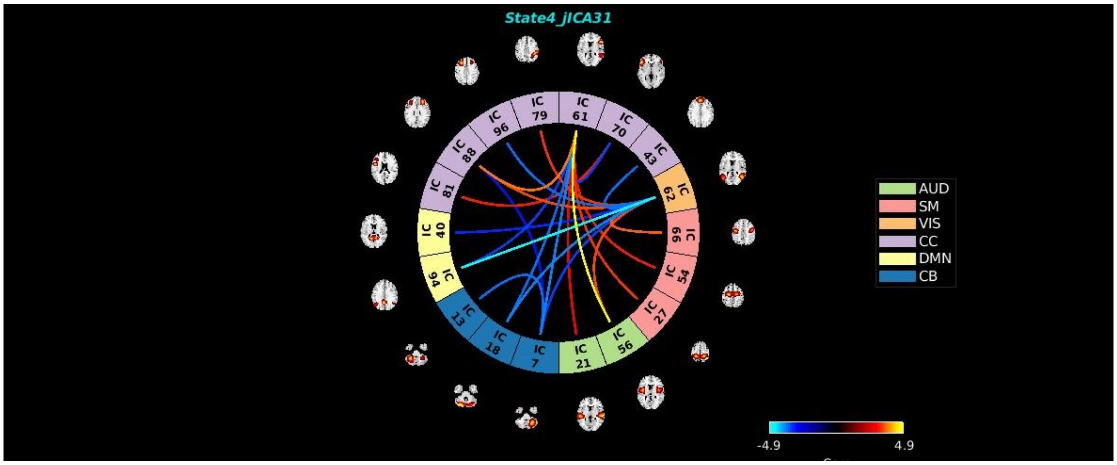


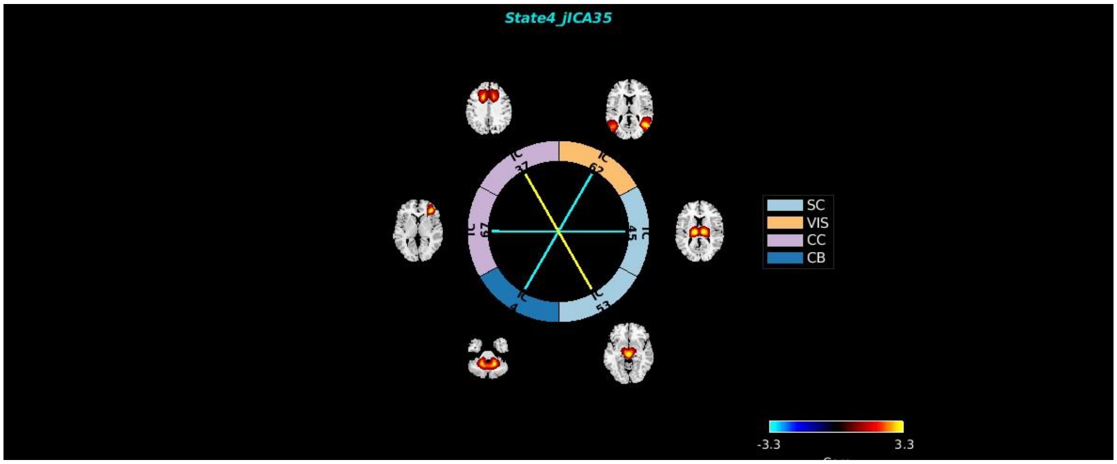


*Published with MATLAB® R2020b*
