## Supplemental File for "Dynamic fusion of genomics and functional network connectivity in UK biobank reveals schizophrenia-related SNP manifolds"

### Supplemental Text 1: resting-state fMRI data collection

COBRE study: Images were collected on a 3T Siemens Trio scanner with a 12-channel radio frequency coil. T_2_*-weighted functional images were acquired using a gradient-echo echo planar imaging sequence with TE = 29 ms, TR = 2 s, flip angle = 75°, slice thickness = 3.5 mm, slice gap = 1.05 mm, field of view 240 mm, matrix size = 64 × 64, voxel size = 3.75 mm × 3.75 mm × 4.55 mm, 150 volumes.

FBIRN study: Imaging data were collected on 3T scanners (one site used GE Discovery and the other sites used Siemens Tim Trio). Resting-state fMRI images were acquired using a standard gradient-echo echo planar imaging paradigm: FOV of 220 × 220 mm (64 × 64 matrix), TR = 2 s, TE = 30 ms, flip angle = 77°, slice thickness = 4 mm, 162 volumes.

B-SNIP study: 3T scanners with an 8-channel head coil were used for MRI acquisition at six sites. A T_2_*-weighted gradient-echo echo planar imaging sequence was used, TR = 2638 ms, TE = 30 ms, flip angle = 90°, acquisition matrix = 64 × 64; in-plane resolution = 4 mm × 4mm; 45 axial slices; slice thickness = 3 mm, 203 volumes.

MPRC study: The data collection involved three protocols. The first dataset was collected on a Siemens 3T scanner (Allegra), TR = 2000 ms, TE = 27 ms, matrix size = 64 × 64, voxel size = 3.44 mm × 3.44 mm × 4 mm, 150 volumes. The second dataset was collected on a Siemens 3T scanner (Trio), TR = 2210 ms, TE = 27 ms, matrix size = 64 × 64, voxel size = 3.44 mm × 3.44 mm × 3.99 mm, 140 volumes. The third dataset was collected on a Siemens 3T scanner (Tim Trio), TR = 2000 ms, TE = 30 ms, matrix size = 128 × 128, voxel size = 1.72 mm × 1.72 mm × 4 mm, 444 volumes.

**Supplemental Text 2: resting-state fMRI data preprocessing**

The first ten volumes were discarded to guarantee tissue reaches a steady state of radiofrequency excitation. Slice-time correction was applied using the middle slice as the reference frame. After the slicing timing, subject head motions were corrected by the realignment function in SPM12. Subjects with maximum translation of head motion greater than 3 mm or the rotation of head motion exceeding 3° in any axe were excluded. The preprocessed images were then normalized, resampled, and smoothed in the same way as for UKB.

**Supplemental Text 3: identify super-Gaussian components for further investigation**

Since Infomax ICA is expected to yield super-Gaussian components, it would be an indicator of balanced data fusion if both the SNP and dFNC part of the joint components (referred to as SNP and dFNC elements in the following text) showed super-Gaussian histograms rather than one modality dominating and the other modality under-represented in the data decomposition. Consequently, we conducted post-jICA QC to inspect the histogram and kurtosis of SNP and dFNC for each joint component. Those components with non-super-Gaussian SNP or dFNC histograms or with histogram kurtosis < 3 were excluded from the subsequent analysis
