## Supplemental Table 1 for "Dynamic fusion of genomics and functional network connectivity in UK biobank reveals schizophrenia-related SNP manifolds"

| Table S1: Annotated genes of the SZ-related components. | | |
|---|---|---|
| State | Comp | Genes |
| 1 | 2 | AFG3L1P,ARHGAP27,C1orf210,CD4,CRHR1,CTB-12O2.1,DPEP1,EFTUD1P1,FAM65B,FANCL, FTCDNL1,GRIN2A,HEATR5B,HSP90B1,IGSF9B,KANSL1,KLHL20,LINC01122,LOC100506023,LOC101243545,LOC101926964,LOC101927134,LOC101927641,LOC283177,LRRC16A,MAPT,MAPT,MAPT-AS1,MAPT-AS1,MEI1,MYBPC3,NLRP1,NRXN3,NSF,NWD2,PDCD11,PFDN1,PSMC3,RABGAP1L,ROBO1,RORB,SATB2,SLC28A1,SLC4A10,SORCS3,SPHKAP,SPIRE2,SREBF2,TMEM180,TMEM192,TMTC1,TNN,TRPV4,TTC7B,TXNRD1,WBSCR17,WNT3,WSCD2, |
| 1 | 3 | AIG1,AMZ1,ANKRD11,ARHGAP27,ARHGAP40,BANK1,BCL11B,C12orf79,C14orf23, CACNB2,CALN1,CDC25C,CDH13,CRHR1,CSMD1,CTB-12O2.1,DBNDD1,DEF8,DOCK6,DPEP1,EYS,FAM120A,FAM65B,FANCL,FOXO3,FRMD5,FYN,GALNT10,GARNL3,GATAD2A,GIGYF2,GLB1L2,GOLPH3L,GPM6A,GRM1,HCN1,HIVEP1,IGSF9B,KANSL1,KIAA1549,KLHL20,LINC00558,LINC01088,LINC01122,LOC100128239,LOC100506023,LOC101243545,LOC101927560,LOC101927587,MIR548AP,LOC101928855,RPTOR,LOC388906,LOC643733,LRP1,LRRC4B,LTBP3,MAPT,MEF2C,MEI1,MFHAS1,MIR548AJ2,MLXIP,MMP16,MSI2,MSRA,MVP,NCAM1,NPTX1,NSF,NWD2,NXPH1,PLCL1,POC1B,PRICKLE2,PRKD1,PRKD3,PRUNE,PTGIS,PUS7,RABGAP1L,RACGAP1,RELA,RGS6,RNU6-28P,TP53BP1,ROBO1,RORB,RPTOR,SF3A2,SLC39A8,SLC45A1,SLC47A2,SLC4A10,SMG1P5,SNAP91,SPATS2L,SPIRE2,SREBF2,STAG1,TDG,TNN,TRPC4,TUSC5,WNT3, |
| 1 | 4 | ANKRD44,ANKS1B,ARHGAP27,B9D1,BBX,BCL11B,C12orf43,CACNB2,CCDC63,CPEB1, CRHR1,CSMD1,CTB-12O2.1,DBNDD1,DOCK3,DPYD,EFNA5,ELFN1,EYS,FES,FUT9,GAK,GNL3,GNN,GPM6A,GPR98,GRM2,KANSL1,KCNC2,KCNJ3,KIAA1549,KLHL20,LINC00222,LINC00558,LIPC,LOC100128239,LOC100506023,LOC101927134,LOC101927273,LOC101927572,LOC101927587,LOC101929690,LRRC16A,LRRC4B,MAPT,ME1,MFHAS1,MIR548AJ2,MRPS14,NCAM1,NEURL1,NSF,NXPH1,PFKFB4,PLCL1,PSD3,PTPRD,PUS7,RABGAP1L,RASSF1,RBFOX1,RELA,RERE,RGS6,RIMS1,ROBO1,SCGN,SFXN5,SGCD,SLC28A1,SLC47A1,SMPD3,SNX8,SOX2-OT,SPHKAP,SPIRE2,SREBF2,SYNE4,TMEM180,TMEM89,TMTC1,TNN,TTC7B,VIT,WNT3,WSCD2,ZNF71, |

| | | |
|---|---|---|
| 1 | 6 | AIG1,AMZ1,ANKRD44,ANKRD44-IT1,ANKS1B,ARHGAP27,ASH2L,C12orf79,C14orf23,CA8,CACNB2,CALN1,CNTN4,CRHR1,CTB-12O2.1,CYP7B1,DCC,DOCK4,DPYD,DPYD,DPYD-AS1,EFCAB6,EFHD1,EXOC4,FANCL,FTCDNL1,FUT9,FYN,GNE,GOLPH3L,GPR98,HOMER2,IL20RB,ITPKB,KANSL1,KCNB1,KCNJ3,KLC4,KLHL20,LACE1,LINC00051,LINC00222,LINC00240,LINC01122,LOC100128239,LOC100506023,LOC100507283,LOC101927229,LOC101927587,LOC101927641,LOC101928174,LOC101928663,LRRC16A,LRRC4B,LUZP2,MAPK15,MAPT,MEF2C,MGAT3,MIR548AJ2,MRPS14,MSRA,NDRG4,NGEF,NRXN3,NSF,OPRD1,PARP8,PDCD11,PFDN1,PPP1R16B,PRUNE,RABGAP1L,RBMS3,RBMS3-AS1,RGS6,RORB,SAMD3,SATB2,SCGN,SDCCAG8,SFXN5,SGSM2,SHANK3,SLC45A1,SLC4A10,SLC9C2,SMG6,SPATS2L,SPHKAP,SULT4A1,TBC1D5,TMEM131,TMTC1,TNN,TRPC4,TSNARE1,VIT,WNT3,ZEB2, |
| 1 | 9 | ABCB4,ABHD14A,ABHD14A-ACY1,ACTC1,LOC101928174,AKAP6,AMZ1,ANKRD44,BCL11A,BTN2A1,C12orf79,C14orf23,C3orf62,CA6,CA8,CACNB2,CALN1,CAMKK2,CIR1,CNTN4,CPNE8,CRHR1,CSMD1,CTB-12O2.1,DCC,DGKI,EXOC4,EYS,FAM65B,FOXO3,GALNT10,GLTP,GNE,GPM6A,GPR98,GRAMD1B,GRIN2A,IQCF6,ITPKB,KANSL1,KCNJ3,KIAA1324L,KLF12,LGR4,LIN28B,LINC00558,LINC00624,LINC01122,LOC101926905,LOC101929690,LRRC16A,LRRC4B,MAPT,MIR548AI,MIR548AJ2,MLF2,MORN1,MSRA,MVP,NDRG4,NEBL,NLRP1,NSF,NUP210L,PARD6G,PARD6G-AS1,PCDHB16,PCGEM1,PDE4B,PFDN1,POLD1,PRICKLE2,PRKD1,PTBP2,PTGIS,PTN,RAB3C,RANGAP1,RBFOX1,RGS6,RILPL2,RIMS1,ROBO1,RPP30,SATB2,SESN2,SHANK3,SLC27A3,SLC4A10,SMPD3,SNX19,SORCS3,SOX2-OT,SP4,SPATS2L,SREBF2,TAS1R1,TMEM180,TMEM192,TMTC1,TRPC4,TRPM6,TSNARE1,TTYH3,WNT3,ZNF365,ZNF804A, |
| 1 | 10 | ADAMTSL3,AKAP6,AKT3,ALPK3,AMZ1,ARHGAP27,ASH2L,ASTN1,BAG4,BCL11A,BRE,CACNA1C,CACNB2,CALCOCO1,CALN1,CAMKK2,CCDC134,CHRNA2,CPEB1,CPNE8,CRHR1,CSMD1,CTB-12O2.1,DEF8,DPP4,FAM65B,FOXO3,GAK,GBF1,GLB1L2,GNN,GPATCH11,GPM6A,GPR98,HEATR5B,HIVEP1,HOMER2,HSP90B1,ITPKB,KANSL1,KCNJ3,KLF12,KLHL20,LACE1,LHFPL3,LOC100506023,LOC100506472,TAB1,LOC100507283,LOC101927134,LOC101927641,LOC101929567,LOC388906,LOC646522,LRRC16A,LRRC4B,LSM1,MAD1L1,MAPT,MEF2BNB-MEF2B,MEF2C-AS1,MFHAS1,MIR548AI,MOB3A,MYBPC2,MYO1E,NALCN,NEBL,NRXN3,NWD2,NXPH1,PAK6,PARD6G-AS1,PDE4B,PLCH2,PLCL1,PRKD1,PRR13,PSD3,PSMC3,PTK2B,RAB3C,RANGAP1,RASSF1,RER1,RNF111,RPARP-AS1,RPP30,SATB2,SFXN5,SGCD,SLC28A1,SLC4A10,SNX8,SPATS2L,SYNE4,TACC2,TBC1D2,TDG,TMPRSS12,TNKS,UBE2Q2P1,WNT3, |

| | | |
|---|---|---|
| 1 | 12 | ADAM22,AKAP6,AKT3,SDCCAG8,ANKRD11,ANKS1B,BCL11B,C1orf106,CA6,CA8,CACNA1S,CACNB2,CCDC39,CD4,CHKB-AS1,CIR1,CRHR1,CRK,CSMD1,CTB-12O2.1,DCC,DOCK6,EFNA5,EXOC4,EYS,FAM65B,FANCL,FYN,GRM2,KANSL1,KIAA1549,KLF12,KLRG2,LIN28B,LINC00558,LINC00624,LOC100506472,TAB1,LOC101927641,LOC101928174,LRRC16A,LTBP3,LUZP2,MAPT,MED27,MEI1,MIR548AJ2,MKL1,MSRA,MYO1E,NDUFAF7,NEGR1,NLRP1,NSF,OLA1,P2RX7,PARD6G-AS1,PDE4B,PRICKLE2,PSD3,PTGIS,PTN,PTPRD,RBFOX1,RILPL2,RNF111,ROBO1,RPP30,RPTOR,SATB2,SGCD,SLC28A1,SLC38A7,SLC45A1,SLC4A10,SMPD3,SORCS3,SPPL3,TACC2,TMTC1,TRPM6,TXNRD1,UBE2Q2P1,VIT,WBSCR17,WNT3, |
| 1 | 13 | AKAP6,ALG14,ANGPTL2,RALGPS1,ANKK1,ANKS1B,ARHGAP27,BBX,BCL11B,BTN2A1,C12orf65,C1orf106,CACNA1D,CACNA1S,CACNB2,CALN1,CAMKK2,CD4,CDIP1,CKAP5,CRHR1,CSMD1,CTB-12O2.1,DBNDD1,DOCK4,DPEP1,DPYD,DPYD,DPYD-AS1,EFHD1,EFNA5,EXOC4,EYS,FES,GABBR2,GAK,GNE,GPM6A,GRM1,GRM8,HIVEP1,HSP90B1,IMMP2L,IQCE,KANSL1,KCNC2,KIAA1324L,KIF21B,KLHL20,LINC00320,LINC01122,LOC100128239,LOC100506023,LOC101927134,LOC101927641,LOC283177,MAPT,MAPT,MAPT-AS1,MAPT-AS1,MAPT-AS1,SPPL2C,MEI1,MIR548AI,MIR548AJ2,MORN1,MSRA,MYBPC3,NGEF,NLRP1,NRXN3,NSF,NWD2,NXPH1,OPRD1,PCDH17,PCGEM1,PDE4B,POLD1,PPP1R16B,PRKD1,PSMC3,RAB3C,RABGAP1L,RBFOX1,RER1,RGS6,RPP30,SAMD3,SESN2,SGK223,SHANK3,SLC4A10,SMG6,SMPD3,SOX2-OT,SP4,SPHKAP,SPIRE2,TBC1D5,TIE1,TMEM180,TMEM192,TNN,WNT3,ZDHHC12, |
| 1 | 16 | ABHD14A,ABHD14A-ACY1,AIG1,AKAP6,ALPK3,ANKK1,ANKRD11,ANKRD44,ANKS1B,BBX,C12orf65,C1orf106,CALN1,CAMKK2,CDH13,CDK10,CHCHD3,CNTN4,CPEB1,CPNE7,CRHR1,CSMD1,CTB-12O2.1,DEF8,DPP4,DPYD,EFCAB6,EFTUD1,ELFN1,FANCL,FES,GAS8,GNN,GPR98,GRAMD1B,HOMER2,HSP90B1,ITPKB,KIF21B,KLF12,KLHL20,LHFPL3,LHFPL3,LHFPL3-AS2,LINC00051,LINC00558,LINC00635,LINC01122,LOC100506023,LOC101926905,LOC101927572,LOC101927641,LOC101928174,LOC646522,LRRC16A,MAD1L1,MAPT,MAPT,MAPT-AS1,MIR548AI,MIR548AJ2,MLXIP,MORN1,MSRA,NALCN,NSF,OASL,OLA1,OPRD1,P2RX4,P2RX7,PAK6,PLCH2,POC1B,PRICKLE2,PTGIS,RABGAP1L,RBFOX1,RER1,RGS6,RILPL2,RIMS1,RORB,SAMD3,SLC28A1,SLC4A10,SNAP91,SORCS3,SPATS2L,SPIRE2,TACC2,TBC1D5,TBL1XR1,TDG,TMEM192,TRPC4,TSPAN16,VPS72,WNT3,WSCD2,ZNF365, |

| | | |
|---|---|---|
| 1 | 19 | AKAP6,AMZ1,ANKS1B,ANXA9,ARFGAP2,BBX,BCL11A,BCL11B,C14orf23,CACNA1C,CACNA1D,CACNB2,CALN1,CAMKK2,CCDC63,CD4,CIR1,CKAP5,CLIP1,CNTN4,CPEB1,CSMD1,CTB-12O2.1,CTDP1,DNAH11,DOCK3,DOCK6,DPEP1,DPYD,EIF2AK2,ELFN1,EPPK1,FAM65B,FES,FOXO3,FTCDNL1,GAK,GAPT,GNN,GPR98,GRM2,HEATR5B,HIVEP1,KCNB1,KLF12,KLF6,LACE1,LHFPL3,LHFPL3,LHFPL3-AS2,LINC00624,LINC00936,LINC01122,LOC100505474,LOC101927134,LOC101927273,LOC101927587,LOC101927641,LOC101928782,LRP1,LRRC16A,LRRC4B,LTBP3,MAD1L1,MED27,MEF2B,MEF2BNB-MEF2B,MIR548AJ2,MIR548AP,TTLL7,MSRA,MYBPC3,NALCN,NDST3,NDUFAF7,NGEF,PCGEM1,PDE4B,PFKFB4,PLCL1,PPP1R16B,RABGAP1L,RASSF1,RBFOX1,RBMS3,RELA,RGS6,RIMS1,ROBO1,RRAS,SATB2,SESN2,SFXN5,SGCD,SGK223,SGSM2,SLC28A1,SLC4A10,SLC9C2,SNX8,SOX2-OT,SPATS2L,SPHKAP,SPIRE2,STAG1,SYNE4,TDG,TMEM131,TMEM180,TMTC1,TSPAN16,TTC7B,WSCD2,ZEB2,ZNF804A, |
| 1 | 29 | CRHR1,CTB-12O2.1,HCN1,LOC101928663,LRRC16A,PARP8,SCGN,SLC4A10,WNT3, |
| 1 | 30 | MFHAS1,MSRA,SGK223, |
| 1 | 33 | CTB-12O2.1,KLHL20,LOC100506023,MMP16,RABGAP1L, |
| 2 | 2 | ACD,ACTC1,LOC101928174,ADAMTSL3,AKAP6,ANKRD11,ANXA9,ARHGAP27,ARHGAP40,B3GAT1,BBX,C12orf79,C1orf106,CA8,CACNA1C,CACNB2,CALN1,CCDC39,CD4,CRHR1,CSMD1,CTB-12O2.1,CTDP1,DGKI,DPYD,EIF2AK2,FAM65B,FANCL,FRMD5,FTCDNL1,FYN,GAPT,GARNL3,GLB1L2,GLI1,GNAI1,GNL3,GOLPH3L,GPM6A,GPR98,GRM1,KANSL1,KCNJ3,KLHL2,LINC00635,LINC01122,LOC100506023,LOC101928882,LOC101929690,LOC283177,LRRC16A,LRRC4B,LSM1,LTBP3,MAPT,MAPT-AS1,MEF2C,MEF2C,MEF2C-AS1,MFAP1,MIR548AJ2,MSRA,NALCN,NDRG4,NDST3, NDUFAF7,NSF,ORC5,OSBPL3,PAK6,PARD6G-AS1,PDE4B,PFDN1,PLCL2,PRICKLE2,PTK7,QPCT,RAB3C,RABGAP1L,RALGPS1,RBMS3,RELA,RELN,RIMS1,RNU6-28P,TP53BP1,ROBO1,RORB,RPTOR,SGCD,SH3RF1,SLC28A1,SLC39A8,SLC9C2,SNX19,SNX8,SORCS3,SULT4A1,TACC2,TMEM192,TMTC1,TRPC4,TRPM6,TTC7B,TUSC5,VIT,ZAP70,ZEB2, |
| 2 | 4 | ABCB1,AC093375.1,AKAP6,ALG14,ALPK3,AMZ1,ARHGAP40,BBX,C16orf96,CACNA1S,CACNB2,CAMKK2,CD4,CDC23,CDC25C,CDIP1,CNTN4,CPNE7,CPNE8,CTB-12O2.1,DLX2,DOT1L,DPP4,FUT9,GABBR2,GMIP,GPM6A,GRAMD1B,HEATR5B,IQCE,KCNB1,KCNJ3, KIAA1549,KLF12,KLHL20,KLRG2,LACE1,LINC00051,LINC00320,LOC101926964,LOC101927134,LOC101927572,LOC101927641,LOC101929690,LRRC16A,LTBP3,LUZP2,MAPT,MED27,MEF2BNB,MEF2BNB-MEF2B,MEF2BNB-MEF2B,MEF2C,MEF2C-AS1,MIR548AJ2,MORN1,MSANTD2,MSRA,MYO1E,NALCN,NSUN6,NXPH1,P2RX7,PAK6,PFDN1,PLCL1,PLCL2,PPP1R16B,PTPRD,RAB3C,RABGAP1L,RBFOX1,RGS6,RNF111,RORB,RPTOR,SDCCAG8,SGK223,SLC39A8,SLC4A10,SMPD3,SNAP91,SPATS2L,SPHKAP,SPRYD3,SYNE4,TCF4,TMEM192,TMEM200A,TMEM56,TMTC1,TNN,TRAF3IP2,TRAF3IP2-AS1,TSNARE1,TTC7B,WNT3, |

| | | |
|---|---|---|
| 2 | 5 | ABCB1,ABCB4,ABCD2,ACD,ACTC1,LOC101928174,ADAMTSL3,AIG1,AKAP6, ALPK3,BANK1,BBX,C1orf95,CA8,CACNA1D,CACNB2,CD4,CNTN4,CPNE8,CRHR1,CSMD1,CTB-12O2.1,DNAH11,DPYD,DPYD-AS1,EFNA5,EPPK1,EYS,FAM65B,FTCDNL1,FUT9,FYN,GLB1L2,GLTP,GOLPH3L,GRAMD1B,GRIN2A,GRM3,GTDC1,HAT1,HOMER2,HSP90B1,ITPKB,KCNH6,KCNJ3,KIAA1549,LAG3, LIPC,LOC101927587,LOC101928174,LOC101928569,LOC101928663,LOC101928882, LOC101929690,LRRC16A,LTBP3,MAD1L1,MAN2A1,MANBA,MAPT,MSI2,MVP,MYO1E,NDRG4,NFKB1,NSF,NUP210L,NXPH1,OSBPL3,PAK6,PARP8,PCDHB16,PRDM7,PRICKLE2,PTBP2,RAB3C,RGS6,SCGN,SHANK3,SLC28A1,SLC38A7,SLC39A8,SLC4A10,SMG1P5,SMPD3,SNX8,SPATS2L,SPHKAP,STAG1,TDG,TMEM192,TTC7B,TTYH3,UBD,UBE2Q2P1,WNT3,WSCD2, |
| 2 | 8 | ACD,ADAMTSL3,ALPK3,AMZ1,ANKK1,ANKRD11,ANKRD44,C1orf106,C1orf210, CA8,CACNA1D,CACNB2,CALN1,CAMTA1,CD4,CDC23,CDIP1,CDK10,CIR1,CORO7,CORO7-PAM16,CPNE7,CPNE8,CRHR1,CTB-12O2.1,CTCF,CTDP1,CYP7B1,DBNDD1,DEF8,DFNA5,DOCK3,DPEP1,DPYD,DPYD-AS1,EIF2AK2,ENOX1,FAM65B,GLTP,GMIP,GNN,GPM6A,GRIN2A,HEATR5B,HIP1R,IQCE,IQCF6,KCNJ3,KLHL20,KLRG2,LINC01122,LOC101927134,LOC101927587,LOC101928569,LOC101928663,LRRC16A,LRRC36,MED27,MEF2BNB-MEF2B,MEF2C,MFHAS1,MIR548AI,MIR548AJ2,NALCN,NGEF,OPCML,PARD6G-AS1,PLCL2,POLD1,PRDM7,PRICKLE2,PSD3,RABGAP1L,RASSF1,RBFOX1,RBMS3,RILPL2, ROBO1,SCRIB,SHANK3,SLC28A1,SLC4A10,SLC7A6OS,SMPD3,SORCS3,SOX2-OT,SPIRE2,STAG1,TBC1D5,TCF4,TDG,TMTC1,TTC7B,TUSC5,UBE2Q2P1,WBSCR17,WNT3,WSCD2,YWHAE, |
| 2 | 9 | ALPK3,ANXA9,BCL11B,C8orf86,CACNA1D,CACNA1S,CACNB2,CALN1,CAMTA1, CDC25C,CKAP5,CNTN4,CPNE8,CSMD1,CTB-12O2.1,DCC,DPP4,DPYD,DPYD-AS1,EFNA5,FAM65B,FAM83G,SLC5A10,FUT9,GNAI1,GPM6A,IGSF9B,ITPKB,JAM3,LAG3,LINC00051,LOC100128239,LOC101926905,LOC101927134,LOC101928663,LOC283335,LRRC16A,LRRC4B,LTBP3,LUZP2,LYSMD1,TNFAIP8L2-SCNM1,MAD1L1,MEI1,MFHAS1,MIR548AI,MIR548AJ2,MLF2,NEBL,NGEF,NUP210L,NXPH1,ORC5,OSBPL3,PARD6G-AS1,PDIA3,PFDN1,PHGR1,PLCL1,PSD3,PTGIS,RASSF1,RBFOX1,RBMS3,RELA,RERE,RILPL1,ROBO1,RPRD2,RPTOR,SLC39A8,SLC4A10,SLC5A10,SMIM17,SNX19,SPATS2L,SPRYD3,ST3GAL3,TACC2,TCF4,TMEM131,TMEM192,TRIM31,TRPC4,TRPV4,TUSC5,UBD,WNT3, ZNF365, |
| 2 | 12 | ACR,ACTC1,LOC101928174,ALG14,BANK1,BBX,BLM,BRE,BTN2A1,C1orf210, C4orf27,CACNB2,CAMTA1,CKAP5,CLCN3,CNTN4,CSMD1,DLX2,DPYD,ELFN1,FAM120A, FAM124B,FAM65B,FOXO3,FTCDNL1,GLTP,GNL3,GNN,GPM6A,GPR98,HAT1,KLHL2,KLHL20,LIN28B,LOC101928174,LOC101929690,LOC283335,LRRC16A,MAD1L1,MAN2A1, MAPT,ME1,MED27,MEF2C,MEF2C-AS1,MIR548AJ2,MYO1E,NGEF,NPTX1,OPCML,OSBPL3,PCDH17,PLCL1,PLCL2,PTBP2,QPCT,RAB3C,RABGAP1L,RELA,RILPL2,ROBO1,RORB,RPRD2,RPTOR,SAMD3,SCGN,SDCCAG8,SFXN2,SLC25A12,SLC28A1,SLC45A1,SORCS3,TBC1D5,TBL1XR1,TDG,TMEM56,TMTC1,TRPC4,TRPV4,WBSCR17,WNT3,YWHAE,ZNF71, |

| | | |
|---|---|---|
| 2 | 22 | ADAM22,ADAMTSL3,ARHGAP27,C1orf106,CACNA1S,CACNB2,CAMKK2,CLIP1,CNTN4,CPEB1,CPNE8,CRHR1,CSMD1,CTB-12O2.1,CTDP1,DBNDD1,DEF8,DFNA5,DNAH1,DOCK6,DOT1L,DPYD,DPYD,DPYD-AS1,EXOC4,EYS,FES,FOXO3,GABBR2,GNL3,GPM6A,GPR98, HSP90B1,IGSF9B,INPP4B,IQCE,ITPKB,KANSL1,KLF12,LACE1,LGR4,LHFPL3,LHFPL3-AS2,LINC01122,LIPC,LOC101928174,LOC101928782,LOC646522,LRRC16A,LRRC4B,LRRC8A,LUZP2,MAPT,MFHAS1,MIR548AI,MSRA,MYBPC3,NRXN3,NSF,NSUN6,NXPH1,P2RX4,PARP8,PDCD11,PFDN1,PLCL2,PRDM7,PRICKLE2,PRKD1,PRUNE,PSMC3,PTK2B,QPCT,RANGAP1,ROBO1,RPTOR,SFXN5,SLC28A1,SLC39A8,SLC4A10,SMPD3,SORCS3,SOX2-OT,SPATS2L,SPHKAP,SPIRE2,ST3GAL3,TACC2,TCF4,TDG,TMEM180,TMTC1,TRPC4,TXNRD1,VIT,WBSCR17,WNT3,ZDHHC12,ZNF184,ZNF804A, |
| 2 | 25 | ALG14,ANKK1,ANKS1B,ARHGAP27,BBX,C3orf62,CA6,CACNA1C,CACNB2, CALN1,CDIP1,CHKB-AS1,CNTN4,CPNE8,CRHR1,CSMD1,CTB-12O2.1,CTC-436P18.1,CTDP1,DCC,DOCK3,DPEP1,DPP4,DPYD,EFCAB6,EIF4B,FAM120A,FYN,GLB1L2,GPR98,GRAMD1B,GRM3,IGSF9B,KCNC2,KCNJ3,KCTD18,KLF12,LIPC,LOC101929690,LOC283177,LOC388906,LRRC16A,LRRC4B,MAD1L1,MAPT,MLXIP,MSRA,NDRG4,NDST3,NEBL,NPTX1,NRXN3,NSF,NWD2,NXPH1,OSBPL3,P2RX7,PAK6,PARD6G,PCDHB16,PDE4B,PLCL2,PTK2B,RAB3C,RANGAP1,RBFADN,RBFOX1,RBMS3,RBMS3-AS1,RNU6-28P,TP53BP1,RORB,RPTOR,SCGN,SH3RF1,SLC39A8,SLC4A10,SLC5A10,SMPD3,SORCS3,SPHKAP,SPIRE2,SPRYD3,ST3GAL3,TMEM56,TRPC4,TUSC5,WBSCR17,WNT3,YWHAE, |
| 2 | 27 | ABCB1,AKT3,AKT3,SDCCAG8,ALG14,ATP2A2,BBX,BCL11A,BLM,BRE,C3orf62, CACNA1C,CACNB2,CAMKK2,CCDC134,CDIP1,CRHR1,CSMD1,CTB-12O2.1,DFNA5,DPH1,DPYD,EFNA5,EXOC4,EYS,FAM65B,GABBR2,GLTP,GNN,GPM6A,GPR98,GPX6,INPP4B,KCNJ3,KIAA1549,KLF6,LGR4,LINC00635,LINC01122,LOC100128239,LOC100505474,LOC100506023,LOC101927134,LOC101927587,LOC101928174,LOC101928663,LOC101928782,LOC283177,LRRC4B,MAPT,MED27,MIR548AJ2,MKL1,MSI2,MSRA,MYBPC2,MYO1E,NGEF,NPTX1,NXPH1,PARD6G-AS1,PARP8,PCDH17,PFDN1,PRICKLE2,PSD3,RAB3C,RABGAP1L,RGS6,RNU6-28P,TP53BP1,ROBO1,SAMD3,SATB2,SDCCAG8,SFXN2,SFXN5,SGK223,SHANK3,SMG1P5,SMG6,SNX19,SREBF2,TBC1D5,TCF4,TDG,TDRD9,THAP8,TMEM131,TMEM192,TMEM56,TMTC1,TRPC4,TSNARE1,TTC7B,TUSC5,TXNRD1,WBP2NL,WNT3,ZNF365, |

| | | |
|---|---|---|
| 2 | 28 | ADAMTSL3,AKT3,SDCCAG8,ALPK3,ANKRD11,ANKRD44,ANKS1B,ARHGAP27, ASH2L,ASPG,BBX,BRE,CACNA1C,CACNB2,CAMKK2,CD4,CKAP5,CPEB1,CPNE8,CRHR1,DFNA5,DPEP1,DPP4,EFCAB6,EFNA5,EPPK1,EYS,FES,FGFR1,GNN,GRM1,GRM8,HIVEP1,HSP90B1,IQCE,KANSL1,KCTD18,KIAA1549,KLF12,KLHL20,LHFPL3,LHFPL3-AS2,LINC00051,LIPC,LOC100505474,LOC101927560,LOC101927587,MIR548AP,LOC101928174,LOC101928663,LOC101928782,LOC101929567,LOC101929690,LOC729970,LRRC16A,LRRC4B,LSM1,MAN2A2,MAPT,MEF2C,MEF2C-AS1,MIR548AJ2,MIR548AP,TTLL7,MSI2,MYBPC3,NALCN,NDST3,NEURL1,NGEF,NLRP1,NXPH1,OPCML,P2RX4,PFDN1,PLCH2,PLCL1,PLCL2,PRICKLE2,PSMC3,RABGAP1L,RBFOX1,RELA,RERE,RILPL2,SCGN,SESN2,SFXN2,SGCD,SLC39A8,SMPD3,SORCS3,ST3GAL3,TDG,TMEM192,TMEM56,TMTC1,TNN,TRPM6,TTC7B,ULK2,WNT3,ZNF71, |
| 2 | 29 | CPNE8,HCN1,LOC101928663,LRRC16A,PARP8,PDE4B,SCGN,SLC4A10, |
| 2 | 30 | ALPK3,GRAMD1B,MSANTD2,OR8B3,SIAE,SLC4A10,UBE2Q2P1, |
| 2 | 31 | CTB-12O2.1,MIR548AJ2,SLC4A10, |
| 2 | 32 | MFHAS1,MSRA,SGK223, |
| 2 | 35 | GRIN2A, |
| 3 | 2 | ACD,ARHGAP27,ARHGAP40,BBX,C19orf35,CACNB2,CDK10,CHKB-AS1,CHRNA2,CPNE7,CRHR1,CRK,CSMD1,CTB-12O2.1,CTCF,DBNDD1,DEF8,DNAH11,DOCK3,DPEP1,DPYD,GAK,GALNT10,GBF1,GPM6A,GRM2,GRM3,HEATR5B,HIVEP1,KANSL1,KIAA1324L,KLHL20,LAG3,LINC00635,LOC101927560,LOC101927587,MIR548AP, LOC101927641,LOC101928663,LOC101928782,LOC101928855,RPTOR,LOC283335,LRRC36,LTBP3,MAPT,MAPT-AS1,MIR548AJ2,MIR548AP,TTLL7,MLXIP,MSI2,NDRG4,NDUFAF7,NGEF,NRXN3,NSF,NSUN6,NXPH1,OASL,OPCML,P2RX7,PARD6G-AS1,PCBP2,PRDM7,PRKD1,PUS7,RABGAP1L,RELA,ROBO1,RORB,RPARP-AS1,RPTOR,SCGN,SGCD,SLC28A1,SLC47A1,SLC47A1,SNORA59A,SNORA59B,SMPD3,SNX8,SORCS3,SPATS2L,SPHKAP,SPIRE2,SRPK2,TDRD9,TMEM180,TMEM200A,TRPM6,TTC7B,VIT,WBSCR17,WNT3, |
| 3 | 3 | ABCB1,ABCB4,ABHD14A,ABHD14A-ACY1,ACD,ADAMTSL3,AIG1,AKAP6,ALG14,ANKRD11,ARFGAP2,ARHGAP40,BAI3,BCL11A,C3orf62,CA8,CACNA1S,CACNB2,CALN1,CAMTA1,CCDC134,CHRNA2,CPNE8,CPT1C,CRHR1,CTB-12O2.1,CYP7B1,DNAH11,DPYD,DUS2,ELFN1,EPPK1,FAM65B,FANCL,GARNL3,GAS8,GIGYF2,GLTP,GMIP,GNAI1,GPM6A,IMMP2L,LRRN3,KANSL1,KCNB1,KIAA1324L,KIAA1549, LHFPL3,LIN28B,LINC01122,LIPC,LOC101927134,LOC101928174,LOC101928882, LOC101929690,LOC339529,LRRC16A,LTBP3,LYSMD1,TNFAIP8L2-SCNM1,MAN2A1,MAPT,MEI1,MSI2,MSRA,NGEF,NSUN6,PARD6G,PPP1R16B,PRDM7,PRICKLE2,PRKD1,PRKD3,PRUNE,PSMC3,PTK2B,QPCT,RAB3C,RABGAP1L,RBFADN,RBFOX1,RELA,ROBO1,SAMD3,SATB2,SLC47A1,SMPD3,SORCS3,SOX2-OT,SPATS2L,TACC2,TBC1D5,TMEM192,TTYH3,TUSC5,UBD,VIT,WNT3,ZBTB43, |

| | | |
|---|---|---|
| 3 | 4 | ADAM22,ALG14,ANKRD44,ANKRD44,ANKRD44-IT1,ATP2A2,BCL11A,C3orf62, CA6,CACNA1D,CACNA1S,CACNA2D2,CNTN4,CORO7,CORO7-PAM16,CPEB1,CPNE7,CPNE8,CTB-12O2.1,DCC,DLX2,DPEP1,EXOC4,FAM120A,FANCL,FCGRT,FRMD5,GABBR2,GAPT,GNL3,GPN1,GPX6,GRM2,GRM3,IQCE,KCNB1,KCNJ3,KIAA1324L,KLF12,LHFPL3,LHFPL3,LHFPL3-AS2,LINC00635,LINC01122,LOC100505474,LOC100506023,LOC101927134,LOC101928663,LOC101928882,LOC101929567,LRRC16A,LRRC4B,MAPT,MFHAS1,MSI2,MSRA, MVP,NCAM1,PCBP2,PFDN1,PFKFB4,PLCL2,POC1B,PRDM7,PRR13,RAB3C,RBMS3,RELA,ROBO1,SCGN,SCRIB,SH3RF1,SLC4A10,SPATS2L,TMEM180,TMEM56,TRIM31,UBD,ULK2,WSCD2,ZNF71, |
| 3 | 5 | ASPG,ATP2A2,BAI3,BTN2A1,CACNB2,CALCOCO1,CALN1,CLCN3,CPNE7, CTB-12O2.1,DBNDD1,DEF8,DFNA5,DOCK3,DPEP1,EIF2AK2,EXOC4,FAM65B,GABBR2,GLB1L2,GLTP,GNL3,GPR98,GRIN2A,GRM2,GRM3,IQCF6,KCNJ3,KIAA1324L,KIAA1549,KLHL20, LHFPL3,LHFPL3-AS2,LINC00635,LIPC,LOC101927134,LOC101928663,LOC101928855,RPTOR,LOC283177,LOC388906,LRP1,LRRC16A,LTBP3,MANBA,MAPT,MEI1,MIR548AJ2,MVP,NDST3,NDUFAF7,NEBL,NLRP1,NRXN3,NSF,NSUN6,NXPH1,PCDH17,PCGF3,PFDN1,PFKFB4,PLEKHG5,POLD1,PTK2B,QPCT,RABGAP1L,RASSF1,RBFOX1,RELA,RGS6,RORB,RPTOR,SCGN, SFXN2,SGCD,SLC28A1,SLC45A1,SMPD3,SPIRE2,SREBF2,TACC2,TDRD9,TIE1,TMEM89, TRPM6,TTC7B,VIT,WSCD2,YWHAE,ZBTB43, |
| 3 | 7 | ACD,AMZ1,ANKRD11,ANKS1B,ANXA9,ASTN1,BAI3,C3orf62,CACNB2,CCDC63, CDH13,CLCN3,CNTN4,CRHR1,CSMD1,DBNDD1,DEF8,DPYD,EXOC4,FAM65B,GABBR2, GAS8,GMIP,GPR98,GRIN2A,IMMP2L,IQCF6,ITPKB,KANSL1,KCNJ3,LAG3,LGR4,LHFPL3,LHFPL3,LHFPL3-AS2,LINC00558,LOC101927539,MSI2,LOC101927560,LOC101927587,MIR548AP,LOC101927587,LOC101927641,LOC101928663,LOC283177,LRRC16A,MAPT,MEF2C-AS1,MIR548AP,TTLL7,MSI2,MYO19,NALCN,NEGR1,NLRP1,NSF,ORC5,P2RX7,PAK6,PCDHA1,PCDHA2,PCDHA3,PEX5L,PLCL1,POLD1,PRDM7,PSMD6,PSMD6-AS2,PTK2B,QPCT,RAB3C,RASSF1,RBKS,RERE,RIMS1,ROBO1,SAMD3,SCGN,SESN2,SGCD,SMPD3,SOX2-OT,SPIRE2,SREBF2,TCF4,TMEM180,TMEM56,TMEM56-RWDD3,TMTC1,TRPM6,TRPV4,UBE2D3,WBSCR17,WNT3,WSCD2, |
| 3 | 8 | ABHD2,AC093375.1,ACD,AKAP6,AKT3,AMZ1,BCL11A,BCL11B,C1orf106,C1orf210, CACNA1C,CACNA2D2,CACNB2,CALN1,CCDC63,CDH13,CDK10,CHRNA2,CKAP5,CPEB1,CPNE7,CTDP1,DBNDD1,DCC,DEF8,DPEP1,DPYD,EIF2AK2,FOXO3,FTCDNL1,GALNT10,GARNL3,GLTP,GNN,GPM6A,GPX6,GRAMD1B,HOMER2,IQCE,KIAA0319L,KIAA1324L,KLF12,LINC01122,LOC100506023,LOC100507283,LOC101926964,LOC101927587,LOC101928855,RPTOR,LOC101929690,LOC283177,LOC283335,LRRC16A,LRRC4B,LUZP2,MAD1L1,MAPK15,MAPT,MIR548AI,MIR548AJ2,MSI2,MSRA,MYO1E,NALCN,NRXN3,NSUN6, NUP210L,NXPH1,OASL,OSBPL3,P2RX7,PARP8,PCDHB9,PLCH2,PRKD3,PTK7,RAB3C,RABGAP1L,RELA,ROBO1,RPP30,RPTOR,SCGN,SDCCAG8,SGCD,SLC25A12,SLC45A1,SMIM17,SMPD3,SORCS3,SPATS2L,SPIRE2,SRPK2,STAG1,TACC2,TDG,TMEM131,TMEM180,TNN,TTC7B,VPS72,WNT3,ZNF365, |

| | | |
|---|---|---|
| 3 | 9 | ACD,ADAMTSL3,AIG1,AMZ1,ARHGAP27,CACNA1D,CACNB2,CALCOCO1,CNTN4, CPNE8,CRHR1,CSMD1,CTB-12O2.1,CYP7B1,DCC,DEF8,DPP4,DPYD,EFTUD1P1,EIF2AK2,FAM65B,FOXO3,FTCDNL1, GAS8,GLB1L2,GNL3,GPX6,GRM2,HEATR5B,HOMER2,IGSF9B,IQCE,ITPKB,KANSL1,KCNH6,KCNJ3,LOC101927134,LOC101927641,LOC101928174,LOC101928663,LOC101928782,LOC283177,LRRC16A,MAPT,MAPT,MAPT-AS1,MEF2C-AS1,MIR548AJ2,MSI2,MSRA,NALCN,NGEF,NSF,NSUN6,ORC5,P2RX7,PARP8,PCBP2,PHGR1,POC1A,PRDM7,PTN,RASSF1,SATB2,SCGN,SHANK3,SLC4A10,SMPD3,TANC2,TCF4, TDRD9,TMEM192,TMTC1,TRPV4,TTC7B,TTYH3,UBD,ULK2,WNT3, |
| 3 | 11 | ADAM22,AIG1,ANKRD11,ANKRD44,ANKS1B,BANK1,BCL11A,BCL11B,BRE, C14orf23,C1orf106,CA8,CACNA1C,CACNA1D,CACNA1S,CACNB2,CALN1,CD4,CNTN4, CSMD1,CTB-12O2.1,CYP7B1,DCC,DLX2,DOCK3,DPYD,DPYD-AS1,EPB42,EXOC4,FAM65B,FAM83G,SLC5A10,FRMD5,GALNT10,GNL3,GRM2,HAT1,HOMER2,IGSF9B,KANSL1,KCNC2,KLF12,LHFPL3,LHFPL3,LHFPL3-AS2,LINC01122,LOC100128239,LOC100506023,LOC100507283,LOC101927134,LOC101927572,LOC283177,LOC283335,LRRC16A,LRRC4B,MAD1L1,MANBA,MEI1,MFHAS1,MGAT3,MGRN1,MIR548AJ2,MIR548AP,TTLL7,MOB3A,NEGR1,OSBPL3,PDIA3,PFKFB4,PLCL2,PRICKLE2,PTGIS,PTK2B,RASSF1,RNU6-28P,TP53BP1,ROBO1,RORB,RPP30,RPTOR,SAMD3,SCGN,SDCCAG8,SFXN5,SGCD,SLC28A1,SLC45A1,SLC4A10,SLC5A10,SMG6,SNX8,SORCS3,SPHKAP,TBC1D15,TBC1D5,TMEM192,TMEM56,TMEM89,TNN,TRIM31,TRPM6,TRPV4,TTC7B,WBSCR17,WNT3,ZNF565,ZNF71, |
| 3 | 15 | AKAP6,AMZ1,ANKRD11,BAI3,C12orf43,C1orf95,CA8,CACNA1C,CACNA1D, CACNB2,CAMKK2,CAMTA1,CDC23,CHKB-AS1,CNTN4,CPNE7,CTDP1,DBNDD1,DCC,DEF8,DPEP1,DPYD,DPYD,DPYD-AS1,EFNA5,EXOC4,FAM65B,FANCL,FTCDNL1,GAS8,GLI1,GNE,HIVEP1,KCNB1,KCNJ3,KIF21B,KLF12,KLHL2,LAG3,LINC01122,LIPC,LOC100128239,LOC100505474,LOC101927587,LOC101927641,LOC646522,LRP1,LYSMD1,TNFAIP8L2-SCNM1,MAD1L1,MAN2A1,MED27,MEF2C,MEF2C-AS1,NEURL1,NXPH1,OPCML,OPRD1,PARD6G-AS1,PARP8,PCGEM1,PDE4B,PLCH2,PLCL2,PRKD1,PUS7,RBFOX1,RBMS3,RELA,RILPL1, RNF111,RORB,RPTOR,SCRIB,SGCD,SGSM2,SLC45A1,SNAP91,SORCS3,SPATS2L,SPHKAP,SPIRE2,SPPL3,STAB1,STAG1,TACC2,TBC1D5,TMEM192,TRPM6,TTC7B,TTYH3,TUSC5,WDR62,WDR66,YWHAE,ZNF365, |

| | | |
|---|---|---|
| 3 | 17 | ABHD14A,ABHD14A-ACY1,ACTC1,LOC101928174,AKT3,ALPK3,AMZ1,ARFGAP2, ARHGAP40, BBX,C12orf43,C1orf106,CA6,CACNB2,CALN1,CNTN4,CRHR1,CTB-12O2.1,CYP7B1,DFNA5,DOCK4,DPEP1,DPP4,DPYD,EFHD1,EFNA5,EIF2AK2,EYS,FAM65B,FGFR1,GAS8,GPM6A,HEATR5B,IL20RB,IMMP2L,IMMP2L,LRRN3,KANSL1,KCNB1,KCNJ3,KCTD18,KIAA1549,KLRG2,LGR4,LIMA1,LINC00635,LOC100506023,LOC101927134,LOC101927587,LOC101927641,LOC101928174,LOC101928782,LOC101929690,LOC646522,LRRC16A,MAPT,MIR548AI,MSRA,NCAM1,NEURL1,NXPH1,OASL,OSBPL3,P2RX7, PCGEM1,PDIA3,PRICKLE2,PTPRD,RBFOX1,RELA,RNU6-28P,TP53BP1,ROBO1,RORB,RPTOR,SAMD3,SCGN,SHANK3,SLC39A8,SLC45A1,SLC47A1,SNORA59A,SNORA59B,SLC6A9,SMPD3,SNX8,TACC2,TAS1R1,TBC1D15,TBL1XR1,TCF4,TMTC1,TSNARE1,TTYH3,TXNRD1,ULK2,VIT,WNT3,ZNF184,ZNF365, |
| 3 | 21 | ABCB4,ACD,AIG1,ANKK1,ANKRD44,ANKS1B,BCL9,CACNA1D,CACNA1S, CAMKK2,CD4,CLIP1,CNTN4,CPNE8,CRHR1,CTB-12O2.1,DEF8,DRD2,FAM65B,FANCL,GIGYF2,GNL3,GPN1,GPR98,GPX6,GRAMD1B,GRIN2A,GRM1,GRM2,HOMER2,KCNH6,KLF12,KLHL2,KLHL20,LINC01122,LOC101926905,LOC101927134,LOC101927560,LOC101927587,MIR548AP,LOC101928663,LRRC16A,LRRC8A,LTBP3,MAPT,MIR548AI,MIR548AJ2,MMP16,MYO1E,NALCN,NGEF,NRXN3,NSF,NSUN6,NXPH1,OASL,OPCML,P2RX4,PARD6G-AS1,PCBP2,PCGEM1,PLCL2,POC1A,PRICKLE2,PUS7,RASSF1,RBFOX1,RBMS3,RBMS3,RBMS3-AS1,RELA,RERE,RGS6,RNF111,RPTOR,SCGN,SGCD,SGSM2,SH3GLB2,SLC4A10,SPATS2L,SREBF2,TANC2,TBC1D5,TMEM180,TMEM192,TRPC4,TSPAN16,UBD,WNT3,WSCD2,YWHAE, |
| 3 | 22 | ABCB1,ADAMTSL3,AKT3,ANKRD11,ANKS1B,ASPG,BBX,BCL9,BLM,C12orf43, C1orf106,CACNA1C,CACNA1D,CACNB2,CALN1,CD4,CDC23,CNTN4,CPEB1,CPNE7,CRK,CSMD1,DBNDD1,DFNA5,DOCK4,EXOC4,FAM65B,FAM73B,FES,FURIN,GARNL3,GAS8,GNN,GPX6,GRAMD1B,GRIN2A,HIVEP1,KCNJ3,LOC100506472,TAB1,LOC100507283,LOC101926964,LOC101927587,LOC101928174,LOC101928663,LOC101928855,RPTOR, LOC101929690,LRRC16A,MAD1L1,MAN2A2,MAPT,MEF2C,MEF2C-AS1,MEI1,MFHAS1,MSRA,MYBPC2,NALCN,NDST3,OSBPL3,P2RX4,PFDN1,PLCL1,PRICKLE2,PTGIS,PTK2B,PTK7,PTPRD,RABGAP1L,RPRD2,RPTOR,SAMD3,SCGN,SLC28A1,SMPD3,SNX8,SPHKAP,SPIRE2,TACC2,TCF4,TDG,TMEM192,TRPC4,TRPM6,TTC7B,TXNRD1,UBD,UTRN,ZNF71, |
| 3 | 23 | AC093375.1,ACD,AIG1,ANKRD11,ATP2A2,BANK1,BBX,BTN2A1,C1orf210,CACNB2, CCDC63,CDH13,CKAP5,CNTN4,CPEB1,CRHR1,CRK,CSMD1,DBNDD1,DEF8,DFNA5,DPEP1,EFNA5,FAM65B,FRMD5,FTCDNL1,GALNT10,GBF1,GLTP,GNN,GPM6A,HOMER2,KCNJ3,KIAA1549,LINC00624,LOC100506023,LOC101243545,LOC101927273,LOC101927641,LOC101928663,LOC101929690,LOC283177,LOC283335,LRP1,LRRC16A,LRRC8A,MAPT,MED27,MIR548AJ2,NSUN6,PDIA3,PFDN1,PHGR1,PLCL2,PSMC3,RBFOX1,RBMS3,RGS6,RNU6-28P,TP53BP1,SAMD3,SATB2,SCGN,SGCD,SLC28A1,SMIM17,SMPD3,SNX8,SPATS2L,SPHKAP,SPIRE2,ST3GAL3,TBC1D5,TIE1,TMTC1,TRPM6,TSNARE1,TTC7B,TTYH3,UTRN,ZNF184, |

| | | |
|---|---|---|
| 3 | 24 | ACD,AKAP6,ANKS1B,ARFGAP2,ARHGAP27,BAI3,BBX,C1orf106,CACNA1D,CACNA1I,CACNB2,CCDC39,CDH13,CDIP1,CKAP5,CNTN4,CPEB1,CPNE8,CRHR1,CTB-12O2.1,DEF8,DGKI,DNAH11,DOCK3,DOCK4,DPYD,DPYD,DPYD-AS1,EIF2AK2,EPPK1,EXOC4,FAM120A,FANCL,FES,FOXO3,GALNT10,GATAD2A,GNL3,GOLPH3L,GPM6A,HEATR5B,HOMER2,IGSF9B,KANSL1,KCNC2,KIF21B,KLF12,KLHL20,LACE1,LGR4,LHFPL3,LHFPL3-AS2,LINC01122,LOC101926964,LOC101927560,LOC101927587,MIR548AP,LOC101927587,LOC101927641,LOC101929567,LOC101929690,LOC283177,LRRC16A,LRRC4B,LTBP3,MAN2A1,MAN2A2,MAPT,MEF2C,MEF2C-AS1,MSI2,MSRA,MYO1E,NALCN,NDRG4,NDUFAF7,NEBL,NGEF,NWD2,OASL,PAK6,PDE4B,PDIA3,PHF2,PLCH2,PPP1R16B,PRDM7,PRICKLE2,PRKD1,PRKD3,PSD3,PTK2B,QPCT,RAB3D,RABGAP1L,ROBO1,RORB,RPTOR,SMG6,SMPD3,SORCS3,SPATS2L,SPIRE2,SRPK2,ST3GAL3,SULT4A1,TACC2,TBL1XR1,TMEM192,TMTC1,TUSC5,VIT,WNT3,WSCD2,ZNF365, |
| 3 | 28 | ADAMTSL3,ALPK3,CACNB2,CPEB1,CRHR1,CTB-12O2.1,DEF8,EFTUD1,EFTUD1P1,HOMER2,KANSL1,KIAA1549,LINC00635,LOC101928663,LRRC16A,MAPT,NSF,SCGN,SLC28A1,SPIRE2,TM6SF1,TMEM192,UBE2Q2P1,WDR73,WNT3,ZNF804A, |
| 3 | 30 | MFHAS1,MSRA,SGK223,TNKS, |
| 3 | 31 | GRAMD1B,MIR548AJ2,MSANTD2,OR8B3,PFDN1,RANGAP1,SIAE, |
| 4 | 2 | ABCD2,AC093375.1,ACD,ADAMTSL3,ALPK3,ANKRD11,C12orf79,C16orf96,C1orf106,CACNA2D2,CALN1,CDIP1,CORO7,CORO7-PAM16,CPEB1,CPNE7,CPNE8,CTB-12O2.1,DBNDD1,DEF8,DFNA5,DPEP1,EFTUD1P1,FTCDNL1,GPATCH11,GPM6A,GRM3,HOMER2,IQCF6,KIAA1549,KIF21B,KLHL20,KLRG2,LINC01122,LOC100506023,LOC283177,LRRC36,LUZP2,MIR548AI,NDUFAF7,NRXN3,NXPH1,OR8B3,PDE4B,PLCB2,PLEKHG5,PPP1R16B,PRDM7,QPCT,RANGAP1,RGS6,RIMS1,ROBO1,SDCCAG8,SESN2,SHANK3,SLC28A1,SLC4A10,SMPD3,SPIRE2,TMEM192,TMEM56,TMEM56-RWDD3,TNN,TTC7B,TTYH3,UBE2D3,UBE2Q2P1,VIT, |
| 4 | 9 | ABCB1,ACTC1,LOC101928174,ADAT2,ALG14,ALPK3,BBX,BCL11B,BRE,C12orf79,C1orf95,CA8,CACNB2,CALCOCO1,CALN1,CAMTA1,CDC25C,CDH13,CNNM4,CNTN4,CORO7,CORO7-PAM16,CPEB1,CPNE8,CSMD1,DOCK3,DPEP1,DPYD,DPYD-AS1,EFHD1,ELFN1,EPB42,FAM124B,FAM65B,GAS8,GNL3,GPM6A,GPN1,GRM8,HIVEP1,KCNB1,KLHL20,KLRG2,LIMA1,LINC00635,LINC01088,LINC01305,LOC101927572,LOC101928174,LRRC16A,MAN2A1,MAPT,MEF2C,MEI1,MGAT3,MIEF1,MIR548AP,TTLL7,MSI2,MYO1E,NGEF,OPCML,PAK6,PEX5L,PFDN1,PPP2R2A,PRICKLE2,PRKD1,PRUNE,PSD3,PSMC3,PTBP2,PUS7,RAB3C,RABGAP1L,RBMS3,RERE,RIMS1,SAMD3,SGCD,SGSM2,SHANK3,SLC5A10,SLC9C2,SMPD3,SOX2-OT,SPATS2L,SPIRE2,ST3GAL3,TACC2,TBL1XR1,TNN,TRPC4,TTC7B,UBE2Q2P1,UTRN,WBSCR17,ZAP70,ZDHHC12, |

| | | |
|---|---|---|
| 4 | 15 | ACD,ADAM22,ADAMTSL3,AIG1,ANKRD11,BCL11B,C1orf210,C9orf129,CACNA1C,CALN1,CDH13,CKAP5,CNTN4,CPNE8,CRHR1,CSMD1,DBNDD1,DEF8,DFNA5,DOCK3,DPEP1,DPH1,DUS2,EFCAB6,EFTUD1P1,ELFN1,EYS,FCGRT,FTCDNL1,FYN,GABBR2,GALNT10,GLB1L2,GOLPH3L,GPR98,GRM2,HCG17,HCG18,HSP90B1,KCNB1,KCNJ3,LHFPL3,LHFPL3-AS2,LINC01122,LOC100506472,TAB1,LOC101927572,LOC101928782, LOC101928882,LOC101929567,LOC283177,LOC646522,LRRC4B,LRRC8A,MED27,MFHAS1,MSI2,MSRA,MYBPC2,NUP210L,OLA1,OSBPL3,PDCD11,PDE4B,PLEKHG5,POLD1, PRDM7,PRICKLE2,PRKD1,PSMC3,PSMD6,PSMD6-AS2,PTGIS,RAB3C,RANGAP1,RORB,RPTOR,SATB2,SGK223,SHANK3,SLC25A12,SLC28A1,SLC38A7,SLC4A10,SLC5A10,SMG6,SMPD3,SORCS3,SOX2-OT,SPIRE2,SREBF2,SULT4A1,TIE1,TMEM131,TMTC1,TNN,TRPC4,TTC7B,VIT,WBSCR17,WNT3,WSCD2,ZDHHC12,ZNF804A, |
| 4 | 18 | ACTC1,LOC101928174,AIG1,AKAP6,BAG4,BCL11B,BRE,C15orf56,PAK6, C1orf106,CACNB2,CALN1,CCDC63,CDH13,CLIP1,CORO7,CORO7-PAM16,CPT1C,CRHR1,CTB-12O2.1,DPYD,EFNA5,ENOX1,EPPK1,ESAM,EYS,FCGRT,FGFR1,FTCDNL1,GAK,GAS8,GLB1L2,GLI1,GOLPH3L,GPN3,GRAMD1B,GRIN2A,GRM2,HCG17,HCG18,HIP1R,HIVEP1,KLF12,LHFPL3,LOC101927134,LOC101927587,LOC101927641,LOC101928174,LRRC16A, LRRC36,LRRC4B,LSM1,MAD1L1,MAPT,MED27,MEF2B,MEF2BNB-MEF2B,MEF2BNB,MEF2BNB-MEF2B,MEF2BNB-MEF2B,MEF2C,MGRN1,MIR548AJ2,MSI2,MYO19,MYO1C,NALCN,NDRG4,NGEF,NLRP1, NWD2,OPCML,P2RX7,PCDH17,PFDN1,PFKFB4,PLCB2,PLCL1,PRDM7,PRICKLE2,PRKD1,PSD3,PTBP2,PTGIS,QPCT,RBFOX1,RBMS3,RELA,RILPL1,ROBO1,SDR9C7,SF3B1,SGCD, SHANK3,SLC27A3,SLC47A2,SLC4A10,SMPD3,SNX8,SOX2-OT,SPIRE2,TACC2,TMTC1,TRPM6,TTC7B,UTRN,VIT,WDR66,WNT3,ZNF365, |
| 4 | 23 | ACD,ACR,ADAMTSL3,AKAP6,ALPK3,AMZ1,ARHGAP27,BBX,C12orf79, C14orf23,C1orf95,CA8,CACNB2,CAMKK2,CAMTA1,CCDC63,CD4,CDH13,CDIP1,CNTN4,CPNE8,CRHR1,CSMD1,CTB-12O2.1,CTCF,DOCK6,EFTUD1P1,ENOX1,FUT9,GNN,GPM6A,GPR98,GRM3,HAT1,HEATR5B,HIVEP1,HSP90B1,IQCE,ITPKB,KANSL1,KCNB1,KIAA1324L,KLHL20,LIMA1,LOC100506023,LOC101926905,LOC101927229,LOC101927587,LOC101928855,RPTOR,LRRC16A,LRRC36,MAD1L1,MAPT,ME1,MIR548AJ2,MSRA,NALCN,NCAM1,NWD2,OPCML,OPRD1,PCGEM1,PDE4B,PDIA3,PFDN1,PLCL2,PPP1R16B,PRDM7,QPCT,RABGAP1L,RBFADN,RBFOX1,RGS6,RIMS1,RPTOR,SGCD,SLC39A8,SLC4A10,SMPD3,SNX8,ST3GAL3,STAG1,SULT4A1,TCF4,TMTC1,TRIM31,TRPC4,TUSC5,UBE2D3,UBE2Q2P1,WNT3,WSCD2,ZEB2, |
| 4 | 30 | ADAMTSL3,ALPK3,CPNE8,CTB-12O2.1,EFTUD1P1,HCN1,LOC100506023, LOC101928663,LRRC16A,OR8B3,PARP8,SCGN,SLC4A10,UBE2Q2P1, |
| 4 | 31 | ADAMTSL3,ALPK3,CPNE8,CTB-12O2.1,KLHL20,LOC100506023,MMP16, RABGAP1L,SLC28A1,UBE2Q2P1, |
| 4 | 35 | GRIN2A, |
