## Supplemental Table 2 for "Dynamic fusion of genomics and functional network connectivity in UK biobank reveals schizophrenia-related SNP manifolds"

| |
|---|
| Table S2: Anatomical labels of the top connectivity pairs for individual SZ-relevant components. |
| state1_jica2 |
| "Right inferior frontal gyrus ([R IFG], 61)" - "Superior temporal gyrus ([STG], 21)" |
| "Right inferior frontal gyrus ([R IFG], 61)" - "Middle temporal gyrus ([MTG], 56)" |
| "Right inferior frontal gyrus ([R IFG], 61)" - "Postcentral gyrus ([PoCG], 3)" |
| "Right inferior frontal gyrus ([R IFG], 61)" - "Left postcentral gyrus ([L PoCG], 9)" |
| "Right inferior frontal gyrus ([R IFG], 61)" - "Paracentral lobule ([ParaCL], 2)" |
| "Right inferior frontal gyrus ([R IFG], 61)" - "Right postcentral gyrus ([R PoCG], 11)" |
| "Postcentral gyrus ([PoCG], 72)" - "Superior parietal lobule ([SPL], 27)" |
| "Right inferior frontal gyrus ([R IFG], 61)" - "Superior parietal lobule ([SPL], 27)" |
| "Postcentral gyrus ([PoCG], 72)" - "Paracentral lobule ([ParaCL], 54)" |
| "Right inferior frontal gyrus ([R IFG], 61)" - "Paracentral lobule ([ParaCL], 54)" |
| "Postcentral gyrus ([PoCG], 72)" - "Precentral gyrus ([PreCG], 66)" |
| "Right inferior frontal gyrus ([R IFG], 61)" - "Precentral gyrus ([PreCG], 66)" |
| "Right inferior frontal gyrus ([R IFG], 61)" - "Superior parietal lobule ([SPL], 80)" |
| "Inferior parietal lobule ([IPL], 68)" - "Postcentral gyrus ([PoCG], 72)" |
| "Superior medial frontal gyrus ([SMFG], 43)" - "Postcentral gyrus ([PoCG], 72)" |
| "Superior frontal gyrus ([SFG], 96)" - "Postcentral gyrus ([PoCG], 72)" |
| "Left inferior parietal lobue ([L IPL], 81)" - "Postcentral gyrus ([PoCG], 72)" |
| "Posterior cingulate cortex ([PCC], 94)" - "Postcentral gyrus ([PoCG], 72)" |
| "Right inferior frontal gyrus ([R IFG], 61)" - "Right middle occipital gyrus ([R MOG], 12)" |
| "Right inferior frontal gyrus ([R IFG], 61)" - "Inferior occipital gyrus ([IOG], 20)" |
| "Right inferior frontal gyrus ([R IFG], 61)" - "Inferior parietal lobule ([IPL], 68)" |
| "Left inferior parietal lobue ([L IPL], 81)" - "Inferior parietal lobule ([IPL], 68)" |
| "Right inferior frontal gyrus ([R IFG], 61)" - "Superior medial frontal gyrus ([SMFG], 43)" |
| "Left inferior parietal lobue ([R IPL], 79)" - "Right inferior frontal gyrus ([R IFG], 61)" |
| "Superior frontal gyrus ([SFG], 96)" - "Right inferior frontal gyrus ([R IFG], 61)" |
| "Left inferior parietal lobue ([L IPL], 81)" - "Right inferior frontal gyrus ([R IFG], 61)" |
| "Middle cingulate cortex ([MCC], 37)" - "Right inferior frontal gyrus ([R IFG], 61)" |
| Precuneus (40) - "Right inferior frontal gyrus ([R IFG], 61)" |
| "Anterior cingulate cortex ([ACC], 23)" - "Right inferior frontal gyrus ([R IFG], 61)" |
| "Posterior cingulate cortex ([PCC], 71)" - "Right inferior frontal gyrus ([R IFG], 61)" |
| "Anterior cingulate cortex ([ACC], 17)" - "Right inferior frontal gyrus ([R IFG], 61)" |
| "Posterior cingulate cortex ([PCC], 94)" - "Right inferior frontal gyrus ([R IFG], 61)" |
| "Cerebellum ([CB], 18)" - "Right inferior frontal gyrus ([R IFG], 61)" |
| "Cerebellum ([CB], 7)" - "Right inferior frontal gyrus ([R IFG], 61)" |
| state1_jica3 |
| "Middle frontal gyrus ([MiFG], 38)" - Caudate (69) |
| "Middle frontal gyrus ([MiFG], 38)" - "Superior temporal gyrus ([STG], 21)" |
| "Middle frontal gyrus ([MiFG], 38)" - "Middle temporal gyrus ([MTG], 56)" |
| "Middle frontal gyrus ([MiFG], 38)" - "Postcentral gyrus ([PoCG], 3)" |
| "Middle frontal gyrus ([MiFG], 38)" - "Left postcentral gyrus ([L PoCG], 9)" |
| "Middle frontal gyrus ([MiFG], 38)" - "Right postcentral gyrus ([R PoCG], 11)" |
| "Middle frontal gyrus ([MiFG], 38)" - "Superior parietal lobule ([SPL], 27)" |

| |
|---|
| "Middle frontal gyrus ([MiFG], 38)" - "Paracentral lobule ([ParaCL], 54)" |
| "Middle frontal gyrus ([MiFG], 38)" - "Precentral gyrus ([PreCG], 66)" |
| "Middle frontal gyrus ([MiFG], 38)" - "Superior parietal lobule ([SPL], 80)" |
| "Middle frontal gyrus ([MiFG], 38)" - "Postcentral gyrus ([PoCG], 72)" |
| "Middle frontal gyrus ([MiFG], 38)" - "Middle temporal gyrus ([MTG], 77)" |
| "Middle frontal gyrus ([MiFG], 38)" - "Inferior parietal lobule ([IPL], 68)" |
| "Middle frontal gyrus ([MiFG], 38)" - Insula (33) |
| "Middle frontal gyrus ([MiFG], 38)" - "Superior medial frontal gyrus ([SMFG], 43)" |
| "Middle frontal gyrus ([MiFG], 38)" - "Left inferior parietal lobue ([R IPL], 79)" |
| "Middle frontal gyrus ([MiFG], 38)" - "Superior frontal gyrus ([SFG], 96)" |
| "Middle frontal gyrus ([MiFG], 38)" - "Middle frontal gyrus ([MiFG], 88)" |
| "Middle frontal gyrus ([MiFG], 38)" - "Hippocampus ([HiPP], 48)" |
| "Middle frontal gyrus ([MiFG], 38)" - "Left inferior parietal lobue ([L IPL], 81)" |
| "Middle frontal gyrus ([MiFG], 38)" - "Middle cingulate cortex ([MCC], 37)" |
| "Hippocampus ([HiPP], 83)" - "Middle frontal gyrus ([MiFG], 38)" |
| Precuneus (40) - "Middle frontal gyrus ([MiFG], 38)" |
| "Anterior cingulate cortex ([ACC], 23)" - "Middle frontal gyrus ([MiFG], 38)" |
| "Posterior cingulate cortex ([PCC], 71)" - "Middle frontal gyrus ([MiFG], 38)" |
| "Anterior cingulate cortex ([ACC], 17)" - "Middle frontal gyrus ([MiFG], 38)" |
| "Posterior cingulate cortex ([PCC], 94)" - "Middle frontal gyrus ([MiFG], 38)" |
| "Cerebellum ([CB], 18)" - "Middle frontal gyrus ([MiFG], 38)" |
| "Cerebellum ([CB], 4)" - "Middle frontal gyrus ([MiFG], 38)" |
| "Cerebellum ([CB], 7)" - "Middle frontal gyrus ([MiFG], 38)" |
| state1_jica4 |
| Insula (33) - "Superior temporal gyrus ([STG], 21)" |
| Insula (33) - "Middle temporal gyrus ([MTG], 56)" |
| "Right inferior frontal gyrus ([R IFG], 61)" - "Middle temporal gyrus ([MTG], 56)" |
| "Supplementary motor area ([SMA], 84)" - "Middle temporal gyrus ([MTG], 56)" |
| Insula (33) - "Postcentral gyrus ([PoCG], 3)" |
| Insula (33) - "Left postcentral gyrus ([L PoCG], 9)" |
| Insula (33) - "Right postcentral gyrus ([R PoCG], 11)" |
| Insula (33) - "Superior parietal lobule ([SPL], 27)" |
| Insula (33) - "Paracentral lobule ([ParaCL], 54)" |
| Insula (33) - "Precentral gyrus ([PreCG], 66)" |
| Insula (33) - "Superior parietal lobule ([SPL], 80)" |
| Insula (33) - "Postcentral gyrus ([PoCG], 72)" |
| Insula (33) - "Inferior parietal lobule ([IPL], 68)" |
| "Superior medial frontal gyrus ([SMFG], 43)" - Insula (33) |
| "Left inferior parietal lobue ([R IPL], 79)" - Insula (33) |
| "Superior frontal gyrus ([SFG], 96)" - Insula (33) |
| "Middle frontal gyrus ([MiFG], 88)" - Insula (33) |
| "Left inferior parietal lobue ([L IPL], 81)" - Insula (33) |
| "Middle cingulate cortex ([MCC], 37)" - Insula (33) |
| Precuneus (40) - Insula (33) |

| |
|---|
| "Anterior cingulate cortex ([ACC], 23)" - Insula (33) |
| "Posterior cingulate cortex ([PCC], 71)" - Insula (33) |
| "Anterior cingulate cortex ([ACC], 17)" - Insula (33) |
| "Posterior cingulate cortex ([PCC], 94)" - Insula (33) |
| "Cerebellum ([CB], 18)" - Insula (33) |
| "Left inferior parietal lobue ([R IPL], 79)" - "Right inferior frontal gyrus ([R IFG], 61)" |
| "Left inferior parietal lobue ([L IPL], 81)" - "Right inferior frontal gyrus ([R IFG], 61)" |
| "Posterior cingulate cortex ([PCC], 94)" - "Right inferior frontal gyrus ([R IFG], 61)" |
| "Supplementary motor area ([SMA], 84)" - "Left inferior parietal lobue ([R IPL], 79)" |
| "Superior frontal gyrus ([SFG], 96)" - "Supplementary motor area ([SMA], 84)" |
| "Posterior cingulate cortex ([PCC], 94)" - "Supplementary motor area ([SMA], 84)" |
| state1_jica6 |
| "Middle temporal gyrus ([MTG], 77)" - "Superior temporal gyrus ([STG], 21)" |
| "Middle temporal gyrus ([MTG], 77)" - "Middle temporal gyrus ([MTG], 56)" |
| "Middle temporal gyrus ([MTG], 77)" - "Postcentral gyrus ([PoCG], 3)" |
| "Middle temporal gyrus ([MTG], 77)" - "Left postcentral gyrus ([L PoCG], 9)" |
| "Middle temporal gyrus ([MTG], 77)" - "Right postcentral gyrus ([R PoCG], 11)" |
| "Middle temporal gyrus ([MTG], 77)" - "Superior parietal lobule ([SPL], 27)" |
| "Middle temporal gyrus ([MTG], 77)" - "Paracentral lobule ([ParaCL], 54)" |
| "Middle temporal gyrus ([MTG], 77)" - "Precentral gyrus ([PreCG], 66)" |
| "Middle temporal gyrus ([MTG], 77)" - "Superior parietal lobule ([SPL], 80)" |
| "Posterior cingulate cortex ([PCC], 94)" - "Superior parietal lobule ([SPL], 80)" |
| "Middle temporal gyrus ([MTG], 77)" - "Postcentral gyrus ([PoCG], 72)" |
| "Inferior parietal lobule ([IPL], 68)" - "Middle temporal gyrus ([MTG], 77)" |
| "Superior medial frontal gyrus ([SMFG], 43)" - "Middle temporal gyrus ([MTG], 77)" |
| "Left inferior parietal lobue ([R IPL], 79)" - "Middle temporal gyrus ([MTG], 77)" |
| "Superior frontal gyrus ([SFG], 96)" - "Middle temporal gyrus ([MTG], 77)" |
| "Middle frontal gyrus ([MiFG], 88)" - "Middle temporal gyrus ([MTG], 77)" |
| "Hippocampus ([HiPP], 48)" - "Middle temporal gyrus ([MTG], 77)" |
| "Left inferior parietal lobue ([L IPL], 81)" - "Middle temporal gyrus ([MTG], 77)" |
| "Hippocampus ([HiPP], 83)" - "Middle temporal gyrus ([MTG], 77)" |
| Precuneus (40) - "Middle temporal gyrus ([MTG], 77)" |
| "Anterior cingulate cortex ([ACC], 23)" - "Middle temporal gyrus ([MTG], 77)" |
| "Anterior cingulate cortex ([ACC], 17)" - "Middle temporal gyrus ([MTG], 77)" |
| "Posterior cingulate cortex ([PCC], 94)" - "Middle temporal gyrus ([MTG], 77)" |
| "Cerebellum ([CB], 18)" - "Middle temporal gyrus ([MTG], 77)" |
| state1_jica9 |
| "Superior temporal gyrus ([STG], 21)" - Subthalamus/hypothalamus (53) |
| "Middle temporal gyrus ([MTG], 56)" - Subthalamus/hypothalamus (53) |
| "Postcentral gyrus ([PoCG], 3)" - Subthalamus/hypothalamus (53) |
| "Left postcentral gyrus ([L PoCG], 9)" - Subthalamus/hypothalamus (53) |
| "Paracentral lobule ([ParaCL], 2)" - Subthalamus/hypothalamus (53) |
| "Right postcentral gyrus ([R PoCG], 11)" - Subthalamus/hypothalamus (53) |
| "Superior parietal lobule ([SPL], 27)" - Subthalamus/hypothalamus (53) |

| |
|---|
| "Paracentral lobule ([ParaCL], 54)" - Subthalamus/hypothalamus (53) |
| "Precentral gyrus ([PreCG], 66)" - Subthalamus/hypothalamus (53) |
| "Postcentral gyrus ([PoCG], 72)" - Subthalamus/hypothalamus (53) |
| "Inferior parietal lobule ([IPL], 68)" - Subthalamus/hypothalamus (53) |
| "Superior medial frontal gyrus ([SMFG], 43)" - Subthalamus/hypothalamus (53) |
| "Left inferior parietal lobue ([R IPL], 79)" - Subthalamus/hypothalamus (53) |
| "Superior frontal gyrus ([SFG], 96)" - Subthalamus/hypothalamus (53) |
| "Left inferior parietal lobue ([L IPL], 81)" - Subthalamus/hypothalamus (53) |
| Precuneus (40) - Subthalamus/hypothalamus (53) |
| "Anterior cingulate cortex ([ACC], 23)" - Subthalamus/hypothalamus (53) |
| "Posterior cingulate cortex ([PCC], 71)" - Subthalamus/hypothalamus (53) |
| "Posterior cingulate cortex ([PCC], 94)" - Subthalamus/hypothalamus (53) |
| "Cerebellum ([CB], 18)" - Subthalamus/hypothalamus (53) |
| "Superior temporal gyrus ([STG], 21)" - Thalamus (45) |
| "Middle temporal gyrus ([MTG], 56)" - Thalamus (45) |
| "Postcentral gyrus ([PoCG], 3)" - Thalamus (45) |
| "Left postcentral gyrus ([L PoCG], 9)" - Thalamus (45) |
| "Paracentral lobule ([ParaCL], 2)" - Thalamus (45) |
| "Right postcentral gyrus ([R PoCG], 11)" - Thalamus (45) |
| "Superior parietal lobule ([SPL], 27)" - Thalamus (45) |
| "Paracentral lobule ([ParaCL], 54)" - Thalamus (45) |
| "Precentral gyrus ([PreCG], 66)" - Thalamus (45) |
| "Superior parietal lobule ([SPL], 80)" - Thalamus (45) |
| "Postcentral gyrus ([PoCG], 72)" - Thalamus (45) |
| "Inferior parietal lobule ([IPL], 68)" - Thalamus (45) |
| "Superior medial frontal gyrus ([SMFG], 43)" - Thalamus (45) |
| "Left inferior parietal lobue ([R IPL], 79)" - Thalamus (45) |
| "Superior frontal gyrus ([SFG], 96)" - Thalamus (45) |
| "Left inferior parietal lobue ([L IPL], 81)" - Thalamus (45) |
| "Middle cingulate cortex ([MCC], 37)" - Thalamus (45) |
| "Middle frontal gyrus ([MiFG], 38)" - Thalamus (45) |
| Precuneus (40) - Thalamus (45) |
| "Anterior cingulate cortex ([ACC], 23)" - Thalamus (45) |
| "Posterior cingulate cortex ([PCC], 71)" - Thalamus (45) |
| "Posterior cingulate cortex ([PCC], 94)" - Thalamus (45) |
| "Cerebellum ([CB], 18)" - Thalamus (45) |
| "Cerebellum ([CB], 4)" - Thalamus (45) |
| "Cerebellum ([CB], 7)" - Thalamus (45) |
| state1_jica10 |
| "Inferior frontal gyrus ([IFG], 70)" - "Superior temporal gyrus ([STG], 21)" |
| "Inferior frontal gyrus ([IFG], 70)" - "Middle temporal gyrus ([MTG], 56)" |
| "Middle frontal gyrus ([MiFG], 55)" - "Middle temporal gyrus ([MTG], 56)" |
| "Supplementary motor area ([SMA], 84)" - "Middle temporal gyrus ([MTG], 56)" |
| "Cerebellum ([CB], 13)" - "Middle temporal gyrus ([MTG], 56)" |

| |
|---|
| "Inferior frontal gyrus ([IFG], 70)" - "Postcentral gyrus ([PoCG], 3)" |
| "Inferior frontal gyrus ([IFG], 70)" - "Left postcentral gyrus ([L PoCG], 9)" |
| "Middle frontal gyrus ([MiFG], 55)" - "Left postcentral gyrus ([L PoCG], 9)" |
| "Inferior frontal gyrus ([IFG], 70)" - "Right postcentral gyrus ([R PoCG], 11)" |
| "Middle frontal gyrus ([MiFG], 55)" - "Right postcentral gyrus ([R PoCG], 11)" |
| "Inferior frontal gyrus ([IFG], 70)" - "Superior parietal lobule ([SPL], 27)" |
| "Middle frontal gyrus ([MiFG], 55)" - "Superior parietal lobule ([SPL], 27)" |
| "Inferior frontal gyrus ([IFG], 70)" - "Paracentral lobule ([ParaCL], 54)" |
| "Inferior frontal gyrus ([IFG], 70)" - "Precentral gyrus ([PreCG], 66)" |
| "Middle frontal gyrus ([MiFG], 55)" - "Precentral gyrus ([PreCG], 66)" |
| "Inferior frontal gyrus ([IFG], 70)" - "Postcentral gyrus ([PoCG], 72)" |
| "Inferior frontal gyrus ([IFG], 70)" - "Inferior parietal lobule ([IPL], 68)" |
| "Inferior frontal gyrus ([IFG], 70)" - "Superior medial frontal gyrus ([SMFG], 43)" |
| "Middle frontal gyrus ([MiFG], 55)" - "Superior medial frontal gyrus ([SMFG], 43)" |
| "Left inferior parietal lobue ([R IPL], 79)" - "Inferior frontal gyrus ([IFG], 70)" |
| "Superior frontal gyrus ([SFG], 96)" - "Inferior frontal gyrus ([IFG], 70)" |
| "Left inferior parietal lobue ([L IPL], 81)" - "Inferior frontal gyrus ([IFG], 70)" |
| Precuneus (40) - "Inferior frontal gyrus ([IFG], 70)" |
| "Anterior cingulate cortex ([ACC], 23)" - "Inferior frontal gyrus ([IFG], 70)" |
| "Posterior cingulate cortex ([PCC], 71)" - "Inferior frontal gyrus ([IFG], 70)" |
| "Anterior cingulate cortex ([ACC], 17)" - "Inferior frontal gyrus ([IFG], 70)" |
| "Posterior cingulate cortex ([PCC], 94)" - "Inferior frontal gyrus ([IFG], 70)" |
| "Cerebellum ([CB], 18)" - "Inferior frontal gyrus ([IFG], 70)" |
| "Left inferior parietal lobue ([R IPL], 79)" - "Middle frontal gyrus ([MiFG], 55)" |
| "Superior frontal gyrus ([SFG], 96)" - "Middle frontal gyrus ([MiFG], 55)" |
| Precuneus (40) - "Middle frontal gyrus ([MiFG], 55)" |
| "Posterior cingulate cortex ([PCC], 94)" - "Middle frontal gyrus ([MiFG], 55)" |
| "Superior frontal gyrus ([SFG], 96)" - "Supplementary motor area ([SMA], 84)" |
| "Posterior cingulate cortex ([PCC], 94)" - "Supplementary motor area ([SMA], 84)" |
| "Middle frontal gyrus ([MiFG], 88)" - "Superior frontal gyrus ([SFG], 96)" |
| "Cerebellum ([CB], 13)" - "Superior frontal gyrus ([SFG], 96)" |
| "Posterior cingulate cortex ([PCC], 94)" - "Middle frontal gyrus ([MiFG], 88)" |
| "Cerebellum ([CB], 13)" - "Posterior cingulate cortex ([PCC], 94)" |
| state1_jica12 |
| Putamen (98) - Caudate (69) |
| Caudate (99) - Caudate (69) |
| "Superior temporal gyrus ([STG], 21)" - Putamen (98) |
| "Middle temporal gyrus ([MTG], 56)" - Putamen (98) |
| "Postcentral gyrus ([PoCG], 3)" - Putamen (98) |
| "Left postcentral gyrus ([L PoCG], 9)" - Putamen (98) |
| "Paracentral lobule ([ParaCL], 2)" - Putamen (98) |
| "Right postcentral gyrus ([R PoCG], 11)" - Putamen (98) |
| "Superior parietal lobule ([SPL], 27)" - Putamen (98) |
| "Paracentral lobule ([ParaCL], 54)" - Putamen (98) |

| |
|---|
| "Precentral gyrus ([PreCG], 66)" - Putamen (98) |
| "Superior parietal lobule ([SPL], 80)" - Putamen (98) |
| "Postcentral gyrus ([PoCG], 72)" - Putamen (98) |
| "Superior medial frontal gyrus ([SMFG], 43)" - Putamen (98) |
| "Left inferior parietal lobue ([R IPL], 79)" - Putamen (98) |
| "Superior frontal gyrus ([SFG], 96)" - Putamen (98) |
| "Left inferior parietal lobue ([L IPL], 81)" - Putamen (98) |
| "Hippocampus ([HiPP], 83)" - Putamen (98) |
| Precuneus (40) - Putamen (98) |
| "Anterior cingulate cortex ([ACC], 23)" - Putamen (98) |
| "Posterior cingulate cortex ([PCC], 71)" - Putamen (98) |
| "Anterior cingulate cortex ([ACC], 17)" - Putamen (98) |
| "Posterior cingulate cortex ([PCC], 94)" - Putamen (98) |
| "Cerebellum ([CB], 18)" - Putamen (98) |
| "Cerebellum ([CB], 4)" - Putamen (98) |
| "Cerebellum ([CB], 7)" - Putamen (98) |
| "Superior temporal gyrus ([STG], 21)" - Caudate (99) |
| "Middle temporal gyrus ([MTG], 56)" - Caudate (99) |
| "Postcentral gyrus ([PoCG], 3)" - Caudate (99) |
| "Left postcentral gyrus ([L PoCG], 9)" - Caudate (99) |
| "Paracentral lobule ([ParaCL], 2)" - Caudate (99) |
| "Right postcentral gyrus ([R PoCG], 11)" - Caudate (99) |
| "Superior parietal lobule ([SPL], 27)" - Caudate (99) |
| "Paracentral lobule ([ParaCL], 54)" - Caudate (99) |
| "Precentral gyrus ([PreCG], 66)" - Caudate (99) |
| "Superior parietal lobule ([SPL], 80)" - Caudate (99) |
| "Postcentral gyrus ([PoCG], 72)" - Caudate (99) |
| "Middle temporal gyrus ([MTG], 62)" - Caudate (99) |
| "Inferior parietal lobule ([IPL], 68)" - Caudate (99) |
| "Superior medial frontal gyrus ([SMFG], 43)" - Caudate (99) |
| "Left inferior parietal lobue ([R IPL], 79)" - Caudate (99) |
| "Superior frontal gyrus ([SFG], 96)" - Caudate (99) |
| "Middle frontal gyrus ([MiFG], 88)" - Caudate (99) |
| "Hippocampus ([HiPP], 48)" - Caudate (99) |
| "Left inferior parietal lobue ([L IPL], 81)" - Caudate (99) |
| "Middle cingulate cortex ([MCC], 37)" - Caudate (99) |
| "Hippocampus ([HiPP], 83)" - Caudate (99) |
| Precuneus (40) - Caudate (99) |
| "Anterior cingulate cortex ([ACC], 23)" - Caudate (99) |
| "Posterior cingulate cortex ([PCC], 71)" - Caudate (99) |
| "Anterior cingulate cortex ([ACC], 17)" - Caudate (99) |
| "Posterior cingulate cortex ([PCC], 94)" - Caudate (99) |
| "Cerebellum ([CB], 18)" - Caudate (99) |
| "Cerebellum ([CB], 4)" - Caudate (99) |

| |
|---|
| "Cerebellum ([CB], 7)" - Caudate (99) |
| state1_jica13 |
| "Cerebellum ([CB], 13)" - Caudate (69) |
| "Cerebellum ([CB], 13)" - "Superior temporal gyrus ([STG], 21)" |
| "Cerebellum ([CB], 7)" - "Superior temporal gyrus ([STG], 21)" |
| "Cerebellum ([CB], 13)" - "Middle temporal gyrus ([MTG], 56)" |
| "Cerebellum ([CB], 7)" - "Middle temporal gyrus ([MTG], 56)" |
| "Cerebellum ([CB], 13)" - "Postcentral gyrus ([PoCG], 3)" |
| "Cerebellum ([CB], 13)" - "Left postcentral gyrus ([L PoCG], 9)" |
| "Cerebellum ([CB], 7)" - "Left postcentral gyrus ([L PoCG], 9)" |
| "Cerebellum ([CB], 13)" - "Paracentral lobule ([ParaCL], 2)" |
| "Cerebellum ([CB], 13)" - "Right postcentral gyrus ([R PoCG], 11)" |
| "Cerebellum ([CB], 7)" - "Right postcentral gyrus ([R PoCG], 11)" |
| "Cerebellum ([CB], 13)" - "Superior parietal lobule ([SPL], 27)" |
| "Cerebellum ([CB], 7)" - "Superior parietal lobule ([SPL], 27)" |
| "Cerebellum ([CB], 13)" - "Paracentral lobule ([ParaCL], 54)" |
| "Cerebellum ([CB], 13)" - "Precentral gyrus ([PreCG], 66)" |
| "Cerebellum ([CB], 7)" - "Precentral gyrus ([PreCG], 66)" |
| "Cerebellum ([CB], 13)" - "Superior parietal lobule ([SPL], 80)" |
| "Cerebellum ([CB], 7)" - "Superior parietal lobule ([SPL], 80)" |
| "Cerebellum ([CB], 13)" - "Postcentral gyrus ([PoCG], 72)" |
| "Cerebellum ([CB], 7)" - "Postcentral gyrus ([PoCG], 72)" |
| "Cerebellum ([CB], 13)" - "Middle temporal gyrus ([MTG], 62)" |
| "Cerebellum ([CB], 13)" - "Middle temporal gyrus ([MTG], 77)" |
| "Cerebellum ([CB], 13)" - "Inferior parietal lobule ([IPL], 68)" |
| "Cerebellum ([CB], 13)" - "Superior medial frontal gyrus ([SMFG], 43)" |
| "Cerebellum ([CB], 7)" - "Superior medial frontal gyrus ([SMFG], 43)" |
| "Cerebellum ([CB], 13)" - "Left inferior parietal lobue ([R IPL], 79)" |
| "Cerebellum ([CB], 7)" - "Left inferior parietal lobue ([R IPL], 79)" |
| "Cerebellum ([CB], 13)" - "Superior frontal gyrus ([SFG], 96)" |
| "Cerebellum ([CB], 7)" - "Superior frontal gyrus ([SFG], 96)" |
| "Cerebellum ([CB], 13)" - "Middle frontal gyrus ([MiFG], 88)" |
| "Cerebellum ([CB], 13)" - "Hippocampus ([HiPP], 48)" |
| "Cerebellum ([CB], 13)" - "Left inferior parietal lobue ([L IPL], 81)" |
| "Cerebellum ([CB], 7)" - "Left inferior parietal lobue ([L IPL], 81)" |
| "Cerebellum ([CB], 13)" - "Middle cingulate cortex ([MCC], 37)" |
| "Cerebellum ([CB], 13)" - "Hippocampus ([HiPP], 83)" |
| "Cerebellum ([CB], 18)" - "Hippocampus ([HiPP], 83)" |
| "Cerebellum ([CB], 7)" - "Hippocampus ([HiPP], 83)" |
| "Cerebellum ([CB], 13)" - Precuneus (40) |
| "Cerebellum ([CB], 7)" - Precuneus (40) |
| "Cerebellum ([CB], 13)" - "Anterior cingulate cortex ([ACC], 23)" |
| "Cerebellum ([CB], 7)" - "Anterior cingulate cortex ([ACC], 23)" |
| "Cerebellum ([CB], 13)" - "Posterior cingulate cortex ([PCC], 71)" |

| |
|---|
| "Cerebellum ([CB], 13)" - "Anterior cingulate cortex ([ACC], 17)" |
| "Cerebellum ([CB], 13)" - "Posterior cingulate cortex ([PCC], 94)" |
| "Cerebellum ([CB], 7)" - "Posterior cingulate cortex ([PCC], 94)" |
| "Cerebellum ([CB], 18)" - "Cerebellum ([CB], 13)" |
| "Cerebellum ([CB], 4)" - "Cerebellum ([CB], 13)" |
| "Cerebellum ([CB], 7)" - "Cerebellum ([CB], 13)" |
| "Cerebellum ([CB], 4)" - "Cerebellum ([CB], 18)" |
| "Cerebellum ([CB], 7)" - "Cerebellum ([CB], 4)" |
| state1_jica16 |
| Fusiform gyrus (93) - Caudate (69) |
| Fusiform gyrus (93) - "Superior temporal gyrus ([STG], 21)" |
| Fusiform gyrus (93) - "Middle temporal gyrus ([MTG], 56)" |
| Fusiform gyrus (93) - "Postcentral gyrus ([PoCG], 3)" |
| Fusiform gyrus (93) - "Left postcentral gyrus ([L PoCG], 9)" |
| Fusiform gyrus (93) - "Right postcentral gyrus ([R PoCG], 11)" |
| Fusiform gyrus (93) - "Superior parietal lobule ([SPL], 27)" |
| Fusiform gyrus (93) - "Paracentral lobule ([ParaCL], 54)" |
| Fusiform gyrus (93) - "Precentral gyrus ([PreCG], 66)" |
| "Right middle occipital gyrus ([R MOG], 12)" - "Superior parietal lobule ([SPL], 80)" |
| Fusiform gyrus (93) - "Superior parietal lobule ([SPL], 80)" |
| "Inferior occipital gyrus ([IOG], 20)" - "Superior parietal lobule ([SPL], 80)" |
| Fusiform gyrus (93) - "Postcentral gyrus ([PoCG], 72)" |
| "Superior medial frontal gyrus ([SMFG], 43)" - "Right middle occipital gyrus ([R MOG], 12)" |
| "Left inferior parietal lobue ([R IPL], 79)" - "Right middle occipital gyrus ([R MOG], 12)" |
| "Superior frontal gyrus ([SFG], 96)" - "Right middle occipital gyrus ([R MOG], 12)" |
| "Anterior cingulate cortex ([ACC], 23)" - "Right middle occipital gyrus ([R MOG], 12)" |
| "Posterior cingulate cortex ([PCC], 94)" - "Right middle occipital gyrus ([R MOG], 12)" |
| "Inferior occipital gyrus ([IOG], 20)" - Fusiform gyrus (93) |
| "Middle temporal gyrus ([MTG], 77)" - Fusiform gyrus (93) |
| "Inferior parietal lobule ([IPL], 68)" - Fusiform gyrus (93) |
| "Superior medial frontal gyrus ([SMFG], 43)" - Fusiform gyrus (93) |
| "Left inferior parietal lobue ([R IPL], 79)" - Fusiform gyrus (93) |
| "Superior frontal gyrus ([SFG], 96)" - Fusiform gyrus (93) |
| "Left inferior parietal lobue ([L IPL], 81)" - Fusiform gyrus (93) |
| "Middle cingulate cortex ([MCC], 37)" - Fusiform gyrus (93) |
| "Middle frontal gyrus ([MiFG], 38)" - Fusiform gyrus (93) |
| "Hippocampus ([HiPP], 83)" - Fusiform gyrus (93) |
| Precuneus (40) - Fusiform gyrus (93) |
| "Anterior cingulate cortex ([ACC], 23)" - Fusiform gyrus (93) |
| "Posterior cingulate cortex ([PCC], 71)" - Fusiform gyrus (93) |
| "Anterior cingulate cortex ([ACC], 17)" - Fusiform gyrus (93) |
| "Posterior cingulate cortex ([PCC], 94)" - Fusiform gyrus (93) |
| "Cerebellum ([CB], 18)" - Fusiform gyrus (93) |
| "Cerebellum ([CB], 4)" - Fusiform gyrus (93) |

| |
|---|
| "Cerebellum ([CB], 7)" - Fusiform gyrus (93) |
| "Superior medial frontal gyrus ([SMFG], 43)" - "Inferior occipital gyrus ([IOG], 20)" |
| "Left inferior parietal lobue ([R IPL], 79)" - "Inferior occipital gyrus ([IOG], 20)" |
| "Posterior cingulate cortex ([PCC], 94)" - "Inferior occipital gyrus ([IOG], 20)" |
| state1_jica19 |
| "Middle temporal gyrus ([MTG], 62)" - Putamen (98) |
| "Middle temporal gyrus ([MTG], 62)" - "Superior temporal gyrus ([STG], 21)" |
| "Middle temporal gyrus ([MTG], 62)" - "Middle temporal gyrus ([MTG], 56)" |
| "Middle temporal gyrus ([MTG], 62)" - "Postcentral gyrus ([PoCG], 3)" |
| "Middle temporal gyrus ([MTG], 62)" - "Left postcentral gyrus ([L PoCG], 9)" |
| "Middle temporal gyrus ([MTG], 62)" - "Paracentral lobule ([ParaCL], 2)" |
| "Middle temporal gyrus ([MTG], 62)" - "Right postcentral gyrus ([R PoCG], 11)" |
| "Middle temporal gyrus ([MTG], 62)" - "Superior parietal lobule ([SPL], 27)" |
| "Middle temporal gyrus ([MTG], 62)" - "Paracentral lobule ([ParaCL], 54)" |
| "Middle temporal gyrus ([MTG], 62)" - "Precentral gyrus ([PreCG], 66)" |
| "Middle temporal gyrus ([MTG], 62)" - "Superior parietal lobule ([SPL], 80)" |
| "Middle temporal gyrus ([MTG], 62)" - "Postcentral gyrus ([PoCG], 72)" |
| "Inferior parietal lobule ([IPL], 68)" - "Middle temporal gyrus ([MTG], 62)" |
| "Superior medial frontal gyrus ([SMFG], 43)" - "Middle temporal gyrus ([MTG], 62)" |
| "Left inferior parietal lobue ([R IPL], 79)" - "Middle temporal gyrus ([MTG], 62)" |
| "Superior frontal gyrus ([SFG], 96)" - "Middle temporal gyrus ([MTG], 62)" |
| "Middle frontal gyrus ([MiFG], 88)" - "Middle temporal gyrus ([MTG], 62)" |
| "Left inferior parietal lobue ([L IPL], 81)" - "Middle temporal gyrus ([MTG], 62)" |
| "Middle cingulate cortex ([MCC], 37)" - "Middle temporal gyrus ([MTG], 62)" |
| "Middle frontal gyrus ([MiFG], 38)" - "Middle temporal gyrus ([MTG], 62)" |
| "Hippocampus ([HiPP], 83)" - "Middle temporal gyrus ([MTG], 62)" |
| Precuneus (32) - "Middle temporal gyrus ([MTG], 62)" |
| Precuneus (40) - "Middle temporal gyrus ([MTG], 62)" |
| "Anterior cingulate cortex ([ACC], 23)" - "Middle temporal gyrus ([MTG], 62)" |
| "Posterior cingulate cortex ([PCC], 71)" - "Middle temporal gyrus ([MTG], 62)" |
| "Anterior cingulate cortex ([ACC], 17)" - "Middle temporal gyrus ([MTG], 62)" |
| "Posterior cingulate cortex ([PCC], 94)" - "Middle temporal gyrus ([MTG], 62)" |
| "Cerebellum ([CB], 18)" - "Middle temporal gyrus ([MTG], 62)" |
| "Cerebellum ([CB], 4)" - "Middle temporal gyrus ([MTG], 62)" |
| "Cerebellum ([CB], 7)" - "Middle temporal gyrus ([MTG], 62)" |
| state1_jica29 |
| "Anterior cingulate cortex ([ACC], 23)" - "Superior temporal gyrus ([STG], 21)" |
| "Middle cingulate cortex ([MCC], 37)" - "Middle temporal gyrus ([MTG], 56)" |
| "Middle cingulate cortex ([MCC], 37)" - "Left postcentral gyrus ([L PoCG], 9)" |
| "Middle cingulate cortex ([MCC], 37)" - "Right postcentral gyrus ([R PoCG], 11)" |
| "Middle cingulate cortex ([MCC], 37)" - "Paracentral lobule ([ParaCL], 54)" |
| "Middle cingulate cortex ([MCC], 37)" - "Precentral gyrus ([PreCG], 66)" |
| "Middle cingulate cortex ([MCC], 37)" - Insula (33) |
| "Middle cingulate cortex ([MCC], 37)" - "Left inferior parietal lobue ([R IPL], 79)" |

| |
|---|
| "Middle cingulate cortex ([MCC], 37)" - "Superior frontal gyrus ([SFG], 96)" |
| Precuneus (40) - "Middle cingulate cortex ([MCC], 37)" |
| "Posterior cingulate cortex ([PCC], 94)" - "Middle cingulate cortex ([MCC], 37)" |
| state1_jica30 |
| "Right inferior frontal gyrus ([R IFG], 61)" - Caudate (69) |
| "Right inferior frontal gyrus ([R IFG], 61)" - Caudate (99) |
| "Inferior parietal lobule ([IPL], 63)" - "Lingual gyrus ([LingualG], 8)" |
| "Middle frontal gyrus ([MiFG], 38)" - "Inferior parietal lobule ([IPL], 63)" |
| Precuneus (51) - "Middle cingulate cortex ([MCC], 37)" |
| Precuneus (51) - "Middle frontal gyrus ([MiFG], 38)" |
| "Cerebellum ([CB], 18)" - "Cerebellum ([CB], 13)" |
| state1_jica33 |
| "Middle temporal gyrus ([MTG], 56)" - Thalamus (45) |
| Insula (33) - Thalamus (45) |
| "Middle frontal gyrus ([MiFG], 88)" - Thalamus (45) |
| "Middle cingulate cortex ([MCC], 37)" - "Hippocampus ([HiPP], 48)" |
| "Hippocampus ([HiPP], 83)" - "Middle cingulate cortex ([MCC], 37)" |
| "Anterior cingulate cortex ([ACC], 17)" - "Middle cingulate cortex ([MCC], 37)" |
| state2_jica2 |
| "Middle temporal gyrus ([MTG], 62)" - Putamen (98) |
| "Superior parietal lobule ([SPL], 80)" - "Superior temporal gyrus ([STG], 21)" |
| "Middle temporal gyrus ([MTG], 62)" - "Superior temporal gyrus ([STG], 21)" |
| "Superior parietal lobule ([SPL], 80)" - "Middle temporal gyrus ([MTG], 56)" |
| "Middle temporal gyrus ([MTG], 62)" - "Middle temporal gyrus ([MTG], 56)" |
| "Right inferior frontal gyrus ([R IFG], 61)" - "Middle temporal gyrus ([MTG], 56)" |
| "Superior parietal lobule ([SPL], 80)" - "Postcentral gyrus ([PoCG], 3)" |
| "Middle temporal gyrus ([MTG], 62)" - "Postcentral gyrus ([PoCG], 3)" |
| "Superior parietal lobule ([SPL], 80)" - "Left postcentral gyrus ([L PoCG], 9)" |
| "Middle temporal gyrus ([MTG], 62)" - "Left postcentral gyrus ([L PoCG], 9)" |
| "Superior parietal lobule ([SPL], 80)" - "Paracentral lobule ([ParaCL], 2)" |
| "Middle temporal gyrus ([MTG], 62)" - "Paracentral lobule ([ParaCL], 2)" |
| "Superior parietal lobule ([SPL], 80)" - "Right postcentral gyrus ([R PoCG], 11)" |
| "Middle temporal gyrus ([MTG], 62)" - "Right postcentral gyrus ([R PoCG], 11)" |
| "Left inferior parietal lobue ([R IPL], 79)" - "Right postcentral gyrus ([R PoCG], 11)" |
| "Middle temporal gyrus ([MTG], 62)" - "Superior parietal lobule ([SPL], 27)" |
| "Right inferior frontal gyrus ([R IFG], 61)" - "Superior parietal lobule ([SPL], 27)" |
| "Middle temporal gyrus ([MTG], 62)" - "Paracentral lobule ([ParaCL], 54)" |
| "Middle temporal gyrus ([MTG], 62)" - "Precentral gyrus ([PreCG], 66)" |
| "Middle frontal gyrus ([MiFG], 55)" - "Superior parietal lobule ([SPL], 80)" |
| "Cerebellum ([CB], 18)" - "Superior parietal lobule ([SPL], 80)" |
| "Inferior parietal lobule ([IPL], 68)" - "Middle temporal gyrus ([MTG], 62)" |
| "Superior medial frontal gyrus ([SMFG], 43)" - "Middle temporal gyrus ([MTG], 62)" |
| "Superior frontal gyrus ([SFG], 96)" - "Middle temporal gyrus ([MTG], 62)" |
| Precuneus (32) - "Middle temporal gyrus ([MTG], 62)" |

| |
|---|
| Precuneus (40) - "Middle temporal gyrus ([MTG], 62)" |
| "Posterior cingulate cortex ([PCC], 94)" - "Middle temporal gyrus ([MTG], 62)" |
| "Cerebellum ([CB], 18)" - "Middle temporal gyrus ([MTG], 62)" |
| "Right inferior frontal gyrus ([R IFG], 61)" - "Inferior parietal lobule ([IPL], 68)" |
| "Superior frontal gyrus ([SFG], 96)" - "Right inferior frontal gyrus ([R IFG], 61)" |
| "Cerebellum ([CB], 18)" - "Right inferior frontal gyrus ([R IFG], 61)" |
| state2_jica4 |
| "Inferior frontal gyrus ([IFG], 70)" - "Superior parietal lobule ([SPL], 80)" |
| Precuneus (32) - "Superior parietal lobule ([SPL], 80)" |
| Precuneus (40) - "Superior parietal lobule ([SPL], 80)" |
| "Anterior cingulate cortex ([ACC], 23)" - "Superior parietal lobule ([SPL], 80)" |
| Precuneus (51) - "Superior parietal lobule ([SPL], 80)" |
| "Cerebellum ([CB], 7)" - "Superior parietal lobule ([SPL], 80)" |
| "Inferior frontal gyrus ([IFG], 70)" - "Superior medial frontal gyrus ([SMFG], 43)" |
| "Middle frontal gyrus ([MiFG], 55)" - "Superior medial frontal gyrus ([SMFG], 43)" |
| Precuneus (32) - "Superior medial frontal gyrus ([SMFG], 43)" |
| Precuneus (40) - "Superior medial frontal gyrus ([SMFG], 43)" |
| "Anterior cingulate cortex ([ACC], 23)" - "Superior medial frontal gyrus ([SMFG], 43)" |
| "Posterior cingulate cortex ([PCC], 71)" - "Superior medial frontal gyrus ([SMFG], 43)" |
| Precuneus (51) - "Superior medial frontal gyrus ([SMFG], 43)" |
| "Cerebellum ([CB], 7)" - "Superior medial frontal gyrus ([SMFG], 43)" |
| "Left inferior parietal lobue ([R IPL], 79)" - "Inferior frontal gyrus ([IFG], 70)" |
| "Posterior cingulate cortex ([PCC], 94)" - "Inferior frontal gyrus ([IFG], 70)" |
| "Left inferior parietal lobue ([R IPL], 79)" - "Middle frontal gyrus ([MiFG], 55)" |
| "Superior frontal gyrus ([SFG], 96)" - "Middle frontal gyrus ([MiFG], 55)" |
| "Left inferior parietal lobue ([L IPL], 81)" - "Middle frontal gyrus ([MiFG], 55)" |
| "Anterior cingulate cortex ([ACC], 23)" - "Middle frontal gyrus ([MiFG], 55)" |
| "Posterior cingulate cortex ([PCC], 94)" - "Middle frontal gyrus ([MiFG], 55)" |
| Precuneus (40) - "Left inferior parietal lobue ([R IPL], 79)" |
| "Anterior cingulate cortex ([ACC], 23)" - "Left inferior parietal lobue ([R IPL], 79)" |
| "Posterior cingulate cortex ([PCC], 71)" - "Left inferior parietal lobue ([R IPL], 79)" |
| Precuneus (51) - "Left inferior parietal lobue ([R IPL], 79)" |
| "Cerebellum ([CB], 7)" - "Left inferior parietal lobue ([R IPL], 79)" |
| "Anterior cingulate cortex ([ACC], 23)" - "Left inferior parietal lobue ([L IPL], 81)" |
| Precuneus (51) - "Left inferior parietal lobue ([L IPL], 81)" |
| "Anterior cingulate cortex ([ACC], 23)" - "Inferior frontal gyrus ([IFG], 67)" |
| "Posterior cingulate cortex ([PCC], 94)" - "Anterior cingulate cortex ([ACC], 23)" |
| "Posterior cingulate cortex ([PCC], 94)" - "Posterior cingulate cortex ([PCC], 71)" |
| "Posterior cingulate cortex ([PCC], 94)" - Precuneus (51) |
| "Cerebellum ([CB], 7)" - "Posterior cingulate cortex ([PCC], 94)" |
| state2_jica5 |
| "Cerebellum ([CB], 13)" - Putamen (98) |
| Insula (33) - "Superior temporal gyrus ([STG], 21)" |
| "Cerebellum ([CB], 13)" - "Superior temporal gyrus ([STG], 21)" |

| |
|---|
| "Cerebellum ([CB], 18)" - "Superior temporal gyrus ([STG], 21)" |
| "Cerebellum ([CB], 7)" - "Superior temporal gyrus ([STG], 21)" |
| "Cerebellum ([CB], 13)" - "Middle temporal gyrus ([MTG], 56)" |
| "Cerebellum ([CB], 7)" - "Middle temporal gyrus ([MTG], 56)" |
| Insula (33) - "Postcentral gyrus ([PoCG], 3)" |
| "Cerebellum ([CB], 13)" - "Postcentral gyrus ([PoCG], 3)" |
| "Cerebellum ([CB], 7)" - "Postcentral gyrus ([PoCG], 3)" |
| Insula (33) - "Left postcentral gyrus ([L PoCG], 9)" |
| "Cerebellum ([CB], 7)" - "Left postcentral gyrus ([L PoCG], 9)" |
| Insula (33) - "Right postcentral gyrus ([R PoCG], 11)" |
| "Cerebellum ([CB], 13)" - "Superior parietal lobule ([SPL], 27)" |
| "Cerebellum ([CB], 13)" - "Precentral gyrus ([PreCG], 66)" |
| "Cerebellum ([CB], 18)" - "Precentral gyrus ([PreCG], 66)" |
| "Cerebellum ([CB], 7)" - "Precentral gyrus ([PreCG], 66)" |
| "Cerebellum ([CB], 13)" - Insula (33) |
| "Cerebellum ([CB], 18)" - Insula (33) |
| "Cerebellum ([CB], 7)" - Insula (33) |
| "Cerebellum ([CB], 7)" - "Supplementary motor area ([SMA], 84)" |
| "Cerebellum ([CB], 13)" - "Superior frontal gyrus ([SFG], 96)" |
| "Cerebellum ([CB], 18)" - "Cerebellum ([CB], 13)" |
| "Cerebellum ([CB], 7)" - "Cerebellum ([CB], 13)" |
| "Cerebellum ([CB], 4)" - "Cerebellum ([CB], 18)" |
| state2_jica8 |
| "Inferior frontal gyrus ([IFG], 70)" - "Precentral gyrus ([PreCG], 66)" |
| Precuneus (51) - "Precentral gyrus ([PreCG], 66)" |
| Precuneus (32) - Insula (33) |
| "Posterior cingulate cortex ([PCC], 71)" - Insula (33) |
| "Cerebellum ([CB], 7)" - Insula (33) |
| "Superior frontal gyrus ([SFG], 96)" - "Inferior frontal gyrus ([IFG], 70)" |
| Precuneus (40) - "Inferior frontal gyrus ([IFG], 70)" |
| "Anterior cingulate cortex ([ACC], 23)" - "Inferior frontal gyrus ([IFG], 70)" |
| "Anterior cingulate cortex ([ACC], 17)" - "Inferior frontal gyrus ([IFG], 70)" |
| "Posterior cingulate cortex ([PCC], 94)" - "Inferior frontal gyrus ([IFG], 70)" |
| Precuneus (32) - "Right inferior frontal gyrus ([R IFG], 61)" |
| "Posterior cingulate cortex ([PCC], 71)" - "Right inferior frontal gyrus ([R IFG], 61)" |
| "Posterior cingulate cortex ([PCC], 71)" - "Inferior parietal lobule ([IPL], 63)" |
| "Left inferior parietal lobue ([L IPL], 81)" - "Supplementary motor area ([SMA], 84)" |
| Precuneus (40) - "Supplementary motor area ([SMA], 84)" |
| "Anterior cingulate cortex ([ACC], 23)" - "Supplementary motor area ([SMA], 84)" |
| "Anterior cingulate cortex ([ACC], 17)" - "Supplementary motor area ([SMA], 84)" |
| "Posterior cingulate cortex ([PCC], 94)" - "Supplementary motor area ([SMA], 84)" |
| Precuneus (51) - "Superior frontal gyrus ([SFG], 96)" |
| Precuneus (32) - "Hippocampus ([HiPP], 48)" |
| "Posterior cingulate cortex ([PCC], 71)" - "Hippocampus ([HiPP], 48)" |

| |
|---|
| Precuneus (32) - "Hippocampus ([HiPP], 83)" |
| "Posterior cingulate cortex ([PCC], 71)" - "Hippocampus ([HiPP], 83)" |
| "Anterior cingulate cortex ([ACC], 17)" - Precuneus (32) |
| "Anterior cingulate cortex ([ACC], 17)" - "Posterior cingulate cortex ([PCC], 71)" |
| Precuneus (51) - "Anterior cingulate cortex ([ACC], 17)" |
| "Cerebellum ([CB], 7)" - "Anterior cingulate cortex ([ACC], 17)" |
| "Posterior cingulate cortex ([PCC], 94)" - Precuneus (51) |
| state2_jica9 |
| "Superior medial frontal gyrus ([SMFG], 43)" - "Postcentral gyrus ([PoCG], 3)" |
| "Anterior cingulate cortex ([ACC], 23)" - "Postcentral gyrus ([PoCG], 3)" |
| "Posterior cingulate cortex ([PCC], 94)" - "Postcentral gyrus ([PoCG], 3)" |
| "Superior medial frontal gyrus ([SMFG], 43)" - "Left postcentral gyrus ([L PoCG], 9)" |
| "Superior frontal gyrus ([SFG], 96)" - "Left postcentral gyrus ([L PoCG], 9)" |
| Precuneus (40) - "Left postcentral gyrus ([L PoCG], 9)" |
| "Anterior cingulate cortex ([ACC], 23)" - "Left postcentral gyrus ([L PoCG], 9)" |
| "Anterior cingulate cortex ([ACC], 17)" - "Left postcentral gyrus ([L PoCG], 9)" |
| "Posterior cingulate cortex ([PCC], 94)" - "Left postcentral gyrus ([L PoCG], 9)" |
| "Superior medial frontal gyrus ([SMFG], 43)" - "Paracentral lobule ([ParaCL], 2)" |
| "Superior frontal gyrus ([SFG], 96)" - "Paracentral lobule ([ParaCL], 2)" |
| Precuneus (40) - "Paracentral lobule ([ParaCL], 2)" |
| "Anterior cingulate cortex ([ACC], 23)" - "Paracentral lobule ([ParaCL], 2)" |
| "Posterior cingulate cortex ([PCC], 94)" - "Paracentral lobule ([ParaCL], 2)" |
| "Superior medial frontal gyrus ([SMFG], 43)" - "Right postcentral gyrus ([R PoCG], 11)" |
| "Left inferior parietal lobue ([R IPL], 79)" - "Right postcentral gyrus ([R PoCG], 11)" |
| "Superior frontal gyrus ([SFG], 96)" - "Right postcentral gyrus ([R PoCG], 11)" |
| Precuneus (40) - "Right postcentral gyrus ([R PoCG], 11)" |
| "Anterior cingulate cortex ([ACC], 23)" - "Right postcentral gyrus ([R PoCG], 11)" |
| "Anterior cingulate cortex ([ACC], 17)" - "Right postcentral gyrus ([R PoCG], 11)" |
| "Posterior cingulate cortex ([PCC], 94)" - "Right postcentral gyrus ([R PoCG], 11)" |
| "Superior medial frontal gyrus ([SMFG], 43)" - "Superior parietal lobule ([SPL], 27)" |
| "Anterior cingulate cortex ([ACC], 23)" - "Superior parietal lobule ([SPL], 27)" |
| "Posterior cingulate cortex ([PCC], 94)" - "Superior parietal lobule ([SPL], 27)" |
| "Inferior frontal gyrus ([IFG], 67)" - "Superior medial frontal gyrus ([SMFG], 43)" |
| "Anterior cingulate cortex ([ACC], 23)" - "Inferior parietal lobule ([IPL], 63)" |
| "Inferior frontal gyrus ([IFG], 67)" - "Superior frontal gyrus ([SFG], 96)" |
| "Anterior cingulate cortex ([ACC], 23)" - "Inferior frontal gyrus ([IFG], 67)" |
| "Posterior cingulate cortex ([PCC], 94)" - "Inferior frontal gyrus ([IFG], 67)" |
| state2_jica12 |
| "Superior temporal gyrus ([STG], 21)" - Caudate (69) |
| Insula (33) - Caudate (69) |
| "Cerebellum ([CB], 13)" - Caudate (69) |
| "Cerebellum ([CB], 7)" - Caudate (69) |
| "Superior temporal gyrus ([STG], 21)" - Subthalamus/hypothalamus (53) |
| "Middle temporal gyrus ([MTG], 56)" - Subthalamus/hypothalamus (53) |

| |
|---|
| Insula (33) - Subthalamus/hypothalamus (53) |
| "Cerebellum ([CB], 13)" - Subthalamus/hypothalamus (53) |
| "Cerebellum ([CB], 18)" - Subthalamus/hypothalamus (53) |
| "Cerebellum ([CB], 7)" - Subthalamus/hypothalamus (53) |
| "Cerebellum ([CB], 7)" - Putamen (98) |
| "Inferior parietal lobule ([IPL], 63)" - "Superior temporal gyrus ([STG], 21)" |
| "Hippocampus ([HiPP], 48)" - "Superior temporal gyrus ([STG], 21)" |
| "Hippocampus ([HiPP], 83)" - "Superior temporal gyrus ([STG], 21)" |
| "Hippocampus ([HiPP], 48)" - "Middle temporal gyrus ([MTG], 56)" |
| "Hippocampus ([HiPP], 83)" - "Middle temporal gyrus ([MTG], 56)" |
| "Hippocampus ([HiPP], 48)" - Insula (33) |
| "Hippocampus ([HiPP], 83)" - Insula (33) |
| "Cerebellum ([CB], 7)" - "Inferior parietal lobule ([IPL], 63)" |
| "Cerebellum ([CB], 13)" - "Hippocampus ([HiPP], 48)" |
| "Cerebellum ([CB], 18)" - "Hippocampus ([HiPP], 48)" |
| "Cerebellum ([CB], 7)" - "Hippocampus ([HiPP], 48)" |
| "Cerebellum ([CB], 7)" - "Middle frontal gyrus ([MiFG], 38)" |
| "Cerebellum ([CB], 13)" - "Hippocampus ([HiPP], 83)" |
| "Cerebellum ([CB], 18)" - "Hippocampus ([HiPP], 83)" |
| "Cerebellum ([CB], 7)" - "Hippocampus ([HiPP], 83)" |
| "Cerebellum ([CB], 7)" - Precuneus (32) |
| "Cerebellum ([CB], 7)" - Precuneus (51) |
| "Cerebellum ([CB], 4)" - "Cerebellum ([CB], 13)" |
| "Cerebellum ([CB], 7)" - "Cerebellum ([CB], 4)" |
| state2_jica22 |
| "Postcentral gyrus ([PoCG], 3)" - Subthalamus/hypothalamus (53) |
| "Left postcentral gyrus ([L PoCG], 9)" - Subthalamus/hypothalamus (53) |
| "Paracentral lobule ([ParaCL], 2)" - Subthalamus/hypothalamus (53) |
| "Right postcentral gyrus ([R PoCG], 11)" - Subthalamus/hypothalamus (53) |
| "Superior parietal lobule ([SPL], 27)" - Subthalamus/hypothalamus (53) |
| "Postcentral gyrus ([PoCG], 3)" - Putamen (98) |
| "Left postcentral gyrus ([L PoCG], 9)" - Putamen (98) |
| "Paracentral lobule ([ParaCL], 2)" - Putamen (98) |
| "Right postcentral gyrus ([R PoCG], 11)" - Putamen (98) |
| "Superior parietal lobule ([SPL], 27)" - Putamen (98) |
| "Cerebellum ([CB], 13)" - Putamen (98) |
| "Cerebellum ([CB], 18)" - Putamen (98) |
| "Postcentral gyrus ([PoCG], 3)" - Caudate (99) |
| "Left postcentral gyrus ([L PoCG], 9)" - Caudate (99) |
| "Paracentral lobule ([ParaCL], 2)" - Caudate (99) |
| "Right postcentral gyrus ([R PoCG], 11)" - Caudate (99) |
| "Superior parietal lobule ([SPL], 27)" - Caudate (99) |
| "Cerebellum ([CB], 13)" - Caudate (99) |
| "Postcentral gyrus ([PoCG], 3)" - Thalamus (45) |

| |
|---|
| "Left postcentral gyrus ([L PoCG], 9)" - Thalamus (45) |
| "Paracentral lobule ([ParaCL], 2)" - Thalamus (45) |
| "Right postcentral gyrus ([R PoCG], 11)" - Thalamus (45) |
| "Superior parietal lobule ([SPL], 27)" - Thalamus (45) |
| "Cerebellum ([CB], 13)" - Thalamus (45) |
| "Cerebellum ([CB], 18)" - Thalamus (45) |
| "Paracentral lobule ([ParaCL], 2)" - "Left postcentral gyrus ([L PoCG], 9)" |
| "Right postcentral gyrus ([R PoCG], 11)" - "Paracentral lobule ([ParaCL], 2)" |
| state2_jica25 |
| "Calcarine gyrus ([CalcarineG], 16)" - Caudate (69) |
| "Middle occipital gyrus ([MOG], 5)" - Caudate (69) |
| Cuneus (15) - Caudate (69) |
| "Right middle occipital gyrus ([R MOG], 12)" - Caudate (69) |
| "Inferior occipital gyrus ([IOG], 20)" - Caudate (69) |
| "Lingual gyrus ([LingualG], 8)" - Caudate (69) |
| "Calcarine gyrus ([CalcarineG], 16)" - Subthalamus/hypothalamus (53) |
| "Middle occipital gyrus ([MOG], 5)" - Subthalamus/hypothalamus (53) |
| Cuneus (15) - Subthalamus/hypothalamus (53) |
| "Right middle occipital gyrus ([R MOG], 12)" - Subthalamus/hypothalamus (53) |
| "Inferior occipital gyrus ([IOG], 20)" - Subthalamus/hypothalamus (53) |
| "Lingual gyrus ([LingualG], 8)" - Subthalamus/hypothalamus (53) |
| "Middle occipital gyrus ([MOG], 5)" - Putamen (98) |
| Cuneus (15) - Putamen (98) |
| "Right middle occipital gyrus ([R MOG], 12)" - Putamen (98) |
| "Inferior occipital gyrus ([IOG], 20)" - Putamen (98) |
| "Lingual gyrus ([LingualG], 8)" - Putamen (98) |
| "Middle occipital gyrus ([MOG], 5)" - Caudate (99) |
| Cuneus (15) - Caudate (99) |
| "Lingual gyrus ([LingualG], 8)" - Caudate (99) |
| "Middle occipital gyrus ([MOG], 5)" - Thalamus (45) |
| Cuneus (15) - Thalamus (45) |
| "Lingual gyrus ([LingualG], 8)" - Thalamus (45) |
| "Hippocampus ([HiPP], 83)" - "Calcarine gyrus ([CalcarineG], 16)" |
| "Hippocampus ([HiPP], 48)" - "Middle occipital gyrus ([MOG], 5)" |
| "Hippocampus ([HiPP], 83)" - "Middle occipital gyrus ([MOG], 5)" |
| "Cerebellum ([CB], 4)" - "Middle occipital gyrus ([MOG], 5)" |
| Cuneus (15) - "Middle temporal gyrus ([MTG], 62)" |
| "Lingual gyrus ([LingualG], 8)" - "Middle temporal gyrus ([MTG], 62)" |
| "Hippocampus ([HiPP], 48)" - Cuneus (15) |
| "Hippocampus ([HiPP], 83)" - Cuneus (15) |
| "Cerebellum ([CB], 4)" - Cuneus (15) |
| "Hippocampus ([HiPP], 48)" - "Right middle occipital gyrus ([R MOG], 12)" |
| "Hippocampus ([HiPP], 83)" - "Right middle occipital gyrus ([R MOG], 12)" |
| "Hippocampus ([HiPP], 48)" - "Inferior occipital gyrus ([IOG], 20)" |

| |
|---|
| "Hippocampus ([HiPP], 83)" - "Inferior occipital gyrus ([IOG], 20)" |
| "Hippocampus ([HiPP], 48)" - "Lingual gyrus ([LingualG], 8)" |
| "Hippocampus ([HiPP], 83)" - "Lingual gyrus ([LingualG], 8)" |
| "Cerebellum ([CB], 4)" - "Lingual gyrus ([LingualG], 8)" |
| state2_jica27 |
| "Calcarine gyrus ([CalcarineG], 16)" - "Postcentral gyrus ([PoCG], 3)" |
| "Middle occipital gyrus ([MOG], 5)" - "Postcentral gyrus ([PoCG], 3)" |
| Cuneus (15) - "Postcentral gyrus ([PoCG], 3)" |
| "Lingual gyrus ([LingualG], 8)" - "Postcentral gyrus ([PoCG], 3)" |
| "Calcarine gyrus ([CalcarineG], 16)" - "Left postcentral gyrus ([L PoCG], 9)" |
| "Middle occipital gyrus ([MOG], 5)" - "Left postcentral gyrus ([L PoCG], 9)" |
| Cuneus (15) - "Left postcentral gyrus ([L PoCG], 9)" |
| "Right middle occipital gyrus ([R MOG], 12)" - "Left postcentral gyrus ([L PoCG], 9)" |
| "Inferior occipital gyrus ([IOG], 20)" - "Left postcentral gyrus ([L PoCG], 9)" |
| "Lingual gyrus ([LingualG], 8)" - "Left postcentral gyrus ([L PoCG], 9)" |
| "Calcarine gyrus ([CalcarineG], 16)" - "Paracentral lobule ([ParaCL], 2)" |
| "Middle occipital gyrus ([MOG], 5)" - "Paracentral lobule ([ParaCL], 2)" |
| Cuneus (15) - "Paracentral lobule ([ParaCL], 2)" |
| "Right middle occipital gyrus ([R MOG], 12)" - "Paracentral lobule ([ParaCL], 2)" |
| "Inferior occipital gyrus ([IOG], 20)" - "Paracentral lobule ([ParaCL], 2)" |
| "Lingual gyrus ([LingualG], 8)" - "Paracentral lobule ([ParaCL], 2)" |
| "Calcarine gyrus ([CalcarineG], 16)" - "Right postcentral gyrus ([R PoCG], 11)" |
| "Middle occipital gyrus ([MOG], 5)" - "Right postcentral gyrus ([R PoCG], 11)" |
| Cuneus (15) - "Right postcentral gyrus ([R PoCG], 11)" |
| "Right middle occipital gyrus ([R MOG], 12)" - "Right postcentral gyrus ([R PoCG], 11)" |
| "Inferior occipital gyrus ([IOG], 20)" - "Right postcentral gyrus ([R PoCG], 11)" |
| "Lingual gyrus ([LingualG], 8)" - "Right postcentral gyrus ([R PoCG], 11)" |
| "Calcarine gyrus ([CalcarineG], 16)" - "Superior parietal lobule ([SPL], 27)" |
| "Middle occipital gyrus ([MOG], 5)" - "Superior parietal lobule ([SPL], 27)" |
| Cuneus (15) - "Superior parietal lobule ([SPL], 27)" |
| "Right middle occipital gyrus ([R MOG], 12)" - "Superior parietal lobule ([SPL], 27)" |
| "Inferior occipital gyrus ([IOG], 20)" - "Superior parietal lobule ([SPL], 27)" |
| "Lingual gyrus ([LingualG], 8)" - "Superior parietal lobule ([SPL], 27)" |
| "Middle occipital gyrus ([MOG], 5)" - "Postcentral gyrus ([PoCG], 72)" |
| Cuneus (15) - "Postcentral gyrus ([PoCG], 72)" |
| "Lingual gyrus ([LingualG], 8)" - "Postcentral gyrus ([PoCG], 72)" |
| state2_jica28 |
| "Superior medial frontal gyrus ([SMFG], 43)" - Putamen (98) |
| "Anterior cingulate cortex ([ACC], 23)" - Putamen (98) |
| "Posterior cingulate cortex ([PCC], 94)" - Putamen (98) |
| "Superior medial frontal gyrus ([SMFG], 43)" - "Middle temporal gyrus ([MTG], 56)" |
| "Superior frontal gyrus ([SFG], 96)" - "Middle temporal gyrus ([MTG], 56)" |
| "Anterior cingulate cortex ([ACC], 23)" - "Middle temporal gyrus ([MTG], 56)" |
| "Posterior cingulate cortex ([PCC], 94)" - "Middle temporal gyrus ([MTG], 56)" |

| |
|---|
| "Superior medial frontal gyrus ([SMFG], 43)" - "Precentral gyrus ([PreCG], 66)" |
| "Left inferior parietal lobue ([R IPL], 79)" - "Precentral gyrus ([PreCG], 66)" |
| "Posterior cingulate cortex ([PCC], 94)" - "Precentral gyrus ([PreCG], 66)" |
| "Superior medial frontal gyrus ([SMFG], 43)" - Insula (33) |
| "Left inferior parietal lobue ([R IPL], 79)" - Insula (33) |
| "Superior frontal gyrus ([SFG], 96)" - Insula (33) |
| "Anterior cingulate cortex ([ACC], 23)" - Insula (33) |
| "Posterior cingulate cortex ([PCC], 94)" - Insula (33) |
| "Right inferior frontal gyrus ([R IFG], 61)" - "Superior medial frontal gyrus ([SMFG], 43)" |
| "Superior frontal gyrus ([SFG], 96)" - "Superior medial frontal gyrus ([SMFG], 43)" |
| "Posterior cingulate cortex ([PCC], 94)" - "Right inferior frontal gyrus ([R IFG], 61)" |
| "Middle frontal gyrus ([MiFG], 88)" - "Superior frontal gyrus ([SFG], 96)" |
| Precuneus (40) - "Superior frontal gyrus ([SFG], 96)" |
| "Posterior cingulate cortex ([PCC], 94)" - "Superior frontal gyrus ([SFG], 96)" |
| "Posterior cingulate cortex ([PCC], 94)" - "Middle frontal gyrus ([MiFG], 88)" |
| state2_jica29 |
| "Right middle occipital gyrus ([R MOG], 12)" - "Middle temporal gyrus ([MTG], 62)" |
| "Inferior occipital gyrus ([IOG], 20)" - "Middle temporal gyrus ([MTG], 62)" |
| Insula (33) - "Middle temporal gyrus ([MTG], 62)" |
| "Middle temporal gyrus ([MTG], 77)" - Fusiform gyrus (93) |
| "Supplementary motor area ([SMA], 84)" - "Superior medial frontal gyrus ([SMFG], 43)" |
| "Supplementary motor area ([SMA], 84)" - "Left inferior parietal lobue ([R IPL], 79)" |
| "Inferior frontal gyrus ([IFG], 67)" - "Left inferior parietal lobue ([R IPL], 79)" |
| "Posterior cingulate cortex ([PCC], 94)" - "Left inferior parietal lobue ([R IPL], 79)" |
| "Superior frontal gyrus ([SFG], 96)" - "Supplementary motor area ([SMA], 84)" |
| "Posterior cingulate cortex ([PCC], 94)" - "Supplementary motor area ([SMA], 84)" |
| "Inferior frontal gyrus ([IFG], 67)" - "Superior frontal gyrus ([SFG], 96)" |
| "Posterior cingulate cortex ([PCC], 94)" - "Inferior frontal gyrus ([IFG], 67)" |
| "Cerebellum ([CB], 18)" - "Inferior frontal gyrus ([IFG], 67)" |
| "Cerebellum ([CB], 7)" - "Inferior frontal gyrus ([IFG], 67)" |
| "Cerebellum ([CB], 7)" - "Cerebellum ([CB], 13)" |
| state2_jica30 |
| "Inferior frontal gyrus ([IFG], 70)" - "Superior parietal lobule ([SPL], 80)" |
| "Superior frontal gyrus ([SFG], 96)" - "Superior parietal lobule ([SPL], 80)" |
| "Middle cingulate cortex ([MCC], 37)" - "Superior parietal lobule ([SPL], 80)" |
| Precuneus (51) - Fusiform gyrus (93) |
| "Cerebellum ([CB], 13)" - "Supplementary motor area ([SMA], 84)" |
| Precuneus (51) - "Middle cingulate cortex ([MCC], 37)" |
| state2_jica31 |
| "Left inferior parietal lobue ([R IPL], 79)" - "Right middle occipital gyrus ([R MOG], 12)" |
| "Anterior cingulate cortex ([ACC], 17)" - "Inferior parietal lobule ([IPL], 68)" |
| "Supplementary motor area ([SMA], 84)" - "Superior medial frontal gyrus ([SMFG], 43)" |
| Precuneus (32) - "Right inferior frontal gyrus ([R IFG], 61)" |
| Precuneus (40) - "Right inferior frontal gyrus ([R IFG], 61)" |

| |
|---|
| Precuneus (40) - "Left inferior parietal lobue ([R IPL], 79)" |
| Precuneus (40) - "Middle cingulate cortex ([MCC], 37)" |
| Precuneus (40) - "Inferior frontal gyrus ([IFG], 67)" |
| state2_jica32 |
| "Posterior cingulate cortex ([PCC], 94)" - "Lingual gyrus ([LingualG], 8)" |
| "Right inferior frontal gyrus ([R IFG], 61)" - "Inferior parietal lobule ([IPL], 68)" |
| "Inferior frontal gyrus ([IFG], 67)" - "Inferior frontal gyrus ([IFG], 70)" |
| Precuneus (32) - "Right inferior frontal gyrus ([R IFG], 61)" |
| state2_jica35 |
| "Right inferior frontal gyrus ([R IFG], 61)" - "Postcentral gyrus ([PoCG], 72)" |
| "Middle temporal gyrus ([MTG], 77)" - "Inferior occipital gyrus ([IOG], 20)" |
| "Inferior parietal lobule ([IPL], 63)" - "Right inferior frontal gyrus ([R IFG], 61)" |
| "Left inferior parietal lobue ([L IPL], 81)" - "Right inferior frontal gyrus ([R IFG], 61)" |
| state3_jica2 |
| Fusiform gyrus (93) - Caudate (69) |
| "Inferior parietal lobule ([IPL], 63)" - Caudate (69) |
| Precuneus (40) - Caudate (69) |
| Precuneus (40) - Subthalamus/hypothalamus (53) |
| Precuneus (40) - Caudate (99) |
| Precuneus (40) - Thalamus (45) |
| Precuneus (40) - "Postcentral gyrus ([PoCG], 3)" |
| Precuneus (40) - "Left postcentral gyrus ([L PoCG], 9)" |
| Precuneus (40) - "Paracentral lobule ([ParaCL], 2)" |
| Precuneus (40) - "Right postcentral gyrus ([R PoCG], 11)" |
| Fusiform gyrus (93) - "Superior parietal lobule ([SPL], 27)" |
| Precuneus (40) - "Superior parietal lobule ([SPL], 27)" |
| Fusiform gyrus (93) - "Precentral gyrus ([PreCG], 66)" |
| Fusiform gyrus (93) - "Superior parietal lobule ([SPL], 80)" |
| "Inferior parietal lobule ([IPL], 63)" - "Superior parietal lobule ([SPL], 80)" |
| Precuneus (40) - "Superior parietal lobule ([SPL], 80)" |
| Precuneus (40) - "Postcentral gyrus ([PoCG], 72)" |
| Fusiform gyrus (93) - "Calcarine gyrus ([CalcarineG], 16)" |
| Precuneus (40) - "Middle temporal gyrus ([MTG], 62)" |
| Precuneus (40) - Cuneus (15) |
| Precuneus (40) - "Right middle occipital gyrus ([R MOG], 12)" |
| "Inferior occipital gyrus ([IOG], 20)" - Fusiform gyrus (93) |
| "Hippocampus ([HiPP], 83)" - Fusiform gyrus (93) |
| "Posterior cingulate cortex ([PCC], 71)" - Fusiform gyrus (93) |
| Precuneus (40) - "Inferior occipital gyrus ([IOG], 20)" |
| "Inferior parietal lobule ([IPL], 63)" - "Superior medial frontal gyrus ([SMFG], 43)" |
| Precuneus (40) - "Middle cingulate cortex ([MCC], 37)" |
| Precuneus (40) - "Middle frontal gyrus ([MiFG], 38)" |
| "Anterior cingulate cortex ([ACC], 23)" - Precuneus (40) |
| "Posterior cingulate cortex ([PCC], 71)" - Precuneus (40) |

| |
|---|
| "Cerebellum ([CB], 13)" - Precuneus (40) |
| "Cerebellum ([CB], 18)" - Precuneus (40) |
| "Cerebellum ([CB], 4)" - Precuneus (40) |
| "Cerebellum ([CB], 7)" - Precuneus (40) |
| state3_jica3 |
| Precuneus (32) - Caudate (69) |
| Precuneus (32) - Subthalamus/hypothalamus (53) |
| Precuneus (32) - Caudate (99) |
| Precuneus (32) - Thalamus (45) |
| Precuneus (32) - "Middle temporal gyrus ([MTG], 56)" |
| Precuneus (32) - "Postcentral gyrus ([PoCG], 3)" |
| Precuneus (32) - "Left postcentral gyrus ([L PoCG], 9)" |
| Precuneus (40) - "Left postcentral gyrus ([L PoCG], 9)" |
| Precuneus (32) - "Paracentral lobule ([ParaCL], 2)" |
| Precuneus (32) - "Right postcentral gyrus ([R PoCG], 11)" |
| Precuneus (40) - "Right postcentral gyrus ([R PoCG], 11)" |
| Precuneus (32) - "Superior parietal lobule ([SPL], 27)" |
| Precuneus (40) - "Superior parietal lobule ([SPL], 27)" |
| Precuneus (32) - "Precentral gyrus ([PreCG], 66)" |
| Precuneus (32) - "Superior parietal lobule ([SPL], 80)" |
| Precuneus (40) - "Superior parietal lobule ([SPL], 80)" |
| Precuneus (32) - "Postcentral gyrus ([PoCG], 72)" |
| Precuneus (32) - "Calcarine gyrus ([CalcarineG], 16)" |
| Precuneus (32) - "Middle occipital gyrus ([MOG], 5)" |
| Precuneus (32) - "Middle temporal gyrus ([MTG], 62)" |
| Precuneus (32) - Cuneus (15) |
| Precuneus (40) - Cuneus (15) |
| Precuneus (32) - "Right middle occipital gyrus ([R MOG], 12)" |
| Precuneus (40) - "Right middle occipital gyrus ([R MOG], 12)" |
| Precuneus (32) - Fusiform gyrus (93) |
| Precuneus (40) - Fusiform gyrus (93) |
| Precuneus (32) - "Inferior occipital gyrus ([IOG], 20)" |
| Precuneus (40) - "Inferior occipital gyrus ([IOG], 20)" |
| Precuneus (32) - "Lingual gyrus ([LingualG], 8)" |
| Precuneus (32) - "Middle temporal gyrus ([MTG], 77)" |
| Precuneus (32) - "Inferior parietal lobule ([IPL], 68)" |
| Precuneus (32) - "Superior medial frontal gyrus ([SMFG], 43)" |
| Precuneus (40) - "Superior medial frontal gyrus ([SMFG], 43)" |
| Precuneus (32) - "Left inferior parietal lobue ([R IPL], 79)" |
| Precuneus (32) - "Superior frontal gyrus ([SFG], 96)" |
| Precuneus (32) - "Middle cingulate cortex ([MCC], 37)" |
| Precuneus (40) - "Middle cingulate cortex ([MCC], 37)" |
| Precuneus (32) - "Middle frontal gyrus ([MiFG], 38)" |
| Precuneus (40) - "Middle frontal gyrus ([MiFG], 38)" |

| |
|---|
| "Anterior cingulate cortex ([ACC], 23)" - Precuneus (32) |
| "Cerebellum ([CB], 13)" - Precuneus (32) |
| "Cerebellum ([CB], 18)" - Precuneus (32) |
| "Cerebellum ([CB], 4)" - Precuneus (32) |
| "Cerebellum ([CB], 7)" - Precuneus (32) |
| "Cerebellum ([CB], 18)" - Precuneus (40) |
| state3_jica4 |
| "Posterior cingulate cortex ([PCC], 94)" - Caudate (99) |
| "Posterior cingulate cortex ([PCC], 94)" - Thalamus (45) |
| "Posterior cingulate cortex ([PCC], 94)" - "Postcentral gyrus ([PoCG], 3)" |
| "Posterior cingulate cortex ([PCC], 94)" - "Left postcentral gyrus ([L PoCG], 9)" |
| "Posterior cingulate cortex ([PCC], 94)" - "Paracentral lobule ([ParaCL], 2)" |
| "Superior frontal gyrus ([SFG], 96)" - "Right postcentral gyrus ([R PoCG], 11)" |
| "Posterior cingulate cortex ([PCC], 94)" - "Right postcentral gyrus ([R PoCG], 11)" |
| "Superior frontal gyrus ([SFG], 96)" - "Superior parietal lobule ([SPL], 27)" |
| "Posterior cingulate cortex ([PCC], 94)" - "Superior parietal lobule ([SPL], 27)" |
| "Posterior cingulate cortex ([PCC], 94)" - "Calcarine gyrus ([CalcarineG], 16)" |
| "Superior medial frontal gyrus ([SMFG], 43)" - "Middle temporal gyrus ([MTG], 62)" |
| "Superior frontal gyrus ([SFG], 96)" - "Middle temporal gyrus ([MTG], 62)" |
| "Posterior cingulate cortex ([PCC], 94)" - "Middle temporal gyrus ([MTG], 62)" |
| "Superior frontal gyrus ([SFG], 96)" - Cuneus (15) |
| "Posterior cingulate cortex ([PCC], 94)" - Cuneus (15) |
| "Superior medial frontal gyrus ([SMFG], 43)" - "Right middle occipital gyrus ([R MOG], 12)" |
| "Superior frontal gyrus ([SFG], 96)" - "Right middle occipital gyrus ([R MOG], 12)" |
| "Posterior cingulate cortex ([PCC], 94)" - "Right middle occipital gyrus ([R MOG], 12)" |
| "Superior medial frontal gyrus ([SMFG], 43)" - "Inferior occipital gyrus ([IOG], 20)" |
| "Superior frontal gyrus ([SFG], 96)" - "Inferior occipital gyrus ([IOG], 20)" |
| "Posterior cingulate cortex ([PCC], 94)" - "Inferior occipital gyrus ([IOG], 20)" |
| "Cerebellum ([CB], 13)" - "Superior frontal gyrus ([SFG], 96)" |
| "Posterior cingulate cortex ([PCC], 94)" - "Middle cingulate cortex ([MCC], 37)" |
| "Posterior cingulate cortex ([PCC], 94)" - "Inferior frontal gyrus ([IFG], 67)" |
| "Posterior cingulate cortex ([PCC], 94)" - "Middle frontal gyrus ([MiFG], 38)" |
| "Posterior cingulate cortex ([PCC], 94)" - "Posterior cingulate cortex ([PCC], 71)" |
| "Cerebellum ([CB], 13)" - "Posterior cingulate cortex ([PCC], 94)" |
| state3_jica5 |
| "Cerebellum ([CB], 13)" - Caudate (69) |
| "Cerebellum ([CB], 7)" - Caudate (69) |
| "Cerebellum ([CB], 13)" - Subthalamus/hypothalamus (53) |
| "Cerebellum ([CB], 7)" - Subthalamus/hypothalamus (53) |
| "Cerebellum ([CB], 7)" - Caudate (99) |
| "Cerebellum ([CB], 13)" - "Postcentral gyrus ([PoCG], 3)" |
| "Cerebellum ([CB], 7)" - "Postcentral gyrus ([PoCG], 3)" |
| "Cerebellum ([CB], 13)" - "Left postcentral gyrus ([L PoCG], 9)" |
| "Cerebellum ([CB], 7)" - "Left postcentral gyrus ([L PoCG], 9)" |

| |
|---|
| "Cerebellum ([CB], 7)" - "Paracentral lobule ([ParaCL], 2)" |
| "Cerebellum ([CB], 13)" - "Right postcentral gyrus ([R PoCG], 11)" |
| "Cerebellum ([CB], 7)" - "Right postcentral gyrus ([R PoCG], 11)" |
| "Cerebellum ([CB], 13)" - "Superior parietal lobule ([SPL], 27)" |
| "Cerebellum ([CB], 7)" - "Superior parietal lobule ([SPL], 27)" |
| "Cerebellum ([CB], 13)" - "Superior parietal lobule ([SPL], 80)" |
| "Cerebellum ([CB], 18)" - "Superior parietal lobule ([SPL], 80)" |
| "Cerebellum ([CB], 7)" - "Superior parietal lobule ([SPL], 80)" |
| "Cerebellum ([CB], 7)" - "Postcentral gyrus ([PoCG], 72)" |
| "Cerebellum ([CB], 13)" - "Middle occipital gyrus ([MOG], 5)" |
| "Cerebellum ([CB], 7)" - "Middle occipital gyrus ([MOG], 5)" |
| "Cerebellum ([CB], 13)" - "Middle temporal gyrus ([MTG], 62)" |
| "Cerebellum ([CB], 13)" - Cuneus (15) |
| "Cerebellum ([CB], 7)" - Cuneus (15) |
| "Cerebellum ([CB], 13)" - "Right middle occipital gyrus ([R MOG], 12)" |
| "Cerebellum ([CB], 18)" - "Right middle occipital gyrus ([R MOG], 12)" |
| "Cerebellum ([CB], 7)" - "Right middle occipital gyrus ([R MOG], 12)" |
| "Cerebellum ([CB], 13)" - "Inferior occipital gyrus ([IOG], 20)" |
| "Cerebellum ([CB], 18)" - "Inferior occipital gyrus ([IOG], 20)" |
| "Cerebellum ([CB], 7)" - "Inferior occipital gyrus ([IOG], 20)" |
| "Cerebellum ([CB], 13)" - "Lingual gyrus ([LingualG], 8)" |
| "Cerebellum ([CB], 7)" - "Lingual gyrus ([LingualG], 8)" |
| "Cerebellum ([CB], 13)" - "Middle temporal gyrus ([MTG], 77)" |
| "Cerebellum ([CB], 7)" - "Middle temporal gyrus ([MTG], 77)" |
| "Cerebellum ([CB], 7)" - "Inferior parietal lobule ([IPL], 68)" |
| "Cerebellum ([CB], 13)" - "Superior medial frontal gyrus ([SMFG], 43)" |
| "Cerebellum ([CB], 13)" - "Superior frontal gyrus ([SFG], 96)" |
| "Cerebellum ([CB], 13)" - "Middle cingulate cortex ([MCC], 37)" |
| "Cerebellum ([CB], 7)" - "Middle cingulate cortex ([MCC], 37)" |
| "Cerebellum ([CB], 7)" - "Anterior cingulate cortex ([ACC], 23)" |
| "Cerebellum ([CB], 7)" - "Posterior cingulate cortex ([PCC], 71)" |
| "Cerebellum ([CB], 13)" - "Posterior cingulate cortex ([PCC], 94)" |
| "Cerebellum ([CB], 18)" - "Cerebellum ([CB], 13)" |
| "Cerebellum ([CB], 4)" - "Cerebellum ([CB], 13)" |
| "Cerebellum ([CB], 7)" - "Cerebellum ([CB], 13)" |
| "Cerebellum ([CB], 4)" - "Cerebellum ([CB], 18)" |
| "Cerebellum ([CB], 7)" - "Cerebellum ([CB], 4)" |
| state3_jica7 |
| "Postcentral gyrus ([PoCG], 3)" - Subthalamus/hypothalamus (53) |
| "Left postcentral gyrus ([L PoCG], 9)" - Subthalamus/hypothalamus (53) |
| "Paracentral lobule ([ParaCL], 2)" - Subthalamus/hypothalamus (53) |
| "Right postcentral gyrus ([R PoCG], 11)" - Subthalamus/hypothalamus (53) |
| "Superior parietal lobule ([SPL], 27)" - Subthalamus/hypothalamus (53) |
| "Calcarine gyrus ([CalcarineG], 16)" - Subthalamus/hypothalamus (53) |

| |
|---|
| "Middle occipital gyrus ([MOG], 5)" - Subthalamus/hypothalamus (53) |
| Cuneus (15) - Subthalamus/hypothalamus (53) |
| "Right middle occipital gyrus ([R MOG], 12)" - Subthalamus/hypothalamus (53) |
| Fusiform gyrus (93) - Subthalamus/hypothalamus (53) |
| "Inferior occipital gyrus ([IOG], 20)" - Subthalamus/hypothalamus (53) |
| "Lingual gyrus ([LingualG], 8)" - Subthalamus/hypothalamus (53) |
| "Inferior parietal lobule ([IPL], 68)" - Subthalamus/hypothalamus (53) |
| "Superior medial frontal gyrus ([SMFG], 43)" - Subthalamus/hypothalamus (53) |
| "Inferior parietal lobule ([IPL], 63)" - Subthalamus/hypothalamus (53) |
| "Superior frontal gyrus ([SFG], 96)" - Subthalamus/hypothalamus (53) |
| "Middle cingulate cortex ([MCC], 37)" - Subthalamus/hypothalamus (53) |
| "Middle frontal gyrus ([MiFG], 38)" - Subthalamus/hypothalamus (53) |
| "Cerebellum ([CB], 18)" - Subthalamus/hypothalamus (53) |
| "Postcentral gyrus ([PoCG], 3)" - Thalamus (45) |
| "Left postcentral gyrus ([L PoCG], 9)" - Thalamus (45) |
| "Paracentral lobule ([ParaCL], 2)" - Thalamus (45) |
| "Right postcentral gyrus ([R PoCG], 11)" - Thalamus (45) |
| "Superior parietal lobule ([SPL], 27)" - Thalamus (45) |
| "Postcentral gyrus ([PoCG], 72)" - Thalamus (45) |
| "Calcarine gyrus ([CalcarineG], 16)" - Thalamus (45) |
| "Middle occipital gyrus ([MOG], 5)" - Thalamus (45) |
| Cuneus (15) - Thalamus (45) |
| "Right middle occipital gyrus ([R MOG], 12)" - Thalamus (45) |
| Fusiform gyrus (93) - Thalamus (45) |
| "Inferior occipital gyrus ([IOG], 20)" - Thalamus (45) |
| "Lingual gyrus ([LingualG], 8)" - Thalamus (45) |
| "Inferior parietal lobule ([IPL], 68)" - Thalamus (45) |
| "Superior medial frontal gyrus ([SMFG], 43)" - Thalamus (45) |
| "Inferior parietal lobule ([IPL], 63)" - Thalamus (45) |
| "Superior frontal gyrus ([SFG], 96)" - Thalamus (45) |
| "Middle cingulate cortex ([MCC], 37)" - Thalamus (45) |
| "Middle frontal gyrus ([MiFG], 38)" - Thalamus (45) |
| "Posterior cingulate cortex ([PCC], 71)" - Thalamus (45) |
| "Cerebellum ([CB], 13)" - Thalamus (45) |
| "Cerebellum ([CB], 18)" - Thalamus (45) |
| "Cerebellum ([CB], 7)" - Thalamus (45) |
| state3_jica8 |
| "Middle temporal gyrus ([MTG], 62)" - "Middle temporal gyrus ([MTG], 56)" |
| "Middle temporal gyrus ([MTG], 62)" - "Postcentral gyrus ([PoCG], 3)" |
| "Middle temporal gyrus ([MTG], 77)" - "Postcentral gyrus ([PoCG], 3)" |
| "Middle temporal gyrus ([MTG], 62)" - "Left postcentral gyrus ([L PoCG], 9)" |
| "Middle temporal gyrus ([MTG], 77)" - "Left postcentral gyrus ([L PoCG], 9)" |
| "Middle temporal gyrus ([MTG], 62)" - "Paracentral lobule ([ParaCL], 2)" |
| "Middle temporal gyrus ([MTG], 77)" - "Paracentral lobule ([ParaCL], 2)" |

| |
|---|
| "Middle temporal gyrus ([MTG], 62)" - "Right postcentral gyrus ([R PoCG], 11)" |
| "Middle temporal gyrus ([MTG], 77)" - "Right postcentral gyrus ([R PoCG], 11)" |
| "Middle temporal gyrus ([MTG], 62)" - "Superior parietal lobule ([SPL], 27)" |
| "Middle temporal gyrus ([MTG], 77)" - "Superior parietal lobule ([SPL], 27)" |
| "Middle temporal gyrus ([MTG], 62)" - "Superior parietal lobule ([SPL], 80)" |
| "Middle temporal gyrus ([MTG], 62)" - "Postcentral gyrus ([PoCG], 72)" |
| "Middle temporal gyrus ([MTG], 77)" - "Postcentral gyrus ([PoCG], 72)" |
| "Middle temporal gyrus ([MTG], 62)" - "Calcarine gyrus ([CalcarineG], 16)" |
| "Middle temporal gyrus ([MTG], 62)" - "Middle occipital gyrus ([MOG], 5)" |
| Cuneus (15) - "Middle temporal gyrus ([MTG], 62)" |
| "Right middle occipital gyrus ([R MOG], 12)" - "Middle temporal gyrus ([MTG], 62)" |
| Fusiform gyrus (93) - "Middle temporal gyrus ([MTG], 62)" |
| "Inferior occipital gyrus ([IOG], 20)" - "Middle temporal gyrus ([MTG], 62)" |
| "Lingual gyrus ([LingualG], 8)" - "Middle temporal gyrus ([MTG], 62)" |
| "Middle temporal gyrus ([MTG], 77)" - "Middle temporal gyrus ([MTG], 62)" |
| "Inferior parietal lobule ([IPL], 68)" - "Middle temporal gyrus ([MTG], 62)" |
| Insula (33) - "Middle temporal gyrus ([MTG], 62)" |
| "Superior medial frontal gyrus ([SMFG], 43)" - "Middle temporal gyrus ([MTG], 62)" |
| "Right inferior frontal gyrus ([R IFG], 61)" - "Middle temporal gyrus ([MTG], 62)" |
| "Inferior parietal lobule ([IPL], 63)" - "Middle temporal gyrus ([MTG], 62)" |
| "Superior frontal gyrus ([SFG], 96)" - "Middle temporal gyrus ([MTG], 62)" |
| "Middle cingulate cortex ([MCC], 37)" - "Middle temporal gyrus ([MTG], 62)" |
| "Middle frontal gyrus ([MiFG], 38)" - "Middle temporal gyrus ([MTG], 62)" |
| "Anterior cingulate cortex ([ACC], 23)" - "Middle temporal gyrus ([MTG], 62)" |
| "Posterior cingulate cortex ([PCC], 71)" - "Middle temporal gyrus ([MTG], 62)" |
| "Posterior cingulate cortex ([PCC], 94)" - "Middle temporal gyrus ([MTG], 62)" |
| "Cerebellum ([CB], 13)" - "Middle temporal gyrus ([MTG], 62)" |
| "Cerebellum ([CB], 18)" - "Middle temporal gyrus ([MTG], 62)" |
| "Cerebellum ([CB], 7)" - "Middle temporal gyrus ([MTG], 62)" |
| "Middle temporal gyrus ([MTG], 77)" - Cuneus (15) |
| "Middle temporal gyrus ([MTG], 77)" - "Right middle occipital gyrus ([R MOG], 12)" |
| "Middle temporal gyrus ([MTG], 77)" - "Inferior occipital gyrus ([IOG], 20)" |
| "Inferior parietal lobule ([IPL], 68)" - "Middle temporal gyrus ([MTG], 77)" |
| "Inferior parietal lobule ([IPL], 63)" - "Middle temporal gyrus ([MTG], 77)" |
| "Middle cingulate cortex ([MCC], 37)" - "Middle temporal gyrus ([MTG], 77)" |
| "Middle frontal gyrus ([MiFG], 38)" - "Middle temporal gyrus ([MTG], 77)" |
| "Posterior cingulate cortex ([PCC], 71)" - "Middle temporal gyrus ([MTG], 77)" |
| "Cerebellum ([CB], 18)" - "Middle temporal gyrus ([MTG], 77)" |
| "Cerebellum ([CB], 7)" - "Middle temporal gyrus ([MTG], 77)" |
| state3_jica9 |
| "Paracentral lobule ([ParaCL], 2)" - Caudate (69) |
| Fusiform gyrus (93) - Caudate (69) |
| "Paracentral lobule ([ParaCL], 2)" - Thalamus (45) |
| "Paracentral lobule ([ParaCL], 2)" - "Postcentral gyrus ([PoCG], 3)" |

| |
|---|
| "Paracentral lobule ([ParaCL], 2)" - "Left postcentral gyrus ([L PoCG], 9)" |
| Cuneus (15) - "Left postcentral gyrus ([L PoCG], 9)" |
| "Right postcentral gyrus ([R PoCG], 11)" - "Paracentral lobule ([ParaCL], 2)" |
| "Superior parietal lobule ([SPL], 27)" - "Paracentral lobule ([ParaCL], 2)" |
| "Superior parietal lobule ([SPL], 80)" - "Paracentral lobule ([ParaCL], 2)" |
| "Postcentral gyrus ([PoCG], 72)" - "Paracentral lobule ([ParaCL], 2)" |
| "Calcarine gyrus ([CalcarineG], 16)" - "Paracentral lobule ([ParaCL], 2)" |
| "Middle occipital gyrus ([MOG], 5)" - "Paracentral lobule ([ParaCL], 2)" |
| "Middle temporal gyrus ([MTG], 62)" - "Paracentral lobule ([ParaCL], 2)" |
| Cuneus (15) - "Paracentral lobule ([ParaCL], 2)" |
| "Right middle occipital gyrus ([R MOG], 12)" - "Paracentral lobule ([ParaCL], 2)" |
| "Inferior occipital gyrus ([IOG], 20)" - "Paracentral lobule ([ParaCL], 2)" |
| "Lingual gyrus ([LingualG], 8)" - "Paracentral lobule ([ParaCL], 2)" |
| "Superior medial frontal gyrus ([SMFG], 43)" - "Paracentral lobule ([ParaCL], 2)" |
| "Middle cingulate cortex ([MCC], 37)" - "Paracentral lobule ([ParaCL], 2)" |
| "Anterior cingulate cortex ([ACC], 23)" - "Paracentral lobule ([ParaCL], 2)" |
| "Posterior cingulate cortex ([PCC], 71)" - "Paracentral lobule ([ParaCL], 2)" |
| "Posterior cingulate cortex ([PCC], 94)" - "Paracentral lobule ([ParaCL], 2)" |
| "Cerebellum ([CB], 18)" - "Paracentral lobule ([ParaCL], 2)" |
| "Cerebellum ([CB], 4)" - "Paracentral lobule ([ParaCL], 2)" |
| Cuneus (15) - "Right postcentral gyrus ([R PoCG], 11)" |
| Fusiform gyrus (93) - "Precentral gyrus ([PreCG], 66)" |
| state3_jica11 |
| "Superior temporal gyrus ([STG], 21)" - Caudate (69) |
| "Superior temporal gyrus ([STG], 21)" - Caudate (99) |
| "Middle temporal gyrus ([MTG], 56)" - Caudate (99) |
| "Superior temporal gyrus ([STG], 21)" - Thalamus (45) |
| "Postcentral gyrus ([PoCG], 3)" - "Superior temporal gyrus ([STG], 21)" |
| "Left postcentral gyrus ([L PoCG], 9)" - "Superior temporal gyrus ([STG], 21)" |
| "Paracentral lobule ([ParaCL], 2)" - "Superior temporal gyrus ([STG], 21)" |
| "Right postcentral gyrus ([R PoCG], 11)" - "Superior temporal gyrus ([STG], 21)" |
| "Superior parietal lobule ([SPL], 27)" - "Superior temporal gyrus ([STG], 21)" |
| "Precentral gyrus ([PreCG], 66)" - "Superior temporal gyrus ([STG], 21)" |
| "Superior parietal lobule ([SPL], 80)" - "Superior temporal gyrus ([STG], 21)" |
| "Postcentral gyrus ([PoCG], 72)" - "Superior temporal gyrus ([STG], 21)" |
| "Calcarine gyrus ([CalcarineG], 16)" - "Superior temporal gyrus ([STG], 21)" |
| "Middle occipital gyrus ([MOG], 5)" - "Superior temporal gyrus ([STG], 21)" |
| "Middle temporal gyrus ([MTG], 62)" - "Superior temporal gyrus ([STG], 21)" |
| Cuneus (15) - "Superior temporal gyrus ([STG], 21)" |
| "Right middle occipital gyrus ([R MOG], 12)" - "Superior temporal gyrus ([STG], 21)" |
| Fusiform gyrus (93) - "Superior temporal gyrus ([STG], 21)" |
| "Inferior occipital gyrus ([IOG], 20)" - "Superior temporal gyrus ([STG], 21)" |
| "Lingual gyrus ([LingualG], 8)" - "Superior temporal gyrus ([STG], 21)" |
| "Middle temporal gyrus ([MTG], 77)" - "Superior temporal gyrus ([STG], 21)" |

| |
|---|
| "Inferior parietal lobule ([IPL], 68)" - "Superior temporal gyrus ([STG], 21)" |
| Insula (33) - "Superior temporal gyrus ([STG], 21)" |
| "Superior medial frontal gyrus ([SMFG], 43)" - "Superior temporal gyrus ([STG], 21)" |
| "Right inferior frontal gyrus ([R IFG], 61)" - "Superior temporal gyrus ([STG], 21)" |
| "Inferior parietal lobule ([IPL], 63)" - "Superior temporal gyrus ([STG], 21)" |
| "Superior frontal gyrus ([SFG], 96)" - "Superior temporal gyrus ([STG], 21)" |
| "Middle cingulate cortex ([MCC], 37)" - "Superior temporal gyrus ([STG], 21)" |
| "Inferior frontal gyrus ([IFG], 67)" - "Superior temporal gyrus ([STG], 21)" |
| "Middle frontal gyrus ([MiFG], 38)" - "Superior temporal gyrus ([STG], 21)" |
| "Anterior cingulate cortex ([ACC], 23)" - "Superior temporal gyrus ([STG], 21)" |
| "Posterior cingulate cortex ([PCC], 71)" - "Superior temporal gyrus ([STG], 21)" |
| "Cerebellum ([CB], 13)" - "Superior temporal gyrus ([STG], 21)" |
| "Cerebellum ([CB], 18)" - "Superior temporal gyrus ([STG], 21)" |
| "Cerebellum ([CB], 7)" - "Superior temporal gyrus ([STG], 21)" |
| "Left postcentral gyrus ([L PoCG], 9)" - "Middle temporal gyrus ([MTG], 56)" |
| "Right postcentral gyrus ([R PoCG], 11)" - "Middle temporal gyrus ([MTG], 56)" |
| "Superior parietal lobule ([SPL], 27)" - "Middle temporal gyrus ([MTG], 56)" |
| "Calcarine gyrus ([CalcarineG], 16)" - "Middle temporal gyrus ([MTG], 56)" |
| Cuneus (15) - "Middle temporal gyrus ([MTG], 56)" |
| "Right middle occipital gyrus ([R MOG], 12)" - "Middle temporal gyrus ([MTG], 56)" |
| "Inferior occipital gyrus ([IOG], 20)" - "Middle temporal gyrus ([MTG], 56)" |
| Insula (33) - "Middle temporal gyrus ([MTG], 56)" |
| "Middle cingulate cortex ([MCC], 37)" - "Middle temporal gyrus ([MTG], 56)" |
| "Middle frontal gyrus ([MiFG], 38)" - "Middle temporal gyrus ([MTG], 56)" |
| state3_jica15 |
| "Middle frontal gyrus ([MiFG], 55)" - Caudate (69) |
| "Middle frontal gyrus ([MiFG], 55)" - Subthalamus/hypothalamus (53) |
| "Middle frontal gyrus ([MiFG], 55)" - Caudate (99) |
| "Middle frontal gyrus ([MiFG], 55)" - Thalamus (45) |
| "Middle frontal gyrus ([MiFG], 55)" - "Postcentral gyrus ([PoCG], 3)" |
| "Inferior frontal gyrus ([IFG], 70)" - "Left postcentral gyrus ([L PoCG], 9)" |
| "Middle frontal gyrus ([MiFG], 55)" - "Left postcentral gyrus ([L PoCG], 9)" |
| "Middle frontal gyrus ([MiFG], 55)" - "Paracentral lobule ([ParaCL], 2)" |
| "Inferior frontal gyrus ([IFG], 70)" - "Right postcentral gyrus ([R PoCG], 11)" |
| "Middle frontal gyrus ([MiFG], 55)" - "Right postcentral gyrus ([R PoCG], 11)" |
| "Middle frontal gyrus ([MiFG], 55)" - "Superior parietal lobule ([SPL], 27)" |
| "Middle frontal gyrus ([MiFG], 55)" - "Precentral gyrus ([PreCG], 66)" |
| "Middle frontal gyrus ([MiFG], 55)" - "Superior parietal lobule ([SPL], 80)" |
| "Middle frontal gyrus ([MiFG], 55)" - "Postcentral gyrus ([PoCG], 72)" |
| "Inferior frontal gyrus ([IFG], 67)" - "Postcentral gyrus ([PoCG], 72)" |
| "Middle frontal gyrus ([MiFG], 55)" - "Calcarine gyrus ([CalcarineG], 16)" |
| "Middle frontal gyrus ([MiFG], 55)" - "Middle occipital gyrus ([MOG], 5)" |
| "Middle frontal gyrus ([MiFG], 55)" - "Middle temporal gyrus ([MTG], 62)" |
| "Middle frontal gyrus ([MiFG], 55)" - Cuneus (15) |

| |
|---|
| "Inferior frontal gyrus ([IFG], 70)" - "Right middle occipital gyrus ([R MOG], 12)" |
| "Middle frontal gyrus ([MiFG], 55)" - "Right middle occipital gyrus ([R MOG], 12)" |
| "Middle frontal gyrus ([MiFG], 55)" - Fusiform gyrus (93) |
| "Inferior frontal gyrus ([IFG], 70)" - "Inferior occipital gyrus ([IOG], 20)" |
| "Middle frontal gyrus ([MiFG], 55)" - "Inferior occipital gyrus ([IOG], 20)" |
| "Middle frontal gyrus ([MiFG], 55)" - "Lingual gyrus ([LingualG], 8)" |
| "Middle frontal gyrus ([MiFG], 55)" - "Middle temporal gyrus ([MTG], 77)" |
| "Middle frontal gyrus ([MiFG], 55)" - "Inferior parietal lobule ([IPL], 68)" |
| "Middle frontal gyrus ([MiFG], 55)" - "Superior medial frontal gyrus ([SMFG], 43)" |
| "Left inferior parietal lobue ([R IPL], 79)" - "Middle frontal gyrus ([MiFG], 55)" |
| "Superior frontal gyrus ([SFG], 96)" - "Middle frontal gyrus ([MiFG], 55)" |
| "Middle cingulate cortex ([MCC], 37)" - "Middle frontal gyrus ([MiFG], 55)" |
| "Middle frontal gyrus ([MiFG], 38)" - "Middle frontal gyrus ([MiFG], 55)" |
| "Anterior cingulate cortex ([ACC], 23)" - "Middle frontal gyrus ([MiFG], 55)" |
| "Posterior cingulate cortex ([PCC], 71)" - "Middle frontal gyrus ([MiFG], 55)" |
| "Posterior cingulate cortex ([PCC], 94)" - "Middle frontal gyrus ([MiFG], 55)" |
| "Cerebellum ([CB], 13)" - "Middle frontal gyrus ([MiFG], 55)" |
| "Cerebellum ([CB], 18)" - "Middle frontal gyrus ([MiFG], 55)" |
| "Cerebellum ([CB], 4)" - "Middle frontal gyrus ([MiFG], 55)" |
| "Cerebellum ([CB], 7)" - "Middle frontal gyrus ([MiFG], 55)" |
| state3_jica17 |
| Insula (33) - Caudate (69) |
| "Middle frontal gyrus ([MiFG], 88)" - Caudate (69) |
| "Middle frontal gyrus ([MiFG], 88)" - Subthalamus/hypothalamus (53) |
| "Middle frontal gyrus ([MiFG], 88)" - Caudate (99) |
| "Middle frontal gyrus ([MiFG], 88)" - Thalamus (45) |
| Insula (33) - "Postcentral gyrus ([PoCG], 3)" |
| "Middle frontal gyrus ([MiFG], 88)" - "Postcentral gyrus ([PoCG], 3)" |
| Insula (33) - "Left postcentral gyrus ([L PoCG], 9)" |
| "Middle frontal gyrus ([MiFG], 88)" - "Left postcentral gyrus ([L PoCG], 9)" |
| "Middle frontal gyrus ([MiFG], 88)" - "Paracentral lobule ([ParaCL], 2)" |
| Insula (33) - "Right postcentral gyrus ([R PoCG], 11)" |
| "Middle frontal gyrus ([MiFG], 88)" - "Right postcentral gyrus ([R PoCG], 11)" |
| Insula (33) - "Superior parietal lobule ([SPL], 27)" |
| "Middle frontal gyrus ([MiFG], 88)" - "Superior parietal lobule ([SPL], 27)" |
| Insula (33) - "Precentral gyrus ([PreCG], 66)" |
| "Middle frontal gyrus ([MiFG], 88)" - "Precentral gyrus ([PreCG], 66)" |
| "Middle frontal gyrus ([MiFG], 88)" - "Superior parietal lobule ([SPL], 80)" |
| "Middle frontal gyrus ([MiFG], 88)" - "Postcentral gyrus ([PoCG], 72)" |
| "Middle frontal gyrus ([MiFG], 88)" - "Calcarine gyrus ([CalcarineG], 16)" |
| "Middle frontal gyrus ([MiFG], 88)" - "Middle occipital gyrus ([MOG], 5)" |
| "Middle frontal gyrus ([MiFG], 88)" - "Middle temporal gyrus ([MTG], 62)" |
| Insula (33) - Cuneus (15) |
| "Middle frontal gyrus ([MiFG], 88)" - Cuneus (15) |

| |
|---|
| Insula (33) - "Right middle occipital gyrus ([R MOG], 12)" |
| "Middle frontal gyrus ([MiFG], 88)" - "Right middle occipital gyrus ([R MOG], 12)" |
| Insula (33) - "Inferior occipital gyrus ([IOG], 20)" |
| "Middle frontal gyrus ([MiFG], 88)" - "Inferior occipital gyrus ([IOG], 20)" |
| "Middle frontal gyrus ([MiFG], 88)" - "Lingual gyrus ([LingualG], 8)" |
| "Middle frontal gyrus ([MiFG], 88)" - "Middle temporal gyrus ([MTG], 77)" |
| "Middle frontal gyrus ([MiFG], 88)" - "Inferior parietal lobule ([IPL], 68)" |
| "Superior frontal gyrus ([SFG], 96)" - Insula (33) |
| "Middle cingulate cortex ([MCC], 37)" - Insula (33) |
| "Middle frontal gyrus ([MiFG], 38)" - Insula (33) |
| "Cerebellum ([CB], 13)" - Insula (33) |
| "Cerebellum ([CB], 18)" - Insula (33) |
| "Cerebellum ([CB], 7)" - Insula (33) |
| "Middle frontal gyrus ([MiFG], 88)" - "Superior medial frontal gyrus ([SMFG], 43)" |
| "Middle frontal gyrus ([MiFG], 88)" - "Superior frontal gyrus ([SFG], 96)" |
| "Middle cingulate cortex ([MCC], 37)" - "Middle frontal gyrus ([MiFG], 88)" |
| "Middle frontal gyrus ([MiFG], 38)" - "Middle frontal gyrus ([MiFG], 88)" |
| "Anterior cingulate cortex ([ACC], 23)" - "Middle frontal gyrus ([MiFG], 88)" |
| "Posterior cingulate cortex ([PCC], 71)" - "Middle frontal gyrus ([MiFG], 88)" |
| "Posterior cingulate cortex ([PCC], 94)" - "Middle frontal gyrus ([MiFG], 88)" |
| "Cerebellum ([CB], 13)" - "Middle frontal gyrus ([MiFG], 88)" |
| "Cerebellum ([CB], 18)" - "Middle frontal gyrus ([MiFG], 88)" |
| "Cerebellum ([CB], 4)" - "Middle frontal gyrus ([MiFG], 88)" |
| "Cerebellum ([CB], 7)" - "Middle frontal gyrus ([MiFG], 88)" |
| state3_jica21 |
| "Inferior parietal lobule ([IPL], 68)" - Caudate (69) |
| "Middle frontal gyrus ([MiFG], 55)" - Caudate (69) |
| "Inferior parietal lobule ([IPL], 63)" - Caudate (69) |
| "Middle frontal gyrus ([MiFG], 55)" - Subthalamus/hypothalamus (53) |
| "Posterior cingulate cortex ([PCC], 71)" - "Inferior frontal gyrus ([IFG], 70)" |
| Precuneus (51) - "Middle frontal gyrus ([MiFG], 55)" |
| Precuneus (51) - "Inferior parietal lobule ([IPL], 63)" |
| Precuneus (32) - "Hippocampus ([HiPP], 83)" |
| Precuneus (51) - "Hippocampus ([HiPP], 83)" |
| "Anterior cingulate cortex ([ACC], 17)" - Precuneus (32) |
| state3_jica22 |
| "Right inferior frontal gyrus ([R IFG], 61)" - Caudate (69) |
| "Right inferior frontal gyrus ([R IFG], 61)" - Subthalamus/hypothalamus (53) |
| "Right inferior frontal gyrus ([R IFG], 61)" - Caudate (99) |
| "Right inferior frontal gyrus ([R IFG], 61)" - Thalamus (45) |
| "Right inferior frontal gyrus ([R IFG], 61)" - "Superior temporal gyrus ([STG], 21)" |
| "Right inferior frontal gyrus ([R IFG], 61)" - "Middle temporal gyrus ([MTG], 56)" |
| "Right inferior frontal gyrus ([R IFG], 61)" - "Postcentral gyrus ([PoCG], 3)" |
| "Right inferior frontal gyrus ([R IFG], 61)" - "Left postcentral gyrus ([L PoCG], 9)" |

| |
|---|
| "Right inferior frontal gyrus ([R IFG], 61)" - "Paracentral lobule ([ParaCL], 2)" |
| "Right inferior frontal gyrus ([R IFG], 61)" - "Right postcentral gyrus ([R PoCG], 11)" |
| "Right inferior frontal gyrus ([R IFG], 61)" - "Superior parietal lobule ([SPL], 27)" |
| "Right inferior frontal gyrus ([R IFG], 61)" - "Precentral gyrus ([PreCG], 66)" |
| "Right inferior frontal gyrus ([R IFG], 61)" - "Superior parietal lobule ([SPL], 80)" |
| "Right inferior frontal gyrus ([R IFG], 61)" - "Postcentral gyrus ([PoCG], 72)" |
| "Right inferior frontal gyrus ([R IFG], 61)" - "Calcarine gyrus ([CalcarineG], 16)" |
| "Right inferior frontal gyrus ([R IFG], 61)" - "Middle occipital gyrus ([MOG], 5)" |
| "Right inferior frontal gyrus ([R IFG], 61)" - "Middle temporal gyrus ([MTG], 62)" |
| "Right inferior frontal gyrus ([R IFG], 61)" - Cuneus (15) |
| "Right inferior frontal gyrus ([R IFG], 61)" - "Right middle occipital gyrus ([R MOG], 12)" |
| "Right inferior frontal gyrus ([R IFG], 61)" - Fusiform gyrus (93) |
| "Right inferior frontal gyrus ([R IFG], 61)" - "Inferior occipital gyrus ([IOG], 20)" |
| "Right inferior frontal gyrus ([R IFG], 61)" - "Lingual gyrus ([LingualG], 8)" |
| "Right inferior frontal gyrus ([R IFG], 61)" - "Middle temporal gyrus ([MTG], 77)" |
| "Right inferior frontal gyrus ([R IFG], 61)" - "Inferior parietal lobule ([IPL], 68)" |
| "Right inferior frontal gyrus ([R IFG], 61)" - "Superior medial frontal gyrus ([SMFG], 43)" |
| "Inferior parietal lobule ([IPL], 63)" - "Right inferior frontal gyrus ([R IFG], 61)" |
| "Left inferior parietal lobue ([R IPL], 79)" - "Right inferior frontal gyrus ([R IFG], 61)" |
| "Superior frontal gyrus ([SFG], 96)" - "Right inferior frontal gyrus ([R IFG], 61)" |
| "Left inferior parietal lobue ([L IPL], 81)" - "Right inferior frontal gyrus ([R IFG], 61)" |
| "Middle cingulate cortex ([MCC], 37)" - "Right inferior frontal gyrus ([R IFG], 61)" |
| "Inferior frontal gyrus ([IFG], 67)" - "Right inferior frontal gyrus ([R IFG], 61)" |
| "Middle frontal gyrus ([MiFG], 38)" - "Right inferior frontal gyrus ([R IFG], 61)" |
| "Hippocampus ([HiPP], 83)" - "Right inferior frontal gyrus ([R IFG], 61)" |
| "Anterior cingulate cortex ([ACC], 23)" - "Right inferior frontal gyrus ([R IFG], 61)" |
| "Posterior cingulate cortex ([PCC], 71)" - "Right inferior frontal gyrus ([R IFG], 61)" |
| "Posterior cingulate cortex ([PCC], 94)" - "Right inferior frontal gyrus ([R IFG], 61)" |
| "Cerebellum ([CB], 13)" - "Right inferior frontal gyrus ([R IFG], 61)" |
| "Cerebellum ([CB], 18)" - "Right inferior frontal gyrus ([R IFG], 61)" |
| "Cerebellum ([CB], 4)" - "Right inferior frontal gyrus ([R IFG], 61)" |
| "Cerebellum ([CB], 7)" - "Right inferior frontal gyrus ([R IFG], 61)" |
| state3_jica23 |
| "Hippocampus ([HiPP], 83)" - "Middle temporal gyrus ([MTG], 56)" |
| "Left inferior parietal lobue ([L IPL], 81)" - Insula (33) |
| "Hippocampus ([HiPP], 83)" - Insula (33) |
| "Anterior cingulate cortex ([ACC], 23)" - Insula (33) |
| "Cerebellum ([CB], 4)" - Insula (33) |
| "Middle frontal gyrus ([MiFG], 88)" - "Middle frontal gyrus ([MiFG], 55)" |
| Precuneus (40) - "Middle frontal gyrus ([MiFG], 55)" |
| "Posterior cingulate cortex ([PCC], 94)" - "Middle frontal gyrus ([MiFG], 55)" |
| Precuneus (40) - "Inferior parietal lobule ([IPL], 63)" |
| "Posterior cingulate cortex ([PCC], 94)" - "Inferior parietal lobule ([IPL], 63)" |
| Precuneus (40) - "Hippocampus ([HiPP], 48)" |

| |
|---|
| "Posterior cingulate cortex ([PCC], 94)" - "Hippocampus ([HiPP], 48)" |
| "Anterior cingulate cortex ([ACC], 17)" - "Left inferior parietal lobue ([L IPL], 81)" |
| Precuneus (40) - "Hippocampus ([HiPP], 83)" |
| "Anterior cingulate cortex ([ACC], 17)" - Precuneus (40) |
| "Posterior cingulate cortex ([PCC], 94)" - "Anterior cingulate cortex ([ACC], 17)" |
| state3_jica24 |
| "Supplementary motor area ([SMA], 84)" - Caudate (69) |
| "Supplementary motor area ([SMA], 84)" - Subthalamus/hypothalamus (53) |
| "Supplementary motor area ([SMA], 84)" - Caudate (99) |
| "Supplementary motor area ([SMA], 84)" - Thalamus (45) |
| "Supplementary motor area ([SMA], 84)" - "Superior temporal gyrus ([STG], 21)" |
| "Supplementary motor area ([SMA], 84)" - "Middle temporal gyrus ([MTG], 56)" |
| "Supplementary motor area ([SMA], 84)" - "Postcentral gyrus ([PoCG], 3)" |
| "Supplementary motor area ([SMA], 84)" - "Left postcentral gyrus ([L PoCG], 9)" |
| "Supplementary motor area ([SMA], 84)" - "Paracentral lobule ([ParaCL], 2)" |
| "Supplementary motor area ([SMA], 84)" - "Right postcentral gyrus ([R PoCG], 11)" |
| "Supplementary motor area ([SMA], 84)" - "Superior parietal lobule ([SPL], 27)" |
| "Supplementary motor area ([SMA], 84)" - "Precentral gyrus ([PreCG], 66)" |
| "Supplementary motor area ([SMA], 84)" - "Superior parietal lobule ([SPL], 80)" |
| "Supplementary motor area ([SMA], 84)" - "Postcentral gyrus ([PoCG], 72)" |
| "Supplementary motor area ([SMA], 84)" - "Calcarine gyrus ([CalcarineG], 16)" |
| "Supplementary motor area ([SMA], 84)" - "Middle occipital gyrus ([MOG], 5)" |
| "Supplementary motor area ([SMA], 84)" - "Middle temporal gyrus ([MTG], 62)" |
| "Supplementary motor area ([SMA], 84)" - Cuneus (15) |
| "Supplementary motor area ([SMA], 84)" - "Right middle occipital gyrus ([R MOG], 12)" |
| "Supplementary motor area ([SMA], 84)" - Fusiform gyrus (93) |
| "Supplementary motor area ([SMA], 84)" - "Inferior occipital gyrus ([IOG], 20)" |
| "Supplementary motor area ([SMA], 84)" - "Lingual gyrus ([LingualG], 8)" |
| "Supplementary motor area ([SMA], 84)" - "Middle temporal gyrus ([MTG], 77)" |
| "Supplementary motor area ([SMA], 84)" - "Inferior parietal lobule ([IPL], 68)" |
| "Supplementary motor area ([SMA], 84)" - "Superior medial frontal gyrus ([SMFG], 43)" |
| "Supplementary motor area ([SMA], 84)" - "Left inferior parietal lobue ([R IPL], 79)" |
| "Superior frontal gyrus ([SFG], 96)" - "Supplementary motor area ([SMA], 84)" |
| "Middle cingulate cortex ([MCC], 37)" - "Supplementary motor area ([SMA], 84)" |
| "Middle frontal gyrus ([MiFG], 38)" - "Supplementary motor area ([SMA], 84)" |
| "Anterior cingulate cortex ([ACC], 23)" - "Supplementary motor area ([SMA], 84)" |
| "Posterior cingulate cortex ([PCC], 71)" - "Supplementary motor area ([SMA], 84)" |
| "Posterior cingulate cortex ([PCC], 94)" - "Supplementary motor area ([SMA], 84)" |
| "Cerebellum ([CB], 13)" - "Supplementary motor area ([SMA], 84)" |
| "Cerebellum ([CB], 18)" - "Supplementary motor area ([SMA], 84)" |
| "Cerebellum ([CB], 4)" - "Supplementary motor area ([SMA], 84)" |
| "Cerebellum ([CB], 7)" - "Supplementary motor area ([SMA], 84)" |
| state3_jica28 |
| "Middle temporal gyrus ([MTG], 77)" - "Paracentral lobule ([ParaCL], 2)" |

| |
|---|
| "Superior frontal gyrus ([SFG], 96)" - "Paracentral lobule ([ParaCL], 2)" |
| "Middle frontal gyrus ([MiFG], 38)" - "Paracentral lobule ([ParaCL], 2)" |
| "Cerebellum ([CB], 18)" - Insula (33) |
| "Cerebellum ([CB], 7)" - "Right inferior frontal gyrus ([R IFG], 61)" |
| state3_jica30 |
| "Superior medial frontal gyrus ([SMFG], 43)" - "Superior temporal gyrus ([STG], 21)" |
| "Hippocampus ([HiPP], 83)" - "Lingual gyrus ([LingualG], 8)" |
| "Hippocampus ([HiPP], 83)" - "Middle temporal gyrus ([MTG], 77)" |
| "Right inferior frontal gyrus ([R IFG], 61)" - "Inferior parietal lobule ([IPL], 68)" |
| "Middle frontal gyrus ([MiFG], 88)" - "Inferior frontal gyrus ([IFG], 70)" |
| "Middle frontal gyrus ([MiFG], 88)" - "Inferior parietal lobule ([IPL], 63)" |
| state3_jica31 |
| "Middle occipital gyrus ([MOG], 5)" - "Postcentral gyrus ([PoCG], 72)" |
| "Right middle occipital gyrus ([R MOG], 12)" - "Postcentral gyrus ([PoCG], 72)" |
| "Inferior occipital gyrus ([IOG], 20)" - "Postcentral gyrus ([PoCG], 72)" |
| "Middle temporal gyrus ([MTG], 77)" - "Calcarine gyrus ([CalcarineG], 16)" |
| Precuneus (51) - "Right inferior frontal gyrus ([R IFG], 61)" |
| state4_jica2 |
| "Left postcentral gyrus ([L PoCG], 9)" - Caudate (69) |
| "Right postcentral gyrus ([R PoCG], 11)" - Caudate (69) |
| "Superior parietal lobule ([SPL], 80)" - "Left postcentral gyrus ([L PoCG], 9)" |
| "Calcarine gyrus ([CalcarineG], 16)" - "Left postcentral gyrus ([L PoCG], 9)" |
| "Middle occipital gyrus ([MOG], 5)" - "Left postcentral gyrus ([L PoCG], 9)" |
| Cuneus (15) - "Left postcentral gyrus ([L PoCG], 9)" |
| "Right middle occipital gyrus ([R MOG], 12)" - "Left postcentral gyrus ([L PoCG], 9)" |
| "Inferior occipital gyrus ([IOG], 20)" - "Left postcentral gyrus ([L PoCG], 9)" |
| "Lingual gyrus ([LingualG], 8)" - "Left postcentral gyrus ([L PoCG], 9)" |
| "Middle temporal gyrus ([MTG], 77)" - "Left postcentral gyrus ([L PoCG], 9)" |
| "Superior medial frontal gyrus ([SMFG], 43)" - "Left postcentral gyrus ([L PoCG], 9)" |
| "Left inferior parietal lobue ([R IPL], 79)" - "Left postcentral gyrus ([L PoCG], 9)" |
| "Hippocampus ([HiPP], 48)" - "Left postcentral gyrus ([L PoCG], 9)" |
| "Left inferior parietal lobue ([L IPL], 81)" - "Left postcentral gyrus ([L PoCG], 9)" |
| "Middle cingulate cortex ([MCC], 37)" - "Left postcentral gyrus ([L PoCG], 9)" |
| "Anterior cingulate cortex ([ACC], 23)" - "Left postcentral gyrus ([L PoCG], 9)" |
| "Anterior cingulate cortex ([ACC], 17)" - "Left postcentral gyrus ([L PoCG], 9)" |
| "Posterior cingulate cortex ([PCC], 94)" - "Left postcentral gyrus ([L PoCG], 9)" |
| "Superior parietal lobule ([SPL], 80)" - "Paracentral lobule ([ParaCL], 2)" |
| "Calcarine gyrus ([CalcarineG], 16)" - "Paracentral lobule ([ParaCL], 2)" |
| "Middle occipital gyrus ([MOG], 5)" - "Paracentral lobule ([ParaCL], 2)" |
| Cuneus (15) - "Paracentral lobule ([ParaCL], 2)" |
| "Right middle occipital gyrus ([R MOG], 12)" - "Paracentral lobule ([ParaCL], 2)" |
| "Inferior occipital gyrus ([IOG], 20)" - "Paracentral lobule ([ParaCL], 2)" |
| "Lingual gyrus ([LingualG], 8)" - "Paracentral lobule ([ParaCL], 2)" |
| "Superior medial frontal gyrus ([SMFG], 43)" - "Paracentral lobule ([ParaCL], 2)" |

| |
|---|
| "Left inferior parietal lobue ([R IPL], 79)" - "Paracentral lobule ([ParaCL], 2)" |
| "Anterior cingulate cortex ([ACC], 23)" - "Paracentral lobule ([ParaCL], 2)" |
| "Posterior cingulate cortex ([PCC], 94)" - "Paracentral lobule ([ParaCL], 2)" |
| "Superior parietal lobule ([SPL], 80)" - "Right postcentral gyrus ([R PoCG], 11)" |
| "Calcarine gyrus ([CalcarineG], 16)" - "Right postcentral gyrus ([R PoCG], 11)" |
| "Middle occipital gyrus ([MOG], 5)" - "Right postcentral gyrus ([R PoCG], 11)" |
| Cuneus (15) - "Right postcentral gyrus ([R PoCG], 11)" |
| "Right middle occipital gyrus ([R MOG], 12)" - "Right postcentral gyrus ([R PoCG], 11)" |
| "Inferior occipital gyrus ([IOG], 20)" - "Right postcentral gyrus ([R PoCG], 11)" |
| "Lingual gyrus ([LingualG], 8)" - "Right postcentral gyrus ([R PoCG], 11)" |
| "Middle temporal gyrus ([MTG], 77)" - "Right postcentral gyrus ([R PoCG], 11)" |
| "Superior medial frontal gyrus ([SMFG], 43)" - "Right postcentral gyrus ([R PoCG], 11)" |
| "Left inferior parietal lobue ([R IPL], 79)" - "Right postcentral gyrus ([R PoCG], 11)" |
| "Hippocampus ([HiPP], 48)" - "Right postcentral gyrus ([R PoCG], 11)" |
| "Left inferior parietal lobue ([L IPL], 81)" - "Right postcentral gyrus ([R PoCG], 11)" |
| "Middle cingulate cortex ([MCC], 37)" - "Right postcentral gyrus ([R PoCG], 11)" |
| "Anterior cingulate cortex ([ACC], 23)" - "Right postcentral gyrus ([R PoCG], 11)" |
| "Posterior cingulate cortex ([PCC], 71)" - "Right postcentral gyrus ([R PoCG], 11)" |
| "Anterior cingulate cortex ([ACC], 17)" - "Right postcentral gyrus ([R PoCG], 11)" |
| "Posterior cingulate cortex ([PCC], 94)" - "Right postcentral gyrus ([R PoCG], 11)" |
| "Superior parietal lobule ([SPL], 80)" - "Superior parietal lobule ([SPL], 27)" |
| state4_jica9 |
| Precuneus (32) - Caudate (69) |
| Precuneus (32) - "Precentral gyrus ([PreCG], 66)" |
| Precuneus (51) - "Precentral gyrus ([PreCG], 66)" |
| Precuneus (32) - "Superior parietal lobule ([SPL], 80)" |
| Precuneus (40) - "Superior parietal lobule ([SPL], 80)" |
| Precuneus (51) - "Superior parietal lobule ([SPL], 80)" |
| Precuneus (32) - "Calcarine gyrus ([CalcarineG], 16)" |
| Precuneus (51) - "Calcarine gyrus ([CalcarineG], 16)" |
| Precuneus (32) - "Middle occipital gyrus ([MOG], 5)" |
| Precuneus (51) - "Middle occipital gyrus ([MOG], 5)" |
| Precuneus (32) - Cuneus (15) |
| Precuneus (40) - Cuneus (15) |
| Precuneus (51) - Cuneus (15) |
| Precuneus (32) - "Right middle occipital gyrus ([R MOG], 12)" |
| Precuneus (40) - "Right middle occipital gyrus ([R MOG], 12)" |
| Precuneus (51) - "Right middle occipital gyrus ([R MOG], 12)" |
| Precuneus (32) - "Inferior occipital gyrus ([IOG], 20)" |
| Precuneus (40) - "Inferior occipital gyrus ([IOG], 20)" |
| Precuneus (51) - "Inferior occipital gyrus ([IOG], 20)" |
| Precuneus (32) - "Lingual gyrus ([LingualG], 8)" |
| Precuneus (40) - "Lingual gyrus ([LingualG], 8)" |
| Precuneus (51) - "Lingual gyrus ([LingualG], 8)" |

| |
|---|
| Precuneus (32) - "Middle temporal gyrus ([MTG], 77)" |
| Precuneus (40) - "Middle temporal gyrus ([MTG], 77)" |
| Precuneus (51) - "Middle temporal gyrus ([MTG], 77)" |
| Precuneus (32) - "Superior medial frontal gyrus ([SMFG], 43)" |
| Precuneus (40) - "Superior medial frontal gyrus ([SMFG], 43)" |
| Precuneus (51) - "Superior medial frontal gyrus ([SMFG], 43)" |
| Precuneus (32) - "Middle frontal gyrus ([MiFG], 55)" |
| Precuneus (32) - "Left inferior parietal lobue ([R IPL], 79)" |
| Precuneus (40) - "Left inferior parietal lobue ([R IPL], 79)" |
| Precuneus (51) - "Left inferior parietal lobue ([R IPL], 79)" |
| Precuneus (32) - "Superior frontal gyrus ([SFG], 96)" |
| Precuneus (51) - "Superior frontal gyrus ([SFG], 96)" |
| Precuneus (32) - "Hippocampus ([HiPP], 48)" |
| Precuneus (32) - "Left inferior parietal lobue ([L IPL], 81)" |
| Precuneus (51) - "Left inferior parietal lobue ([L IPL], 81)" |
| Precuneus (32) - "Middle cingulate cortex ([MCC], 37)" |
| Precuneus (51) - "Middle cingulate cortex ([MCC], 37)" |
| "Anterior cingulate cortex ([ACC], 23)" - Precuneus (32) |
| "Anterior cingulate cortex ([ACC], 17)" - Precuneus (32) |
| "Posterior cingulate cortex ([PCC], 94)" - Precuneus (32) |
| "Anterior cingulate cortex ([ACC], 23)" - Precuneus (40) |
| Precuneus (51) - "Anterior cingulate cortex ([ACC], 23)" |
| Precuneus (51) - "Anterior cingulate cortex ([ACC], 17)" |
| "Posterior cingulate cortex ([PCC], 94)" - Precuneus (51) |
| state4_jica15 |
| "Postcentral gyrus ([PoCG], 3)" - Subthalamus/hypothalamus (53) |
| "Left postcentral gyrus ([L PoCG], 9)" - Subthalamus/hypothalamus (53) |
| "Paracentral lobule ([ParaCL], 2)" - Subthalamus/hypothalamus (53) |
| "Right postcentral gyrus ([R PoCG], 11)" - Subthalamus/hypothalamus (53) |
| "Superior parietal lobule ([SPL], 27)" - Subthalamus/hypothalamus (53) |
| "Cerebellum ([CB], 13)" - Subthalamus/hypothalamus (53) |
| "Cerebellum ([CB], 18)" - Subthalamus/hypothalamus (53) |
| "Postcentral gyrus ([PoCG], 3)" - Putamen (98) |
| "Left postcentral gyrus ([L PoCG], 9)" - Putamen (98) |
| "Paracentral lobule ([ParaCL], 2)" - Putamen (98) |
| "Right postcentral gyrus ([R PoCG], 11)" - Putamen (98) |
| "Superior parietal lobule ([SPL], 27)" - Putamen (98) |
| "Postcentral gyrus ([PoCG], 3)" - Caudate (99) |
| "Left postcentral gyrus ([L PoCG], 9)" - Caudate (99) |
| "Paracentral lobule ([ParaCL], 2)" - Caudate (99) |
| "Right postcentral gyrus ([R PoCG], 11)" - Caudate (99) |
| "Middle temporal gyrus ([MTG], 56)" - Thalamus (45) |
| "Postcentral gyrus ([PoCG], 3)" - Thalamus (45) |
| "Left postcentral gyrus ([L PoCG], 9)" - Thalamus (45) |

| |
|---|
| "Paracentral lobule ([ParaCL], 2)" - Thalamus (45) |
| "Right postcentral gyrus ([R PoCG], 11)" - Thalamus (45) |
| "Superior parietal lobule ([SPL], 27)" - Thalamus (45) |
| "Cerebellum ([CB], 13)" - Thalamus (45) |
| "Cerebellum ([CB], 18)" - Thalamus (45) |
| "Cerebellum ([CB], 13)" - "Left postcentral gyrus ([L PoCG], 9)" |
| "Cerebellum ([CB], 13)" - "Right postcentral gyrus ([R PoCG], 11)" |
| "Cerebellum ([CB], 4)" - "Right postcentral gyrus ([R PoCG], 11)" |
| "Cerebellum ([CB], 13)" - "Superior parietal lobule ([SPL], 27)" |
| "Cerebellum ([CB], 4)" - "Cerebellum ([CB], 13)" |
| state4_jica18 |
| "Middle frontal gyrus ([MiFG], 55)" - Caudate (69) |
| "Inferior frontal gyrus ([IFG], 67)" - Caudate (69) |
| "Inferior frontal gyrus ([IFG], 70)" - "Superior parietal lobule ([SPL], 80)" |
| "Middle frontal gyrus ([MiFG], 55)" - "Superior parietal lobule ([SPL], 80)" |
| "Left inferior parietal lobue ([L IPL], 81)" - "Superior parietal lobule ([SPL], 80)" |
| "Inferior frontal gyrus ([IFG], 67)" - "Superior parietal lobule ([SPL], 80)" |
| "Middle frontal gyrus ([MiFG], 55)" - "Calcarine gyrus ([CalcarineG], 16)" |
| "Inferior frontal gyrus ([IFG], 67)" - "Calcarine gyrus ([CalcarineG], 16)" |
| "Inferior frontal gyrus ([IFG], 70)" - "Middle occipital gyrus ([MOG], 5)" |
| "Middle frontal gyrus ([MiFG], 55)" - "Middle occipital gyrus ([MOG], 5)" |
| "Inferior frontal gyrus ([IFG], 67)" - "Middle occipital gyrus ([MOG], 5)" |
| "Inferior frontal gyrus ([IFG], 70)" - Cuneus (15) |
| "Middle frontal gyrus ([MiFG], 55)" - Cuneus (15) |
| "Left inferior parietal lobue ([L IPL], 81)" - Cuneus (15) |
| "Inferior frontal gyrus ([IFG], 67)" - Cuneus (15) |
| "Inferior frontal gyrus ([IFG], 70)" - "Right middle occipital gyrus ([R MOG], 12)" |
| "Middle frontal gyrus ([MiFG], 55)" - "Right middle occipital gyrus ([R MOG], 12)" |
| "Left inferior parietal lobue ([L IPL], 81)" - "Right middle occipital gyrus ([R MOG], 12)" |
| "Inferior frontal gyrus ([IFG], 67)" - "Right middle occipital gyrus ([R MOG], 12)" |
| "Middle frontal gyrus ([MiFG], 55)" - Fusiform gyrus (93) |
| "Inferior frontal gyrus ([IFG], 70)" - "Inferior occipital gyrus ([IOG], 20)" |
| "Middle frontal gyrus ([MiFG], 55)" - "Inferior occipital gyrus ([IOG], 20)" |
| "Left inferior parietal lobue ([L IPL], 81)" - "Inferior occipital gyrus ([IOG], 20)" |
| "Inferior frontal gyrus ([IFG], 67)" - "Inferior occipital gyrus ([IOG], 20)" |
| "Inferior frontal gyrus ([IFG], 70)" - "Lingual gyrus ([LingualG], 8)" |
| "Middle frontal gyrus ([MiFG], 55)" - "Lingual gyrus ([LingualG], 8)" |
| "Inferior frontal gyrus ([IFG], 67)" - "Lingual gyrus ([LingualG], 8)" |
| "Middle frontal gyrus ([MiFG], 55)" - "Middle temporal gyrus ([MTG], 77)" |
| "Inferior frontal gyrus ([IFG], 67)" - "Middle temporal gyrus ([MTG], 77)" |
| "Middle frontal gyrus ([MiFG], 55)" - "Superior medial frontal gyrus ([SMFG], 43)" |
| "Inferior frontal gyrus ([IFG], 67)" - "Superior medial frontal gyrus ([SMFG], 43)" |
| "Left inferior parietal lobue ([R IPL], 79)" - "Middle frontal gyrus ([MiFG], 55)" |
| "Middle cingulate cortex ([MCC], 37)" - "Middle frontal gyrus ([MiFG], 55)" |

| |
|---|
| "Anterior cingulate cortex ([ACC], 23)" - "Middle frontal gyrus ([MiFG], 55)" |
| "Posterior cingulate cortex ([PCC], 94)" - "Middle frontal gyrus ([MiFG], 55)" |
| "Middle cingulate cortex ([MCC], 37)" - "Left inferior parietal lobue ([L IPL], 81)" |
| "Inferior frontal gyrus ([IFG], 67)" - "Middle cingulate cortex ([MCC], 37)" |
| "Anterior cingulate cortex ([ACC], 23)" - "Inferior frontal gyrus ([IFG], 67)" |
| "Posterior cingulate cortex ([PCC], 94)" - "Inferior frontal gyrus ([IFG], 67)" |
| state4_jica23 |
| "Middle frontal gyrus ([MiFG], 38)" - Caudate (69) |
| "Middle frontal gyrus ([MiFG], 38)" - Subthalamus/hypothalamus (53) |
| "Middle frontal gyrus ([MiFG], 38)" - Putamen (98) |
| "Middle frontal gyrus ([MiFG], 38)" - Caudate (99) |
| "Middle frontal gyrus ([MiFG], 38)" - Thalamus (45) |
| "Middle frontal gyrus ([MiFG], 38)" - "Precentral gyrus ([PreCG], 66)" |
| "Middle frontal gyrus ([MiFG], 88)" - "Superior parietal lobule ([SPL], 80)" |
| "Middle cingulate cortex ([MCC], 37)" - "Superior parietal lobule ([SPL], 80)" |
| "Middle frontal gyrus ([MiFG], 38)" - "Superior parietal lobule ([SPL], 80)" |
| "Middle frontal gyrus ([MiFG], 38)" - "Calcarine gyrus ([CalcarineG], 16)" |
| "Middle frontal gyrus ([MiFG], 38)" - "Middle occipital gyrus ([MOG], 5)" |
| "Middle frontal gyrus ([MiFG], 38)" - "Middle temporal gyrus ([MTG], 62)" |
| "Middle frontal gyrus ([MiFG], 88)" - Cuneus (15) |
| "Middle frontal gyrus ([MiFG], 38)" - Cuneus (15) |
| "Middle frontal gyrus ([MiFG], 88)" - "Right middle occipital gyrus ([R MOG], 12)" |
| "Middle frontal gyrus ([MiFG], 38)" - "Right middle occipital gyrus ([R MOG], 12)" |
| "Middle frontal gyrus ([MiFG], 88)" - "Inferior occipital gyrus ([IOG], 20)" |
| "Middle frontal gyrus ([MiFG], 38)" - "Inferior occipital gyrus ([IOG], 20)" |
| "Middle frontal gyrus ([MiFG], 88)" - "Lingual gyrus ([LingualG], 8)" |
| "Middle frontal gyrus ([MiFG], 38)" - "Lingual gyrus ([LingualG], 8)" |
| "Middle frontal gyrus ([MiFG], 88)" - "Middle temporal gyrus ([MTG], 77)" |
| "Middle frontal gyrus ([MiFG], 38)" - "Middle temporal gyrus ([MTG], 77)" |
| "Middle frontal gyrus ([MiFG], 38)" - "Superior medial frontal gyrus ([SMFG], 43)" |
| "Middle frontal gyrus ([MiFG], 38)" - "Middle frontal gyrus ([MiFG], 55)" |
| "Middle frontal gyrus ([MiFG], 38)" - "Left inferior parietal lobue ([R IPL], 79)" |
| "Middle frontal gyrus ([MiFG], 38)" - "Superior frontal gyrus ([SFG], 96)" |
| "Middle frontal gyrus ([MiFG], 38)" - "Hippocampus ([HiPP], 48)" |
| "Middle frontal gyrus ([MiFG], 38)" - "Left inferior parietal lobue ([L IPL], 81)" |
| "Middle frontal gyrus ([MiFG], 38)" - "Middle cingulate cortex ([MCC], 37)" |
| "Hippocampus ([HiPP], 83)" - "Middle frontal gyrus ([MiFG], 38)" |
| "Anterior cingulate cortex ([ACC], 23)" - "Middle frontal gyrus ([MiFG], 38)" |
| "Posterior cingulate cortex ([PCC], 71)" - "Middle frontal gyrus ([MiFG], 38)" |
| "Anterior cingulate cortex ([ACC], 17)" - "Middle frontal gyrus ([MiFG], 38)" |
| "Posterior cingulate cortex ([PCC], 94)" - "Middle frontal gyrus ([MiFG], 38)" |
| state4_jica30 |
| Precuneus (51) - "Middle occipital gyrus ([MOG], 5)" |
| Precuneus (32) - Cuneus (15) |

| |
|---|
| Precuneus (51) - Cuneus (15) |
| Precuneus (32) - "Right middle occipital gyrus ([R MOG], 12)" |
| Precuneus (51) - "Right middle occipital gyrus ([R MOG], 12)" |
| Precuneus (51) - "Inferior occipital gyrus ([IOG], 20)" |
| Precuneus (51) - "Superior frontal gyrus ([SFG], 96)" |
| state4_jica31 |
| "Right inferior frontal gyrus ([R IFG], 61)" - "Superior temporal gyrus ([STG], 21)" |
| "Middle temporal gyrus ([MTG], 62)" - "Middle temporal gyrus ([MTG], 56)" |
| "Right inferior frontal gyrus ([R IFG], 61)" - "Middle temporal gyrus ([MTG], 56)" |
| "Right inferior frontal gyrus ([R IFG], 61)" - "Superior parietal lobule ([SPL], 27)" |
| "Right inferior frontal gyrus ([R IFG], 61)" - "Paracentral lobule ([ParaCL], 54)" |
| "Middle temporal gyrus ([MTG], 62)" - "Precentral gyrus ([PreCG], 66)" |
| "Superior medial frontal gyrus ([SMFG], 43)" - "Middle temporal gyrus ([MTG], 62)" |
| "Left inferior parietal lobue ([R IPL], 79)" - "Middle temporal gyrus ([MTG], 62)" |
| "Superior frontal gyrus ([SFG], 96)" - "Middle temporal gyrus ([MTG], 62)" |
| "Middle frontal gyrus ([MiFG], 88)" - "Middle temporal gyrus ([MTG], 62)" |
| Precuneus (40) - "Middle temporal gyrus ([MTG], 62)" |
| "Posterior cingulate cortex ([PCC], 94)" - "Middle temporal gyrus ([MTG], 62)" |
| "Cerebellum ([CB], 18)" - "Middle temporal gyrus ([MTG], 62)" |
| "Cerebellum ([CB], 7)" - "Middle temporal gyrus ([MTG], 62)" |
| "Right inferior frontal gyrus ([R IFG], 61)" - "Inferior frontal gyrus ([IFG], 70)" |
| "Left inferior parietal lobue ([L IPL], 81)" - "Inferior frontal gyrus ([IFG], 70)" |
| "Middle frontal gyrus ([MiFG], 88)" - "Right inferior frontal gyrus ([R IFG], 61)" |
| "Posterior cingulate cortex ([PCC], 94)" - "Right inferior frontal gyrus ([R IFG], 61)" |
| "Cerebellum ([CB], 18)" - "Right inferior frontal gyrus ([R IFG], 61)" |
| "Cerebellum ([CB], 7)" - "Right inferior frontal gyrus ([R IFG], 61)" |
| "Cerebellum ([CB], 7)" - "Middle frontal gyrus ([MiFG], 88)" |
| "Cerebellum ([CB], 7)" - "Cerebellum ([CB], 13)" |
| state4_jica35 |
| "Middle cingulate cortex ([MCC], 37)" - Subthalamus/hypothalamus (53) |
| "Inferior frontal gyrus ([IFG], 67)" - Thalamus (45) |
| "Cerebellum ([CB], 4)" - "Middle temporal gyrus ([MTG], 62)" |
